# Effects of drinking water, sanitation, handwashing, and nutritional interventions on immune status in young children: a cluster-randomized controlled trial in rural Bangladesh

**DOI:** 10.1101/2021.11.10.21266206

**Authors:** Audrie Lin, Andrew N. Mertens, Sophia Tan, Md. Ziaur Rahman, Lisa Hester, Lisa Kim, Benjamin F. Arnold, Md. Rabiul Karim, Sunny Shahriar, Shahjahan Ali, Abul K. Shoab, Md. Saheen Hossen, Palash Mutsuddi, Syeda Luthfa Famida, Salma Akther, Mahbubur Rahman, Leanne Unicomb, Zachary Butzin-Dozier, Caitlin Hemlock, Alan E. Hubbard, Christine P. Stewart, Lia C. H. Fernald, John M. Colford, Stephen P. Luby, Firdaus S. Dhabhar

**Affiliations:** Division of Epidemiology and Biostatistics, School of Public Health, University of California, Berkeley, Berkeley Way West, 2121 Berkeley Way, #5302, Berkeley, CA 94704, USA; Department of Medicine, University of California, San Francisco, California, 1001 Potrero Ave, San Francisco, CA 94110; Infectious Diseases Division, International Centre for Diarrhoeal Disease Research, Bangladesh, GPO Box 128, Dhaka 1000, Bangladesh; Department of Medicine, University of Maryland, Room 7-010, Bressler Research Building, 655 West Baltimore Street, Baltimore, MD 21201, USA; Francis I. Proctor Foundation, University of California, San Francisco, California, 429 Illinois Street, San Francisco, CA 94158, USA; Department of Nutrition, University of California Davis, 3253 Meyer Hall, One Shields Avenue, Davis, CA 95616, USA; Division of Community Health Sciences, School of Public Health, University of California, Berkeley, Berkeley Way West, 2121 Berkeley Way, #5302, Berkeley, CA 94704, USA; Division of Infectious Diseases and Geographic Medicine, Stanford University, Y2E2, MC #4205, 473 Via Ortega, Stanford, CA 94305, USA; Department of Psychiatry and Behavioral Sciences, Department of Microbiology & Immunology, Sylvester Comprehensive Cancer Center, Miller School of Medicine, University of Miami, 1120 NW 14^th^ Street, Miami, FL 33136, USA

## Abstract

While studies have speculated that immune function may play a role in how water, sanitation, hygiene, and nutritional (N+WSH) interventions may individually impact child growth and development, the combined effects of these interventions on immune system development are unknown. Here, we report on a trial in rural Bangladesh, where we cluster-randomized pregnant women into control and N+WSH arms. Among the birth cohort, we quantified plasma IL-1β, IL-6, TNF-α, IL-2, IL-12p70, IFN-γ, IL-4, IL-5, IL-13, IL-17A, IL-21, IL-10, and GM-CSF at ages 14 and 28 months. Cytokine ratios were included as prespecified outcomes to examine the net inflammatory environment. We assessed 704 children. After one year, TNF-α/IL-10, IL-12/IL-10, and IL-17A/IL-10 ratios were lower in the intervention group compared to the control group (mean difference: -0.12 to -0.19, p<0.05), indicating the intervention promoted IL-10 driven immunoregulation. Similar reductions in ratios of pro-inflammatory cytokines to IL-10 were sustained in the intervention group after two years. After one year, IL-12/IL-4, IL-12/IL-5, IFN-γ/IL-5, and IL-12/IL-13 ratios were lower in the intervention group (−0.18 to -0.27, p<0.05), suggesting a shift towards a Th2 cytokine response. These findings suggest that the N+WSH intervention enhanced the immunoprotective and immunoregulatory responses, and suppressed/counteracted the inflammatory/immunopathological response, of the immune triad.

## Introduction

Millions of children in low-income countries are at increased risk of growth impairment, neurodevelopmental delay, and early mortality^1^. Postnatal inflammation may occupy a key position in multiple causal pathways related to these outcomes^2,3^. Exposure to stress, chronic infections, allergens, and toxins could contribute to early life inflammation.

Infants are typically exposed to antigens through infections, the microbiome, and vaccinations that prime and stimulate the immune system^4^. Antigens activate naïve T-cells to differentiate into Th1, Th2, and Th17 subsets. Antigenic challenge from intracellular pathogens (e.g., viruses) typically elicit a Th1 profile of cytokines, including interferon(IFN)-γ and interleukin(IL)-12, while extracellular pathogens (e.g., helminths) and allergens induce a Th2 response involving IL-4, IL-5, and IL-13. Maintaining the delicate balance between the Th1 and Th2 responses is a complex process, tightly regulated by IL-10^5^. The Th17 response protects against extracellular pathogens (e.g., fungi) through IL-17A and IL-21 production. The interactions between immunoprotective, immunoregulatory, and immunopathologic responses, which form the immune triad during early childhood, may determine the ability of the individual to mount immunoprotective versus immunopathological responses, resist infectious diseases, and affect lifelong health^6^.

The growth hormone/insulin-like growth factor-1 (GH/IGF-1) axis is a key modulator for linear growth^7^. Growth hormone stimulates the secretion of insulin-like growth factor 1 (IGF-1), an important regulator of cell proliferation, anabolic physiology, and immune function^8^. It is widely speculated that an evolutionary adaptation of humans is to utilize pro-inflammatory cytokines to restrict growth and energy storage and redirect energy to ensure survival against pathogens^9^. Pro-inflammatory cytokines (e.g., IL-1β, IL-6, and tumor necrosis factor (TNF)-α) activate production of C-reactive protein (CRP) and alpha-1 acid glycoprotein (AGP) to suppress growth hormone signaling, inhibit IGF-1 expression, and produce growth faltering^2^. Anticytokine and immunomodulation treatments may reverse these negative effects on the GH/IGF-1 axis^7^.

Receptors for inflammatory mediators are concentrated in the hippocampus, the central site of learning, memory, and spatial navigation, and are thought to mediate potential damage caused by neuroinflammation^3,10^. A dysregulated pro-inflammatory response during early life may impair neurogenesis and brain development^3^. Higher concentrations of IL-1β and IL-6 are predictors of poor neurodevelopmental outcomes^10^. Conversely, experimental evidence supports a neurogenesis-promoting role for IL-4, IL-10, and IL-12^11-13^. Investigation of long-term cognitive and behavioral deficits related to early childhood inflammation is in its infancy.

Chronic exposure to nutritional deficiencies and environmental fecal contamination during the first two years of life may contribute to compromised immune system development and increased susceptibility to infection^14^. Evaluating the relationship between modifiable exposures and immune trajectories in early life within the context of randomized controlled trials may help us identify critical periods of vulnerability in disease pathogenesis and immunity that inform targeted interventions for immunologic and other pathways that promote healthy growth and development. To our knowledge, no trials have comprehensively evaluated the effects of nutritional supplements on immune system development in the first two years of life, as most studies thus far have been limited by the small number of immune markers measured. Furthermore, no trials have evaluated the effect of drinking water, sanitation, and handwashing interventions on cell-mediated immunity.

The WASH Benefits trial in rural Bangladesh was designed to assess the impact of drinking water, sanitation, handwashing, and nutritional interventions on child health in the first two years of life^15^. Previously, we reported that the intervention improved linear growth and child development compared to the control group^1,16^. Here, we evaluate whether the drinking water, sanitation, handwashing, and nutritional intervention reduced systemic inflammation among young children.

## Main

### Enrollment

The cluster-randomized WASH Benefits trial was conducted in rural villages in the Gazipur, Mymensingh, Tangail and Kishoreganj districts of Bangladesh^1^. 13279 pregnant women were assessed for eligibility. Between 31 May 2012 and 7 July 2013, 5551 women were enrolled in the study and randomly allocated to one of the intervention or control arms (Fig. 1). This study, which measured circulating cytokines, CRP, AGP, and IGF-1, enrolled a subsample of clusters that was evenly balanced (allocation ratio 1:1) across the control and combined nutrition, drinking water, sanitation, and handwashing intervention (N+WSH) arms. The team visited 996 children at year one, and 1021 children at year two in the control and combined N+WSH arms (Fig. 1). Immune markers were measured in 596 children (60%) at age 14.4 (IQR, 12.7–15.6) months, and 704 children (69%) at age 28.1 (IQR, 26.9–29.5) months (Fig. 1). Household enrollment characteristics were balanced between the intervention and control arms (Table 1) and were similar to those from the overall trial (Supplementary Table 1).

**Table 1.**
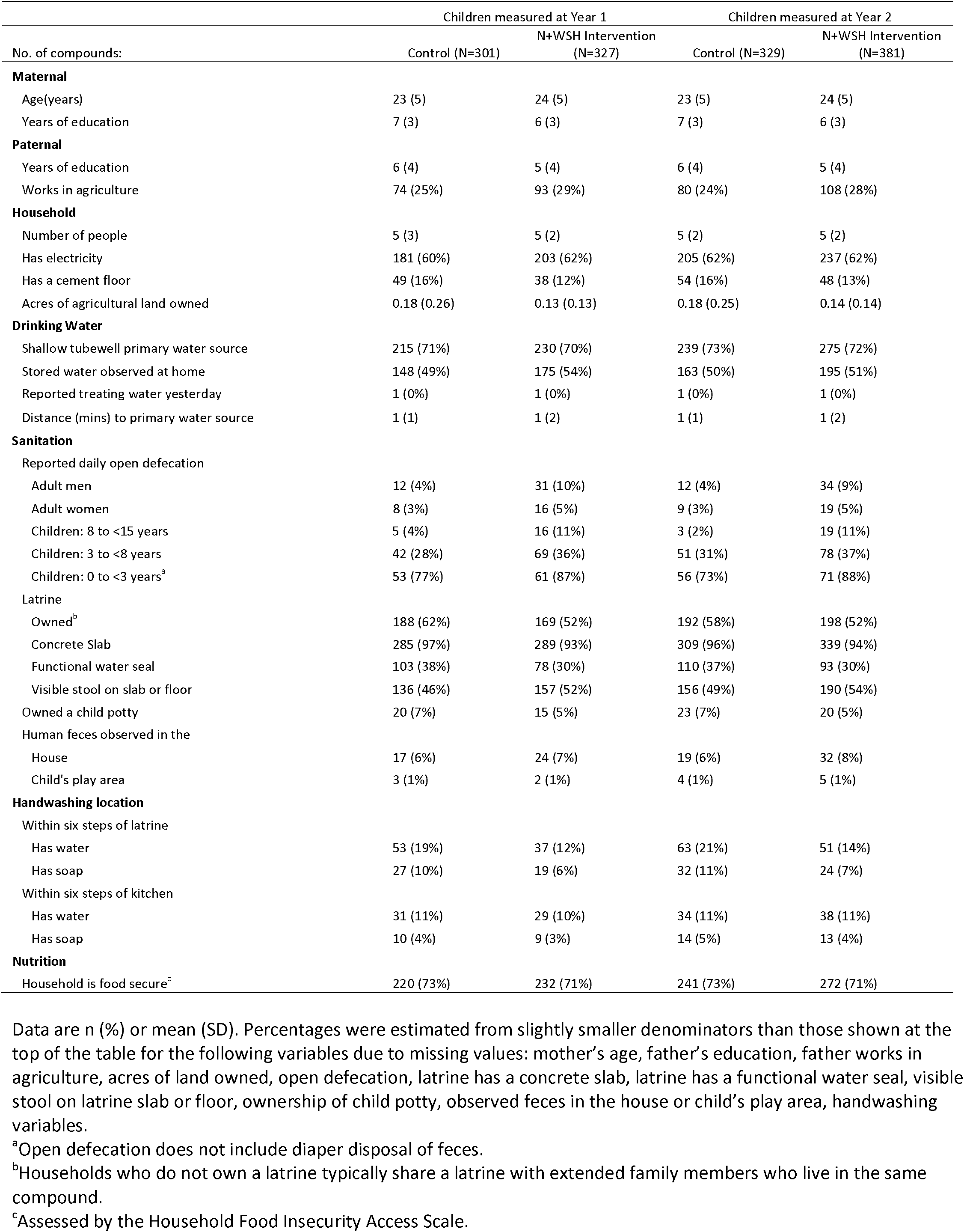
Enrollment characteristics by intervention group

**Figure 1.**
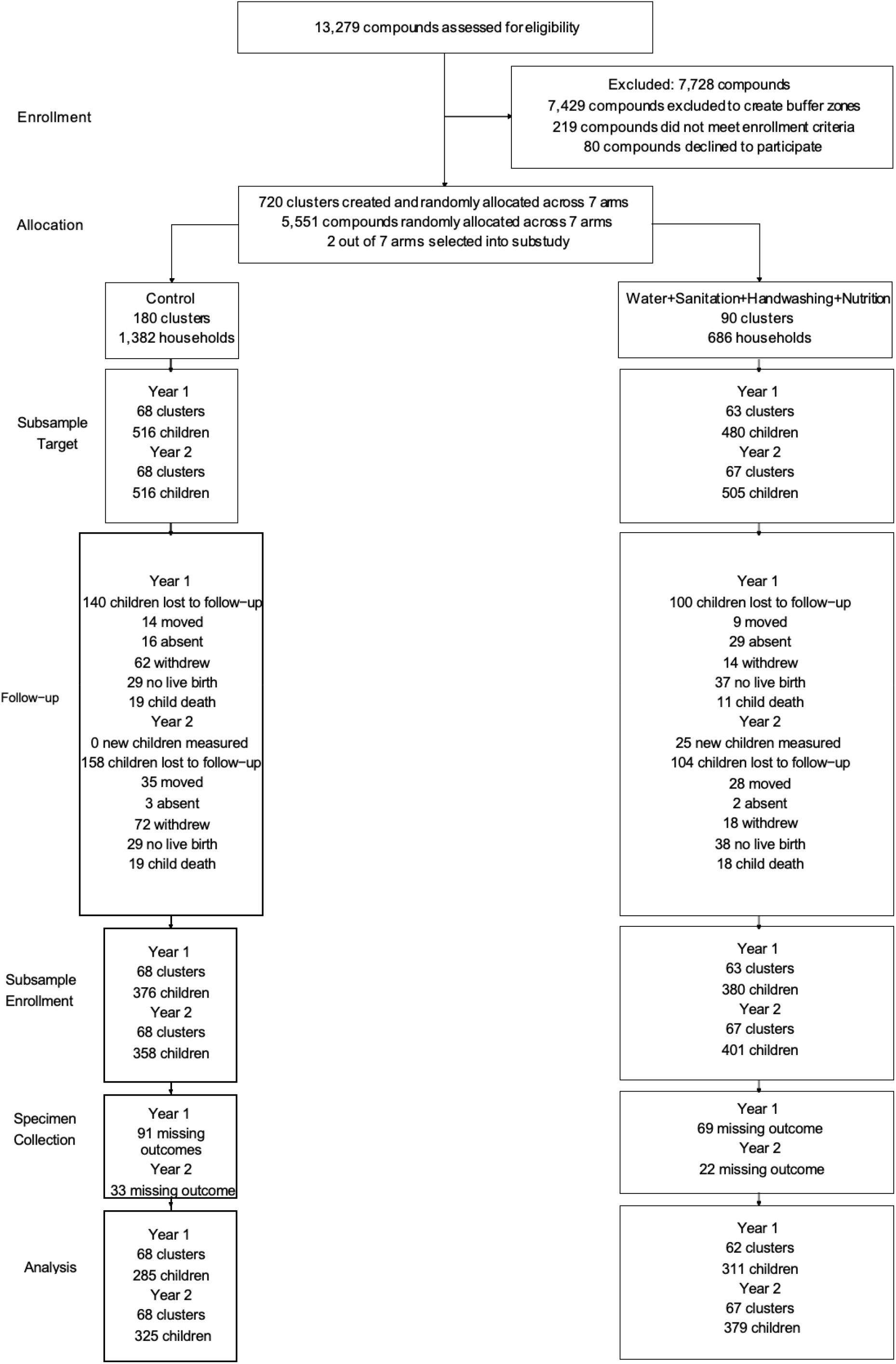
Participant enrollment for the WASH Benefits immune status and growth factor study population.

### Immune status at one year post-intervention

After one year, children in the intervention group had higher circulating concentrations of CRP (+0.37 log mg/ml, CI 0.07, 0.67; adjusted p-value=0.03) and IL-5 (+0.17 log pg/ml, CI 0.05, 0.30; adjusted p-value=0.02) compared to children in the control group (Supplementary Table 2; Supplementary Figs. 1 and 2). To examine the combined effects of pro- and immunoregulatory / anti-inflammatory cytokines, we evaluated ratios of individual pro-inflammatory cytokines to IL-10, a potent immunoregulatory / anti-inflammatory cytokine. After one year, compared to the control arm, the intervention arm had lower TNF-α/IL-10, IL-12/IL-10, and IL-17A/IL-10 ratios (−0.12 to -0.19, adjusted p-values <0.05; Table 2 and Supplementary Table 3; Fig. 2 and Supplementary Fig. 3), due to a non-significant elevation of IL-10 in the intervention group. Overall, the intervention arm exhibited a lower combined Th1/IL-10 ratio compared to controls (−0.13, CI -0.23, -0.03; adjusted p-value=0.02; Table 2 and Supplementary Table 3; Fig. 2 and Supplementary Fig. 3), suggesting that the cytokine balance was tilted toward an immuno-regulated / anti-inflammation state.

**Table 2.**
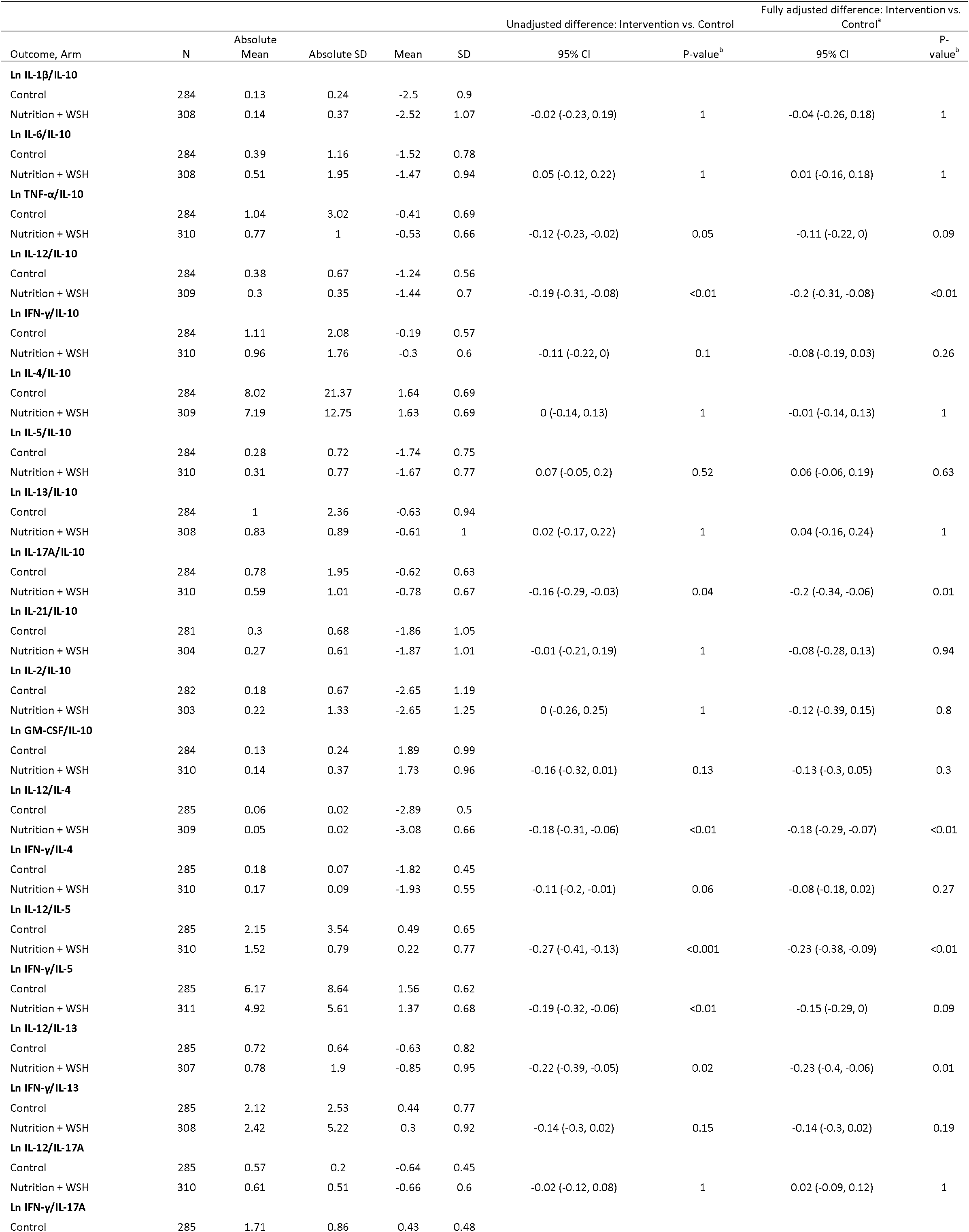
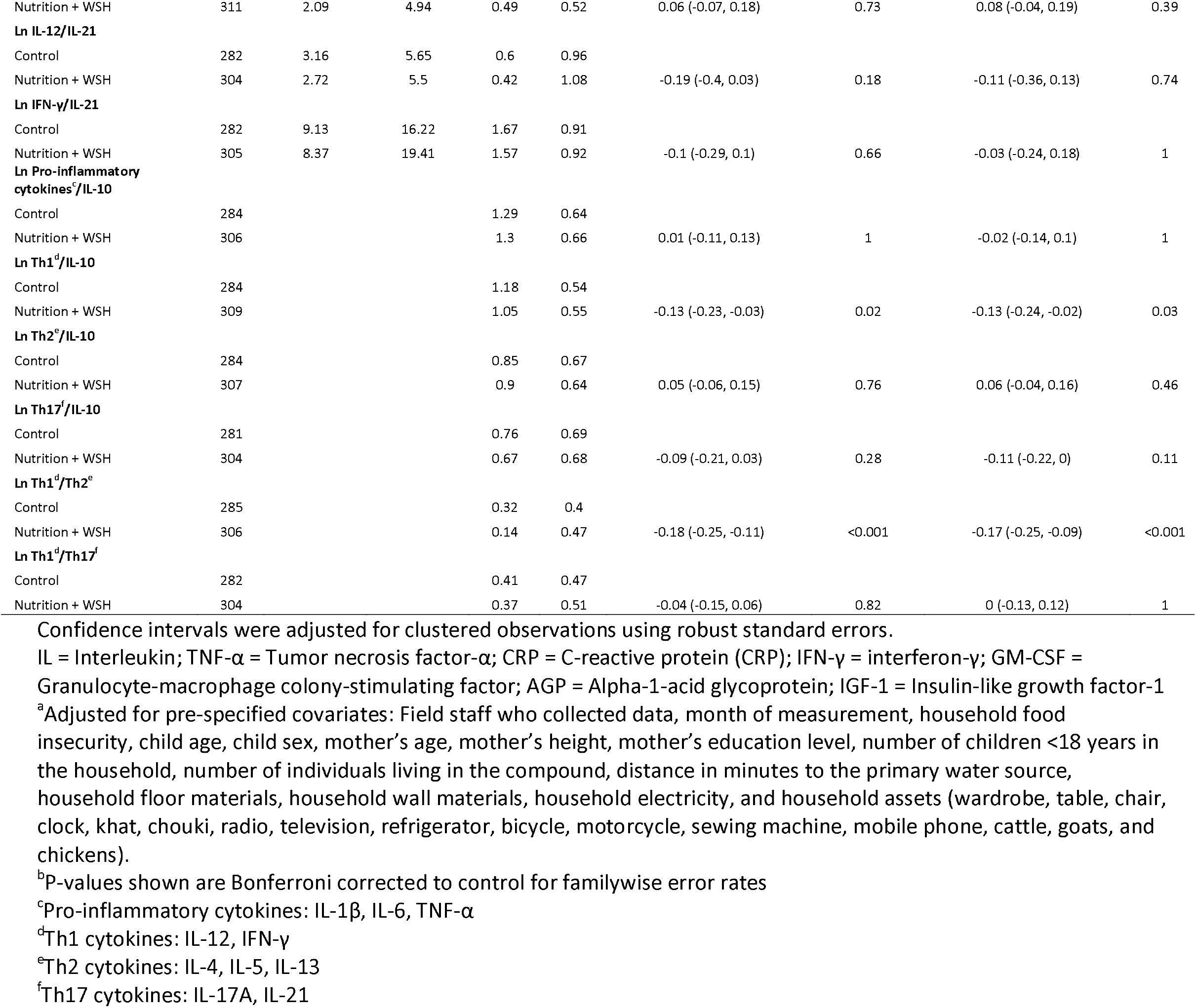
Effect of intervention on cytokine ratios at age 14 months

**Table 3.**
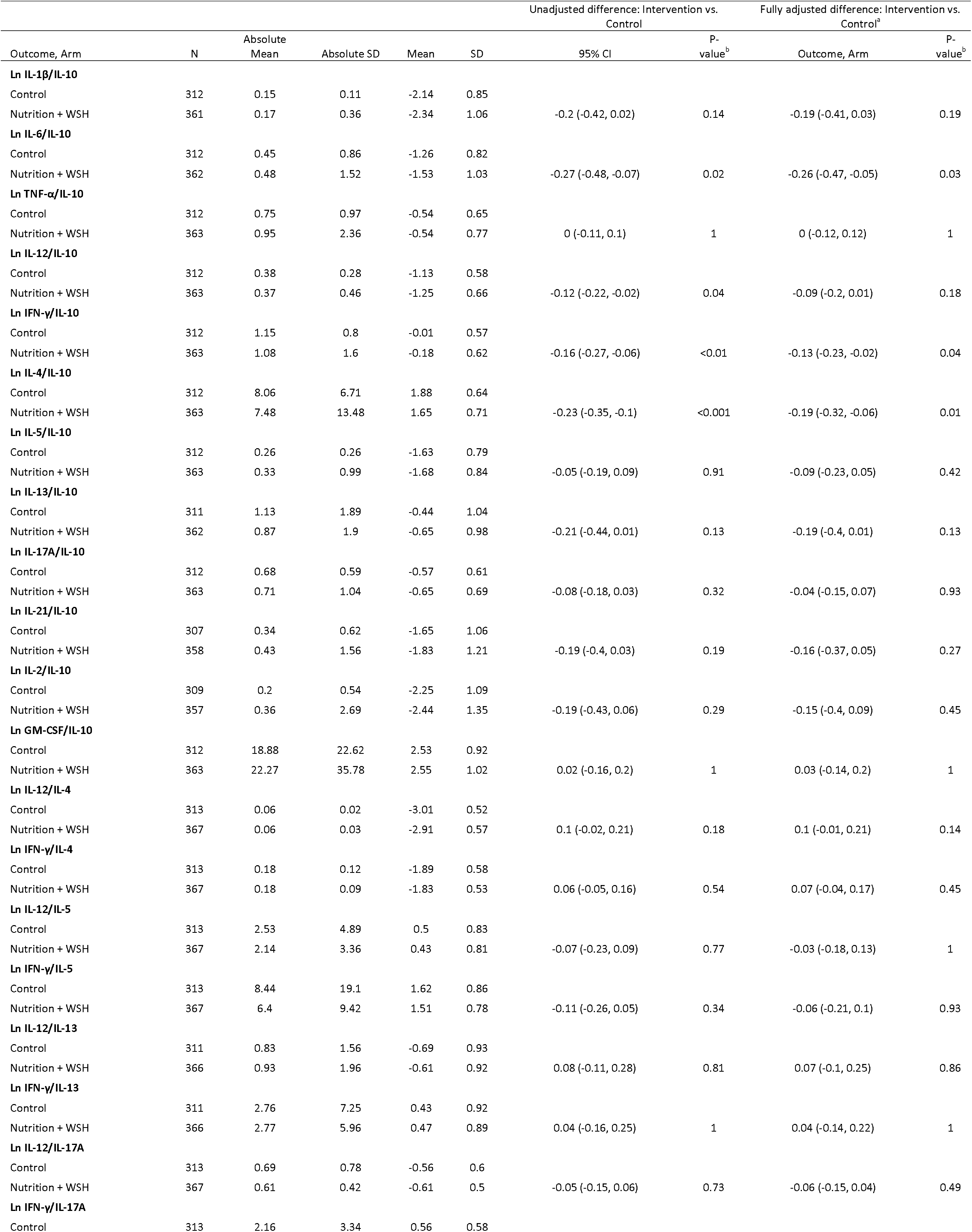
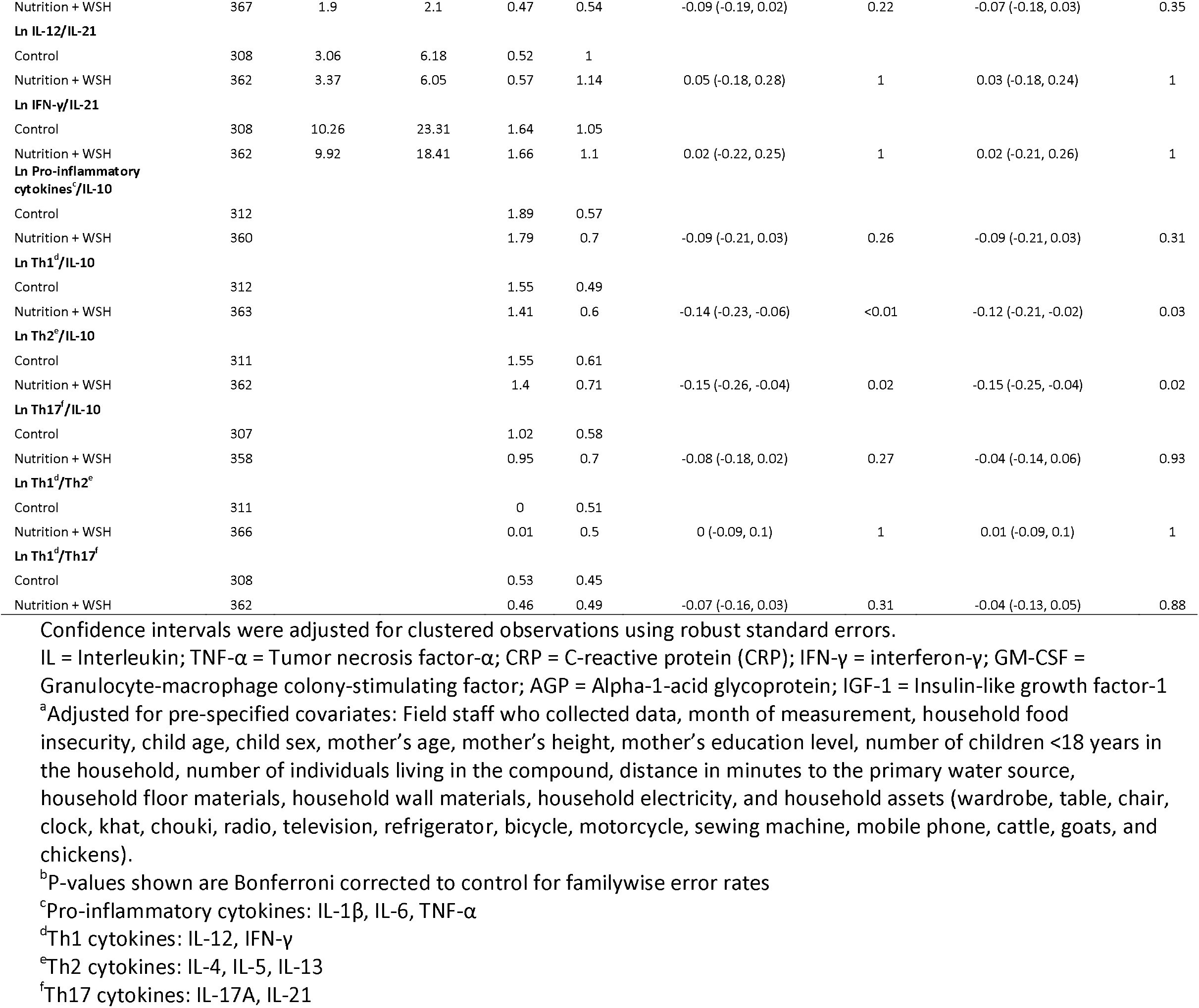
Effect of intervention on cytokine ratios at age 28 months

**Figure 2.**
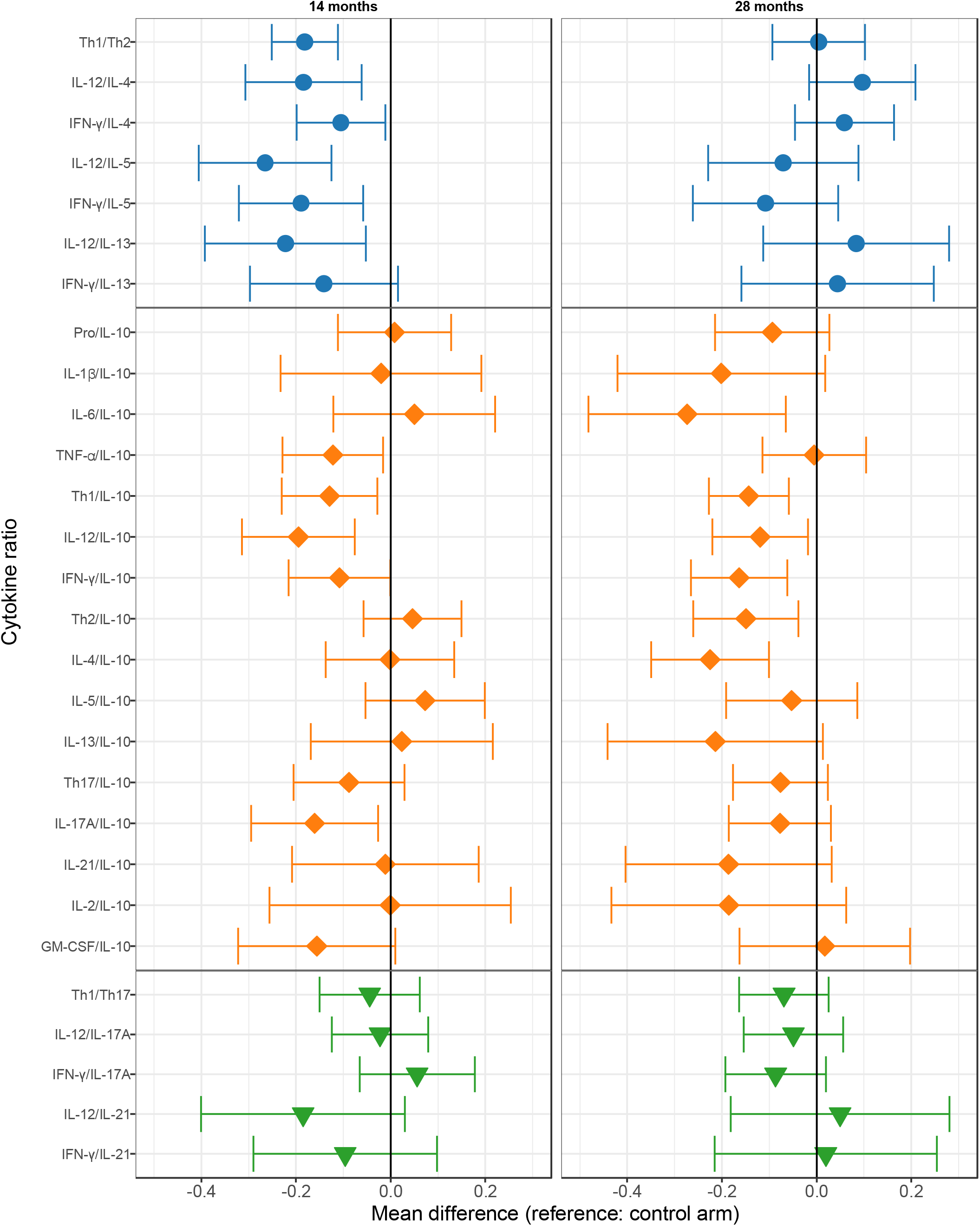
Differences between the control arm and the N+WSH arm for cytokine ratios at ages 14 and 28 months. Pro-inflammatory cytokines (IL-1β, IL-6, TNF-α); Th1 cytokines (IL-12, IFN-γ); Th2 cytokines (IL-4, IL-5, IL-13); Th17 cytokines (IL-17A, IL-21)

We assessed individual Th1-type to Th2-type cytokine ratios. After one year, compared to the control arm, the IL-12/IL-4, IL-12/IL-5, IFN-γ/IL-5, and IL-12/IL-13 ratios were lower in the intervention arm (−0.18 to -0.27, adjusted p-values <0.05; Table 2 and Supplementary Table 3; Fig. 2 and Supplementary Fig. 3). Overall, children in the intervention arm had a lower combined Th1/Th2 ratio compared to children in the control arm (−0.18, CI -0.25, -0.11; adjusted p-value <0.001). Together, these year one findings suggest that the intervention resulted in: 1) a more regulated immune system (overall decrease in pro-to immunoregulatory (IL-10) cytokine ratio) and 2) a shift towards a Th2 cytokine response (decreases in Th1 / Th2 cytokine ratios).

### Immune status at two years post-intervention

At two years, differences in circulating individual cytokine concentrations between the intervention and control arm were not observed (Supplementary Table 4; Supplementary Figs. 1 and 2). The reductions in individual ratios of pro-inflammatory cytokines to IL-10 in the intervention arm were sustained at two years: the IL-12/IL-10, IFN-γ/IL-10, and IL-6/IL-10 ratios were lower (−0.12 to -0.27, adjusted p-values <0.05; Table 3 and Supplementary Table 5; Fig. 2 and Supplementary Fig. 3). The intervention shifted the overall cytokine balance towards an immunoregulatory / anti-inflammatory response. The IL-6/IL-10 reduction at two years was due to the intervention arm having a larger decrease in IL-6/IL-10, compared to the control arm, between years one and two (−0.27, CI -0.51, -0.03; adjusted p-value=0.05; Table 4 and Supplementary Table 6). Furthermore, circulating levels of IL-6 decreased by -0.29 log pg/ml in the intervention group and increased by +0.02 log pg/ml in the control group from years one to two, and the difference between the control and intervention arms in the change in IL-6 from years one to two was -0.31 log pg/ml (CI -0.54, -0.08; adjusted p-value=0.02; Supplementary Table 7; Supplementary Figs. 1 and 2). Overall, IL-10 levels declined from year one to year two in both arms but remained slightly elevated in the intervention arm compared to the control arm (Supplementary Fig. 4). The IL-4/IL-10 ratio was also lower in the intervention arm compared to the control arm (−0.23, CI -0.35, -0.1; adjusted p-value <0.001; Table 3 and Supplementary Table 5; Fig. 2 and Supplementary Fig. 3). Children in the intervention arm at year two exhibited a lower combined Th1/IL-10 ratio (−0.14, CI -0.23, -0.06; adjusted p-value <0.001) and Th2/IL-10 ratio (−0.15, CI -0.26, -0.04; adjusted p-value=0.02; Table 3 and Supplementary Table 5; Fig. 2 and Supplementary Fig. 3), suggesting that an immunoregulatory / anti-inflammatory response continued to predominate in the intervention arm at year two.

**Table 4.**
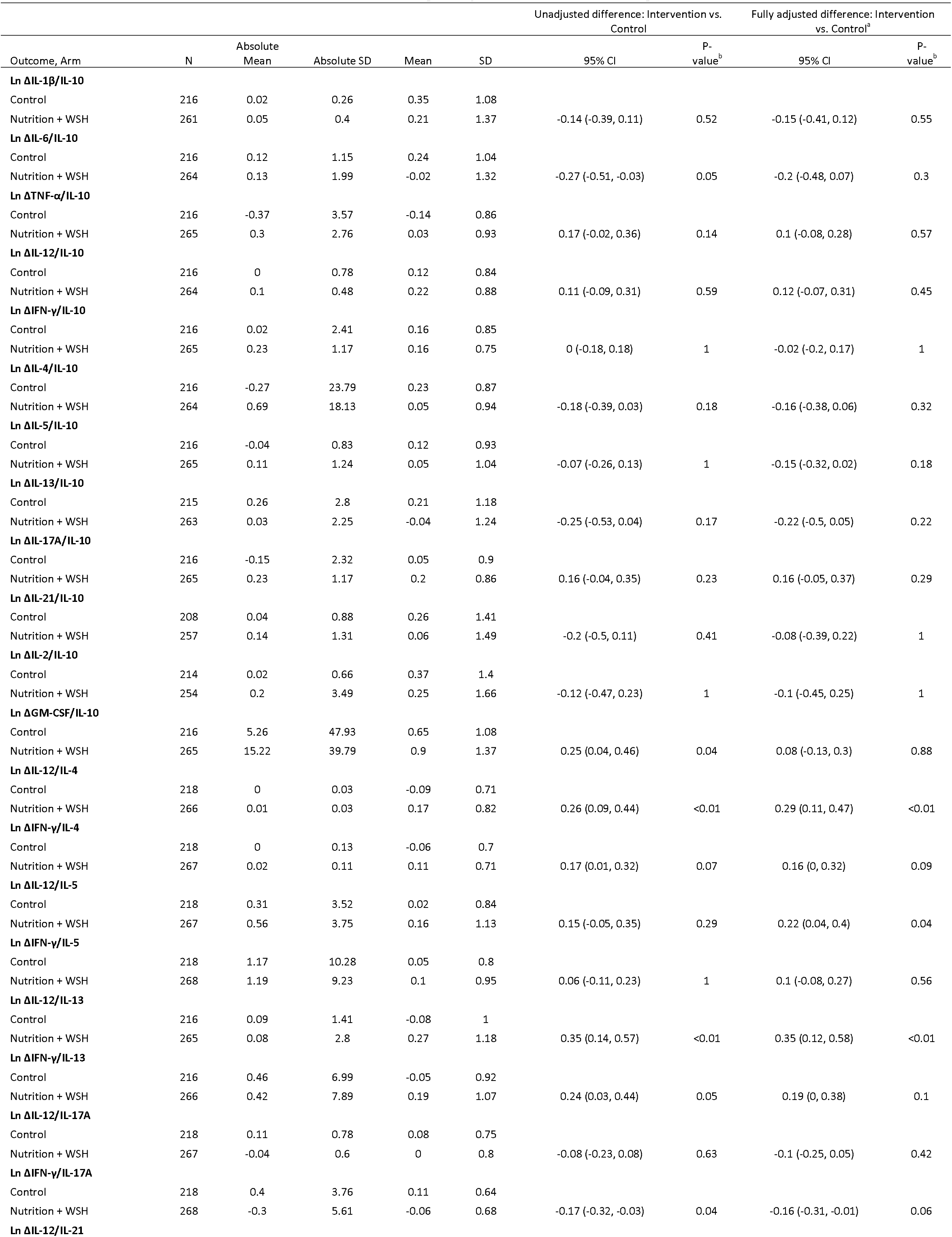
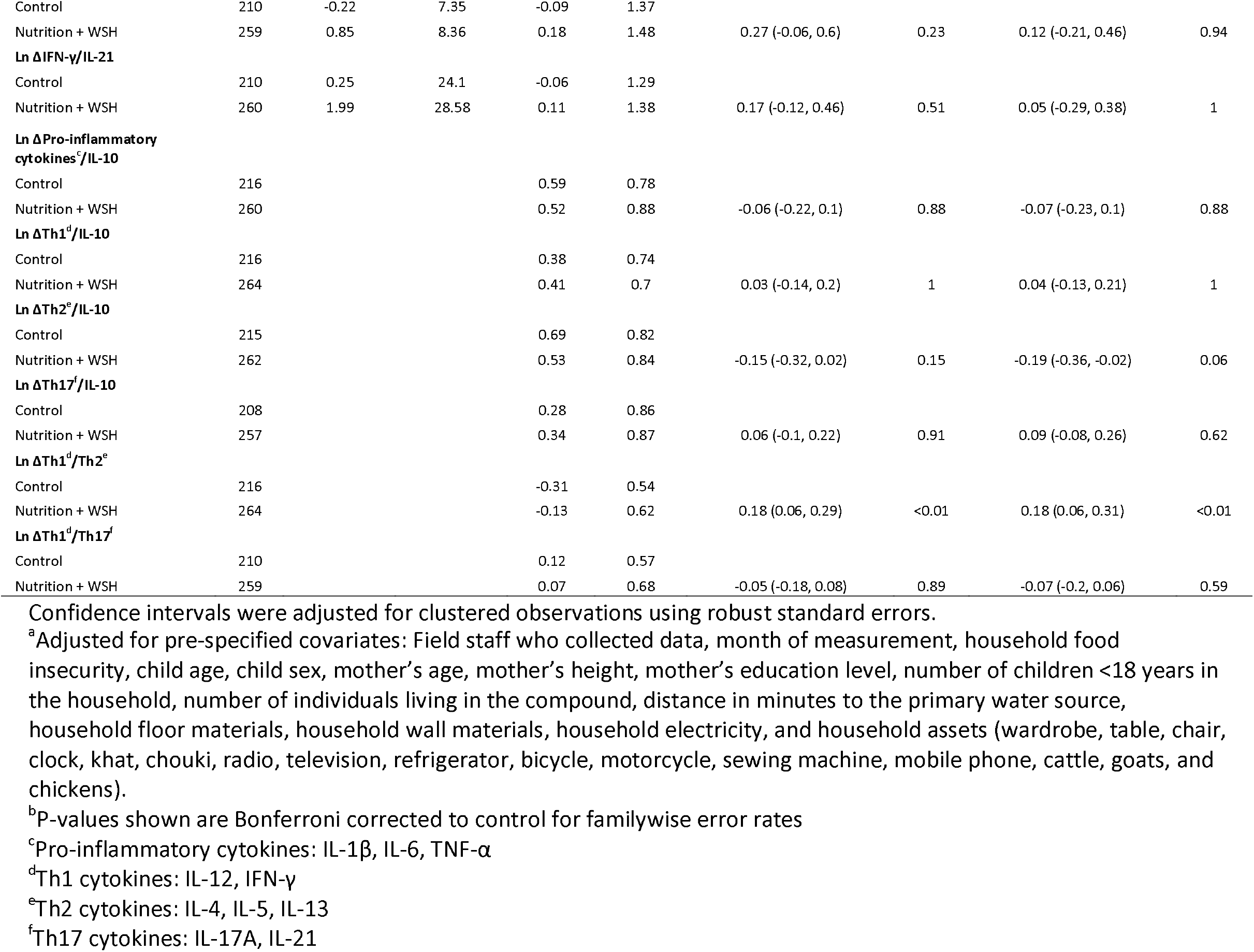
Effect of intervention on change in cytokine ratios between ages 14 and 28 months

### Changes in immune status between years one and two

Household enrollment characteristics were balanced between individuals who had outcome measurements at year one versus those who were lost to follow up at year two (Supplementary Table 1). From years one to two, the intervention arm exhibited a larger increase in GM-CSF/IL-10 (+0.25, CI 0.04, 0.46; adjusted p-value=0.04) and a decrease in IFN-γ/IL-17A (−0.17, CI - 0.32, -0.03; adjusted p-value=0.04) compared to the control arm. The differences ranged from +0.18 to +0.35 between the control and intervention arms in the change in IL-12/IL-4, IL-12/IL-13, IFN-γ/IL-13, and Th1/Th2 cytokine ratios from years one to two (Table 4 and Supplementary Table 6). Although children in the intervention group experienced a predominantly Th2 response at year one compared to the control group, no differences in the individual or combined Th1 and Th2 cytokine ratios were observed between the control and intervention groups by year two (Table 3 and Supplementary Table 5). These trends were due to an overall decline in Th1 cytokine levels in both arms from years one to two, and an increase in Th2 cytokines in the control arm and a decrease in Th2 cytokines in the intervention arm over the one-year period (Supplementary Fig. 4).

### Inflammation sum score and effect modification

There was no difference between the arms for the sum score of inflammation at ages 14 and 28 months (Supplementary Table 8). For all comparisons, unadjusted, adjusted, and IPCW analyses yielded similar estimates (Supplementary Tables 2-8), suggesting there was no imbalance between arms in measured confounders or differential loss to follow-up. After one year, there was some evidence of effect measure modification with child sex (sex by treatment interaction p-value=0.01): among females, the intervention group had higher concentrations of AGP compared to the control group (+0.12, CI 0, 0.24; p-value=0.1), and there was no effect among males (Supplementary Table 9). Sex was not an effect modifier with regards to individual cytokines or cytokine ratios at year one and individual cytokines at year two (Supplementary Tables 10 and 11). At year two, among females, the intervention group had lower IL-12/IL-5, IL-12/IL-17A, and Th1/Th17 ratios (sex by treatment interaction p-values<0.05) compared to the control group, and there was no effect of intervention on these ratios among males (Supplementary Table 12). The combined Th1/Th2 ratio was lower in the intervention group among females and higher in the intervention group among males compared to the control group (sex by treatment interaction p-value=0.03; Supplementary Table 12).

## Discussion

During the first year of life, the combined drinking water, sanitation, handwashing, and nutrition intervention enhanced IL-10 driven immunoregulation which continued through year two. Th2 cytokine driven immune responses, which are important for elimination of extracellular pathogens were also enhanced in year one, but not in year two, perhaps because successful reduction of extracellular pathogens could facilitate a rebalancing of the Th1 and Th2 cytokine systems. These findings highlight the potential for drinking water, sanitation, handwashing, and nutrition interventions to promote an immunoregulatory / anti-inflammatory response during the first two years of life, a critical period when growth and developmental potential are determined.

As a key component of innate immunity, CRP opsonizes microbes and activates the complement system. A recent study suggests that CRP also plays a direct role in regulating adaptive immunity by preferentially binding to naïve T cells, inhibiting Th1 differentiation, and tilting the Th1/Th2 balance in favor of Th2 differentiation^17^. Our findings after one year of intervention are in line with this paradigm: the intervention elevated CRP and IL-5, a Th2 cytokine, at age 14 months. Furthermore, the intervention reduced the individual Th1-type to Th2-type cytokine ratios, thus, eliciting Th2-dominant responses that protect against extracellular parasites. Consistent with these findings, individual and combined ratios of pro-inflammatory cytokines to IL-10 were lower in the intervention arm, skewing the immune response towards an immunoregulatory / anti-inflammatory profile.

The findings of a predominant immunoregulatory / anti-inflammatory response after two years of intervention (age 28 months) were similar to the findings at one year. At year two, the intervention arm was associated with lower individual ratios of pro-inflammatory cytokines to IL-10 and lower combined ratios of Th1 to IL-10 and Th2 to IL-10, compared to the control arm. As a key regulatory cytokine, IL-10 is known to inhibit the production of IL-4^18^. Consistent with this notion, we report lower levels of the IL-4/IL-10 ratio in the intervention arm after two years of intervention. Activated Th1 cells, natural killer cells, and macrophages serve key roles in pathogen clearance but also cause tissue damage^5^. Although overall IL-10 levels declined from year one to year two in both groups, the persistent upregulation of IL-10 expression relative to pro-inflammatory cytokines in the intervention group compared to the control group would suppress the excessive responses of these immune cells and thereby prevent damage to the host. In environments with high microbial contamination, an early childhood intervention favoring an immunoregulatory / anti-inflammatory and Th2 effector program may provide protection against extracellular pathogens and harmful overstimulation of Th1 immunity and tissue damage.

Widespread microbial contamination and lack of water, sanitation, and hygiene infrastructure, commonly observed in low-income settings, could lead to an increased risk of recurrent respiratory and enteric infections, chronic systemic and localized gut inflammation, and microbiome dysbiosis^19^. Early exposure to microbes is needed to prime the infant immune system. However, excessive levels of microbial contamination would deter the development of a healthy immune system, which requires a delicate balance between pro- and anti-inflammatory responses. Previously, we reported that the WSH intervention reduced *Escherichia coli* concentrations in stored drinking water and food given to young children^20^. After one year, the N+WSH intervention may have strengthened the ability of the children to mount Th2 cytokine driven immune responses, which are important for elimination of extracellular pathogens (e.g., helminths). This higher Th2 cytokine driven immunity at year one may have facilitated the reductions in diarrhea,^1^ *Giardia* and hookworm infections^21,22^, and respiratory illness^23^ observed at year two. Successful reductions in extracellular pathogens may have restored the balance in the Th1 and Th2 cytokine systems at year two.

Nutritional deficiencies contribute to immune dysfunction and altered risk of infection, and chronic inflammation is hypothesized to be a mediator of impaired growth and development. In this trial, the intervention reduced the prevalence of anemia and iron deficiency^24^. Adequate micronutrient intake in early life is necessary for healthy gut microbiota maturity, a key process in gut mucosal immunity and immune system priming^25^. Gut mucosal and systemic immune results were consistent: previously, we reported reductions in fecal neopterin levels in the intervention arm at one year^26^, which corresponds to the concurrent Th2 dominant response that may have suppressed the production of IFN-γ, a Th1 cytokine that induces macrophages to produce neopterin. The amelioration of chronic inflammation early in the life-course could have knock-on health benefits, including reductions in adult infections and metabolic syndrome^25^. As this trial has previously found that the intervention substantially improved linear growth^1^ and child development^16^, we speculate that these interventions may have facilitated suppression of immunopathologic responses and/or enhancement of immunoprotective responses, allowing infants to redirect energy towards growth and brain development. Taken together, these results provide strong evidence to support the causal relationship between the N+WSH intervention and a predominantly immunoregulatory / anti-inflammatory and Th2 cytokine driven response.

Sex differences in immune responses contribute to differential susceptibility to infectious diseases. Studies have reported that females mount a more powerful immune response than males, and males experience higher rates of childhood infectious disease morbidity and mortality compared to females^27^. Infants experience a surge in reproductive hormones, the minipuberty period, during the first six months of life that induces maturation of the sexual organs. Child sex was an effect modifier: among females, the intervention group experienced an elevation in AGP at year one and at year two, a shift towards the Th2 and Th17 cytokine driven responses – critical protection against extracellular pathogens. Differences in levels of sex hormones may account for these differential intervention effects. Testosterone exhibits immunosuppressive effects, and estrogen promotes immunoregulation and enhances the Th2 cytokine response.^27^ Identifying the factors that mediate these sex-based differences in immune response will enhance targeted interventions among pediatric populations.

The trial had several limitations. Although the maternal immune system could affect development of the infant inflammatory axis during pregnancy, we lack immune data at birth. However, we assume the immune system initial setting would be similar for both groups because household characteristics were balanced by randomization at enrollment. Although intervention effects on immune responses were associated with the combined N+WSH intervention, we were unable to disentangle whether the effects were primarily driven by the nutrition intervention, the WSH intervention, or both. Differential loss to follow-up may have led to selection bias, but the IPCW analysis and the balance across the two arms in a large set of measurable characteristics between children with outcomes and those lost to follow-up indicated that this type of bias was unlikely.

## Conclusions

This study demonstrated that an intensive, combined drinking water, sanitation, hygiene, and nutritional intervention promoted the immunoprotective and immunoregulatory responses and suppressed the immunopathological response of the immune triad among infants living in a highly contaminated environment. Future research should explore whether early life inflammation is a key mediator for impaired growth and child development. Our findings motivate the trial and scale-up of integrated interventions targeting physiological balance and optimization within the immune system during early childhood, which may ultimately enhance the potential for an individual’s growth and development.

## Methods

### Ethics

Participants provided written informed consent. Human subjects committees at icddr,b (PR-11063 and PR-14108), the University of California, Berkeley (2011-09-3652 and 2014-07-6561) and Stanford University (25863 and 35583) approved study protocols. A data safety monitoring committee convened by icddr,b oversaw the study.

### Participants

We enrolled pregnant women in their first two trimesters and their in utero children. Households with low iron and arsenic levels in drinking water, plans to live in the study village for the next two years, and absence of major water, sanitation, or nutrition programs were eligible for inclusion. Children meeting at least two of the following moderate to severe dehydration criteria were excluded from the venipuncture: (1) restless, irritable, (2) sunken eyes, (3) drinks eagerly, thirsty, (4) pinched skin returns to normal position slowly. Additional exclusion criteria for the venipuncture included children being listless or unable to perform their normal activities. Two children met the exclusion criteria: one child in the control arm was excluded at year one, and one child in the intervention arm was excluded at year two.

### Randomization

Clusters consisted of eight neighboring households with eligible pregnant women. To prevent spillover between clusters, a one km buffer around each cluster was created. Eight geographically-adjacent clusters formed a block. An investigator at UC Berkeley (B.F.A.) used a random number generator to block randomize clusters to the non-intervention control arm or one of the six intervention arms [water; sanitation; handwashing; water, sanitation, and handwashing (WSH); nutrition; nutrition, water, sanitation, and handwashing (N+WSH)]. This study assessed children only in the control and the N+WSH arm.

Participants and outcome assessors were not masked because interventions delivered had visible hardware (e.g., latrines, potties, etc.). Laboratory investigators were masked to group assignments. Two investigators (A.L., A.N.M.) conducted independent masked statistical analyses following the pre-registered analysis protocol. After replication of all masked analyses, results were unmasked.

### Procedures

The combined N+WSH intervention group was previously described^1^ and received the following interventions: nutrition [lipid-based nutrient supplements that included ≥100% of the recommended daily allowance of 12 vitamins and nine minerals with 9.6 g of fat and 2.6 g of protein daily for children 6–24 months old and age-appropriate maternal and infant nutrition recommendations (pregnancy–24 months)], drinking water treatment (chlorine tablets and safe water storage vessel), sanitation (child potties, sani-scoop hoes for feces disposal, and a double pit latrine with a water seal), and handwashing (stations with soapy water near the latrine and kitchen). Community health promoters visited households in the intervention arms at least once per week during the first six months, and subsequently, at least once every two weeks to promote recommended behaviors. The control group did not receive interventions or promoter visits. Intervention adherence was high (>80%) for all interventions^1^.

Five ml venous blood samples were collected into Sarstedt S-monovette^**®**^ lithium heparin tubes at one year (age 14 months) and two years (age 28 months) after intervention delivery. Specimens were transported on ice to the laboratory and immediately centrifuged. Plasma was stored at -80°C.

### Outcomes

The pre-specified outcomes were plasma cytokine concentrations (IL-1β, IL-6, TNF-α, IL-2, IL-12p70, IFN-γ, IL-4, IL-5, IL-13, IL-17A, IL-21, IL-10, and GM-CSF), CRP, AGP, and IGF-1. Outcomes were measured at one year and two years after intervention delivery. Pre-specified outcomes included individual and combined cytokine ratios (full list available at https://osf.io/p9c6q/). To examine the combined effects of proinflammatory, Th1, Th2, and Th17 cytokines, the combinations included: pro-inflammatory cytokines (IL-1β + IL-6 + TNF-α), Th1 (IL-12 + IFN-γ), Th2 (IL-4 + IL-5 + IL-13), and Th17 (IL-17A + IL-21). We created a sum score of all cytokines to represent total inflammation. We log transformed all 13 cytokines and Z-scored each. We used k-nearest neighbor imputation for all missing values. We then summed all Z-scores by year and Z-scored the sum to represent total child inflammation at Year 1 and Year 2.

### Plasma biomarker measurement

Plasma IGF-1 (at one and two years), CRP (at one year), and AGP (at one year) were measured at icddr,b following ELISA kit protocols. The initial dilutions were 1:100 for CRP, 1:10000 for AGP, 1:100 for IGF-1 (R&D Systems, Minneapolis, MN). Out-of-range specimens were rerun at higher or lower dilutions. The coefficient of variation for ELISA outcomes was <5%. CRP and AGP at two years were measured using ELISA methods as previously described with a coefficient of variation of <10% (VitMin Lab)^24^.

The 13 plasma cytokines were measured at the University of Maryland using multiplex Luminex technology (Millipore kit HSTCMAG-28SK). Samples were assayed in duplicate. All plates contained high and low controls. A 96 well plate (Greiner) was wet with 200 μl of assay buffer and placed on a shaker for ten minutes. The plate was then decanted and 25 μl of assay buffer and 25 μl of sample, controls, or standards were added to each well. 25 μl of a mixture containing cytokines (1:50 dilution) conjugated to beads was added and the plate was placed on a shaker at 4° overnight.

The plate was then placed on a magnetic washer, 200 μl of wash buffer was added to each well, the plate was set on a shaker at 500 rpm for one minute, and repeated an additional two times. After the last decanting step, 25 μl of detection antibody was added and the plate was placed on a shaker for one hour at room temperature. 25 μl of Phycoerythrin (1:25 dilution) was added to each well, and the plate was placed on the shaker for 30 minutes. The plate was washed three times, and 150 μl of Sheath Fluid was added to each well. The plate was read using a Luminex 100 reader. The data were calculated using Bio-Rad Bio-Plex Software. The coefficient of variation for the cytokines was <20%.

### Statistical analysis

The pre-registered analysis protocol and replication files for the study are available (https://osf.io/p9c6q/). Analyses were conducted using R statistical software version 3.5.3. All biomarker distributions were right-skewed; therefore, we used log transformation. To aid interpretation, non-transformed means are reported alongside the log-transformed means in the tables.

Assuming a two-sided Type-I error of 5%, 65 clusters enrolled per arm (five children measured per cluster), and a range of cluster-level intra-class correlations for repeated measures (0.01 to 0.20), the study had 90% power to detect a difference of 0.23 to 0.34 standard deviations in log immune status biomarkers between the N+WSH arm and the control.

Analyses were intention-to-treat. We compared the N+WSH arm versus the control arm separately at median age 14 months and 28 months.

These analyses generally followed the same methods as described for the trial’s primary outcomes.^1^ We used generalized linear models with robust standard errors that account for repeated measures within clusters and reported two-tailed p-values. Randomization led to balance in observed covariates across arms, so in accordance with our pre-specified analysis plan, the primary analysis was unadjusted. Two sets of secondary adjusted analyses included: (1) adjusting for child age and sex only and (2) fully adjusting for child age, sex, and covariates associated with each outcome (likelihood ratio test p-value<0.20).

We conducted a pre-specified subgroup analysis stratified by sex because biological differences, differential care practices, or other behavioral practices could modify the effect of the combined N+WSH intervention when stratifying by sex.

We compared enrollment characteristics of those with missing specimens versus those with a full set of specimens. As a sensitivity analysis, we repeated the adjusted analyses using inverse probability of censoring weighting (IPCW) to correct for potential bias due to informative censoring, using all live-birth children enrolled in the subsample arms as the full study population^28^. To control familywise error rates, p-values were Bonferroni corrected to account for multiple testing across two visits.

The trial was registered at ClinicalTrials.gov (NCT01590095). icddr,b convened a data and safety monitoring board to oversee the trial.

## Data Availability

Deidentified individual participant data collected for the study and a data dictionary defining each field in the set, will be made available to others. The pre-specified, registered statistical analysis plan and replication files for the study will be available with publication (https://osf.io/p9c6q/).

## Data availability

Deidentified individual participant data collected for the study and a data dictionary defining each field in the set, will be made available to others. The pre-specified, registered statistical analysis plan and replication files for the study will be available with publication (https://osf.io/p9c6q/). The consort checklist for the study is included in Supplementary Information.

## Contributions

A.L. conceived the study and drafted the research protocol and manuscript with input from all listed co-authors; she coordinated input from the study team throughout the project. M.R., L.U., C.P.S. developed the interventions. M.Z.R., L.H., M.R.K., and S.S. performed the laboratory analyses. A.L., S.A., L.U., M.R., A.K.S., M.S.H., P.M., S.L.F., S.A. oversaw study implementation and responded to threats to validity. F.S.D. primarily conceptualized the immune system-related hypotheses and the immune biomarker quantification panel and guided the interpretation of findings. A.L., A.N.M., S.T., L.K., B.F.A., M.R., L.U., Z.B., C.H., A.E.H., C.P.S., L.C.H.F., J.M.C., S.P.L., and F.S.D. developed the analytical approach and interpreted results. A.L., A.N.M., S.T., and Z.B. conducted the statistical analysis and constructed the tables and figures. All authors have reviewed, contributed to, and approved the final version of the manuscript.

## Acknowledgements

We greatly appreciate the families who participated in the study and the dedication of the icddr,b staff who delivered the interventions and collected the data and specimens. This study was funded by Global Development grant OPPGD759 from the Bill & Melinda Gates Foundation to the University of California, Berkeley and by the National Institute of Allergy and Infectious Diseases of the National Institutes of Health [grant number K01AI136885 to A.L.]. The content is solely the responsibility of the authors and does not necessarily represent the official views of the National Institutes of Health. icddr,b is grateful to the Governments of Bangladesh, Canada, Sweden, and the United Kingdom for providing core/unrestricted support.

