## Supplementary Information for "Effects of drinking water, sanitation, handwashing, and nutritional interventions on immune status in young children: a cluster-randomized controlled trial in rural Bangladesh"

**Table of Contents**

**CONSORT Checklist**	2

**Supplementary figures**

Supplementary Fig. 1: Unadjusted differences between the control arm and the N+WSH arm for individual immune

status and growth factor measurements at age 14 and 28 months	4

Supplementary Fig. 2: Adjusted differences between the control arm and the N+WSH arm for individual immune

status and growth factor measurements at age 14 and 28 months	5

Supplementary Fig. 3: Adjusted differences between the control arm and N+WSH arm for cytokine ratios

at age 14 months and 28 months	6

Supplementary Fig. 4: Unadjusted means and 95% confidence intervals by the control arm and N+WSH arms for

individual cytokines and combined cytokine ratios at ages 14 and 28 months	7

**Supplementary tables**

Supplementary Table 1: Enrollment characteristics by intervention group within the WASH Benefits main trial

study population, within the immune status study population enrolled in Year 1, and

within the immune status study population lost to follow-up at Year 2	9

Supplementary Table 2:	Effect of intervention on individual immune status and growth factor measurements at

age 14 months	10

Supplementary Table 3:	Effect of intervention on cytokine ratios at age 14 months	12

Supplementary Table 4:	Effect of intervention on individual immune status and growth factor measurements at

age 28 months	14

Supplementary Table 5:	Effect of intervention on cytokine ratios at age 28 months	16

Supplementary Table 6:	Effect of intervention on change in cytokine ratios between ages 14 and 28 months	18

Supplementary Table 7:	Effect of intervention on change in individual immune status and growth factor measurements

between ages 14 and 28 months	20

Supplementary Table 8:	Effect of intervention on sum score of inflammation at ages 14 months and 28 months	22

Supplementary Table 9:	Effect modification with child sex on individual immune status and growth factor measurements

at age 14 months	23

Supplementary Table 10: Effect modification with child sex on cytokine ratios at age 14 months	24

Supplementary Table 11: Effect modification with child sex on individual immune status and growth factor measurements

at age 28 months	26

Supplementary Table 12: Effect modification with child sex on cytokine ratios at age 28 months	27

**CONSORT Checklist**

| **Item** | **Description** | **Reported in Section** |
| --- | --- | --- |
| **Title and Abstract** | | |
| 1a | Identification as a randomized trial in the title; Identification as a cluster randomized trial in the title | Title |
| 1b | Structured summary of trial design, methods, results, and conclusions | Summary |
| **Introduction** | | |
| Background and Objectives | | |
| 2a | Scientific background and explanation of rationale; Rationale for using a cluster design | Introduction |
| 2b | Specific objectives or hypotheses; Whether objectives pertain to the cluster level, the individual participant level, or both | Summary (Background); Introduction |
| **Methods** | | |
| Trial Design | | |
| 3a | Description of trial design (such as parallel, factorial) including allocation ratio; Definition of cluster and description of how the design features apply to the clusters | Methods |
| 3b | Important changes to methods after trial commencement (such as eligibility criteria), with reasons | N/A |
| Participants | | |
| 4a | Eligibility criteria for participants; Eligibility criteria for clusters | Methods (Participants & Randomization) |
| 4b | Settings and locations where the data were collected | Methods |
| Interventions | | |
| 5 | The interventions for each group with sufficient details to allow replication, including how and when they were actually administered; Whether interventions pertain to the cluster level, the individual participant level, or both | Methods (Procedures) |
| Outcomes | | |
| 6a | Completely defined pre-specified primary and secondary outcome measures, including how and when they were assessed; Whether outcome measures pertain to the cluster level, the individual participant level, or both | Methods (Outcomes) |
| 6b | Any changes to trial outcomes after the trial commenced, with reasons | N/A |
| Sample Size | | |
| 7a | How sample size was determined; Method of calculation, number of cluster(s) (and whether equal or unequal cluster sizes are assumed), cluster size, a coefficient of intracluster correlation (ICC or k), and an indication of its uncertainty | Methods |
| 7b | When applicable, explanation of any interim analyses and stopping guidelines | N/A |
| **Randomization** | | |
| Sequence Generation | | |
| 8a | Method used to generate the random allocation sequence | Methods (Randomization) |
| 8b | Type of randomization; details of any restriction (such as blocking and block size); Details of stratification or matching if used | Methods (Randomization) |
| Allocation Concealment Mechanism | | |
| 9 | Mechanism used to implement the random allocation sequence (such as sequentially numbered containers), describing any steps taken to conceal the sequence until interventions were assigned; Specification that allocation was based on clusters rather than individuals and whether allocation concealment (if any) was at the cluster level, the individual participant level, or both | Methods (Randomization) |
| Implementation | | |
| 10a | Who generated the random allocation sequence, who enrolled clusters, and who assigned clusters to interventions | Methods (Randomization) |
| 10b | Mechanism by which individual participants were included in clusters for the purposes of the trial (such as complete enumeration, random sampling) | Methods (Randomization) |
| 10c | From whom consent was sought (representatives of the cluster, or individual cluster members, or both) and whether consent was sought before or after randomization | Methods (Ethics) |
| Blinding | | |
| 11a | If done, who was blinded after assignment to interventions (for example, participants, care providers, those assessing outcomes)  and how | Methods (Randomization) |
| 11b | If relevant, description of the similarity of interventions | N/A |
| Statistical Methods | | |
| 12a | Statistical methods used to compare groups for primary and secondary outcomes; How clustering was taken into account | Methods (Statistical analysis) |
| 12b | Methods for additional analyses, such as subgroup analyses and adjusted analyses | Methods (Statistical analysis) |
| **Results** | | |
| Participant Flow | | |
| 13a | For each group, the numbers of participants/clusters who were randomly assigned, received intended treatment, and were analyzed for the primary outcome | Results |
| 13b | For each group, losses and exclusions after randomization, together with reasons, for both clusters and individual cluster members | Methods (Participants) |
| Recruitment | | |
| 14a | Dates defining the periods of recruitment and follow-up | N/A |
| 14b | Why the trial ended or was stopped | N/A |
| Baseline Data | | |
| 15 | A table showing baseline demographic and clinical characteristics for each group; Baseline characteristics for the individual and cluster levels as applicable for each group | Table 1; Supplementary Table 1 |
| Numbers Analyzed | | |
| 16 | For each group, number of participants/clusters (denominator) included in each analysis and whether the analysis was by the original assigned groups | Results; Table 1 |
| Outcomes and Estimation | | |
| 17a | For each primary and secondary outcome, results for each group, and the estimated effect size and its precision (such as 95% confidence interval); Results at the individual and cluster levels as applicable and a coefficient of intracluster correlation (ICC or k) for each primary outcome | Results; Tables 2-4; Supplementary Tables 2-8 |
| 17b | For binary outcome, presentation of both absolute and relative effect sizes is recommended | Tables 2-4; Supplementary Tables 2-8 |
| Ancillary Analyses | | |
| 18 | Results of any other analyses performed, including subgroup analyses and adjusted analyses, distinguishing pre-specified from exploratory | Supplementary Tables 2-12 |
| Harms | | |
| 19 | All important harms or unintended effects in each group | N/A |
| **Discussion** | | |
| Limitations | | |
| 20 | Trial limitations, addressing sources of potential bias, imprecision and, if relevant, multiplicity of analyses | Discussion |
| Generalizability | | |
| 21 | Generalizability (external validity, applicability) of the trial findings; Generalizability to clusters and/or individual participants (as relevant) | Discussion |
| Interpretation | | |
| 22 | Interpretation consistent with results, balancing benefits and harms, and considering other relevant evidence | Interpretation; Discussion |
| **Other Information** | | |
| Registration | | |
| 23 | Registration number and name of trial registry | Methods (Statistical analysis) |
| Protocol | | |
| 24 | Where the full trial protocol can be accessed, if available | Methods (Statistical analysis) |
| Funding | | |
| 25 | Sources of funding and other support (such as supply of drugs), role of funders | Funding |

**Supplementary Fig. 1 Unadjusted differences between the control arm and the N+WSH arm for individual immune status and growth factor measurements at age 14 and 28 months**

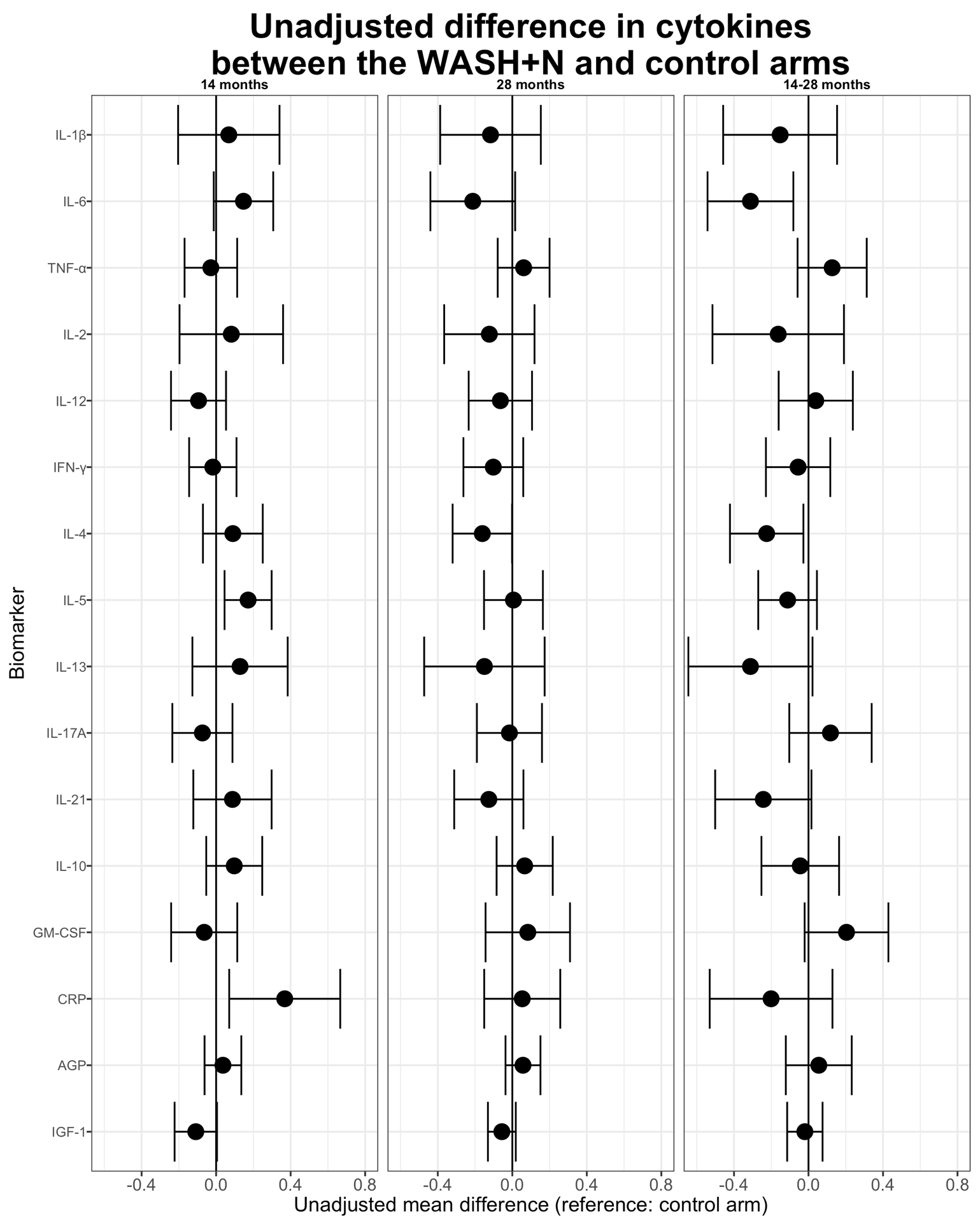

**Supplementary Fig. 2 Adjusted differences between the control arm and the N+WSH arm for individual immune status and growth factor measurements at age 14 and 28 months**

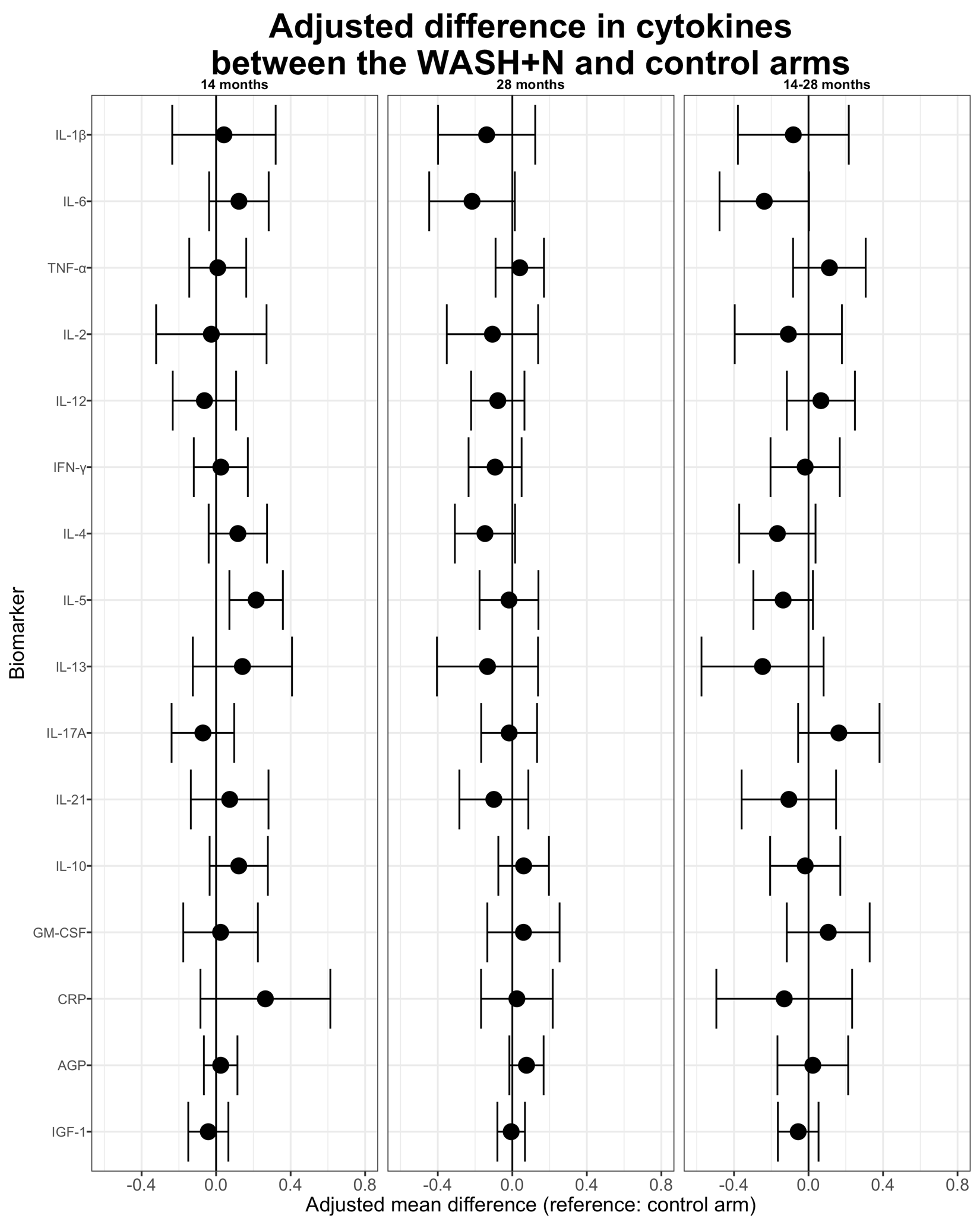

**Supplementary Fig. 3 Adjusted differences between the control arm and N+WSH arm for cytokine ratios at age 14 months and 28 months**

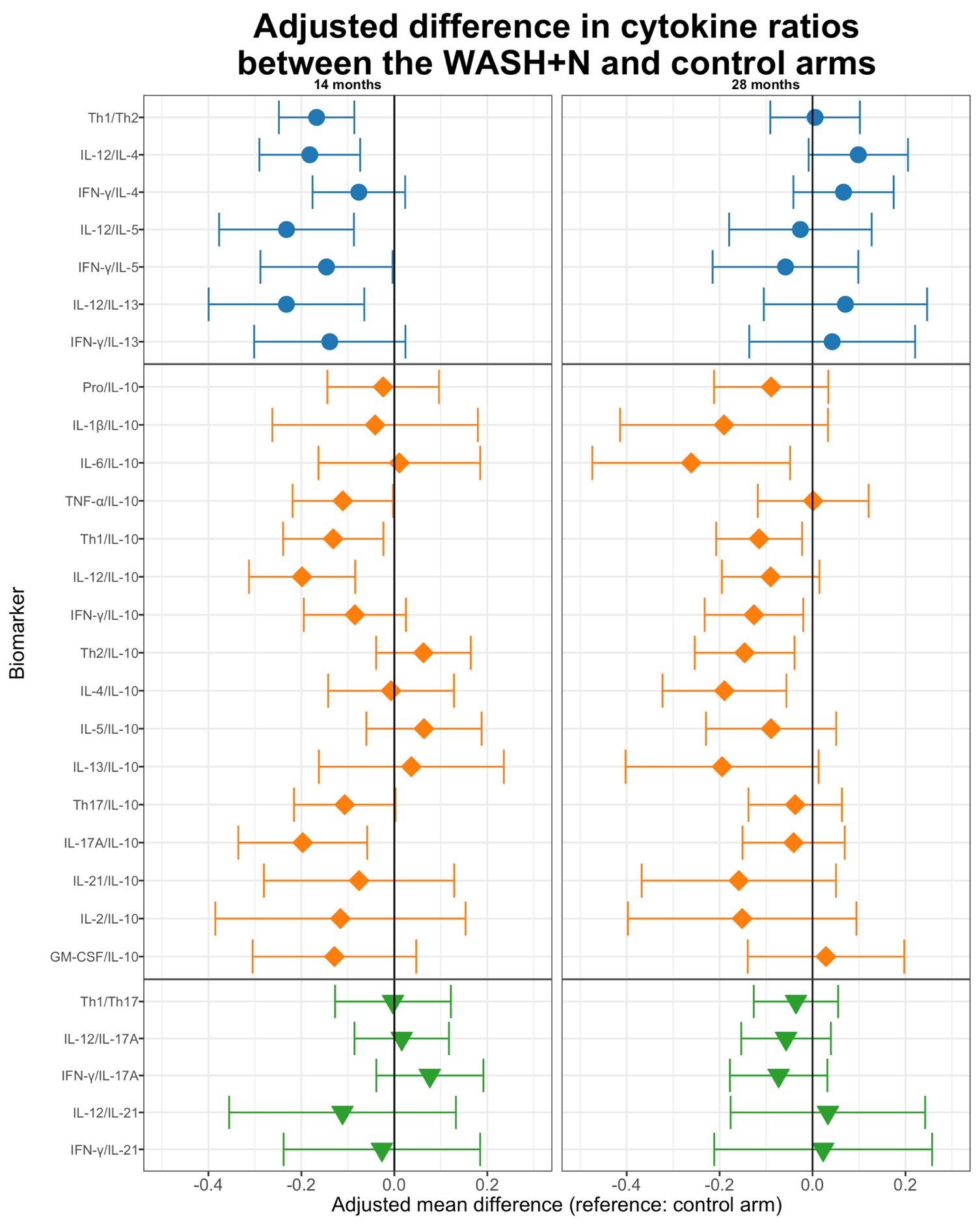

Pro-inflammatory cytokines (IL-1β, IL-6, TNF-α); Th1 cytokines (IL-12, IFN-γ); Th2 cytokines (IL-4, IL-5, IL-13); Th17 cytokines (IL-17A, IL-21)

**Supplementary Fig. 4 Unadjusted means and 95% confidence intervals by the control arm and N+WSH arms for individual cytokines and combined cytokine ratios at ages 14 and 28 months.** The ratio is plotted prior to log-transformation for more comparable values to the constituent biomarkers, but the ratio was log-transformed prior to analysis

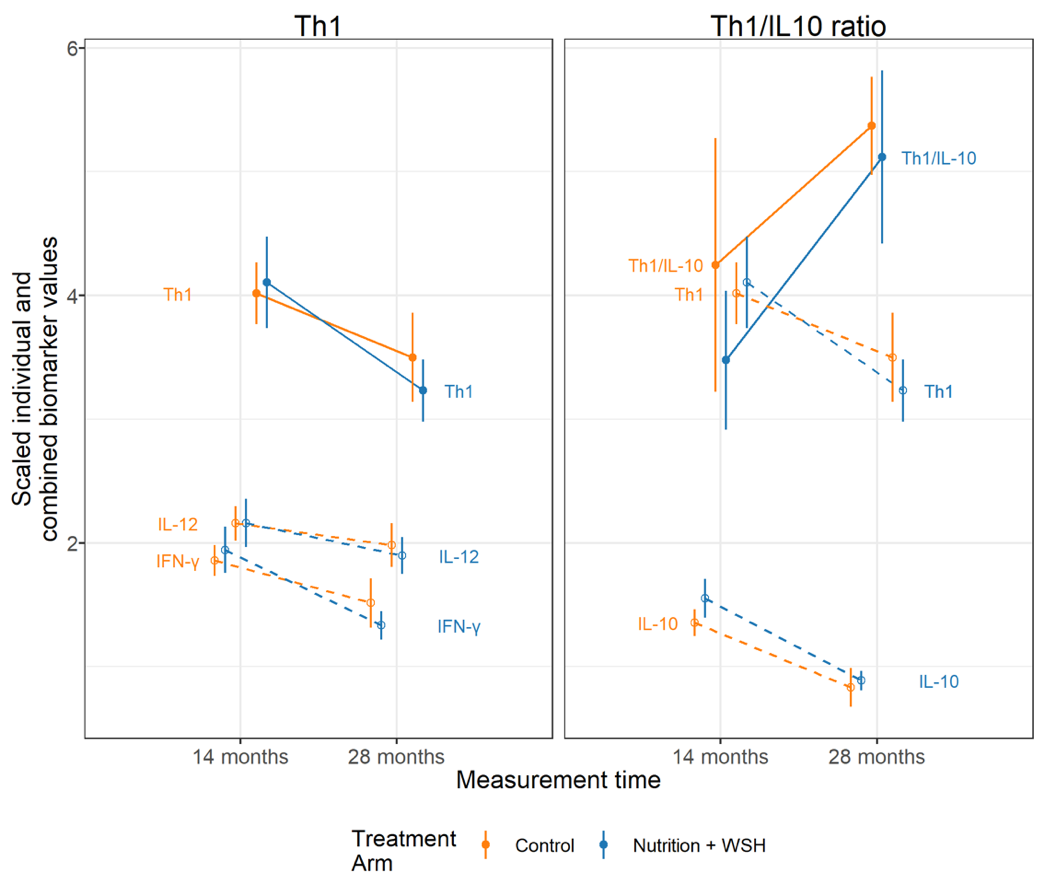
**A.**

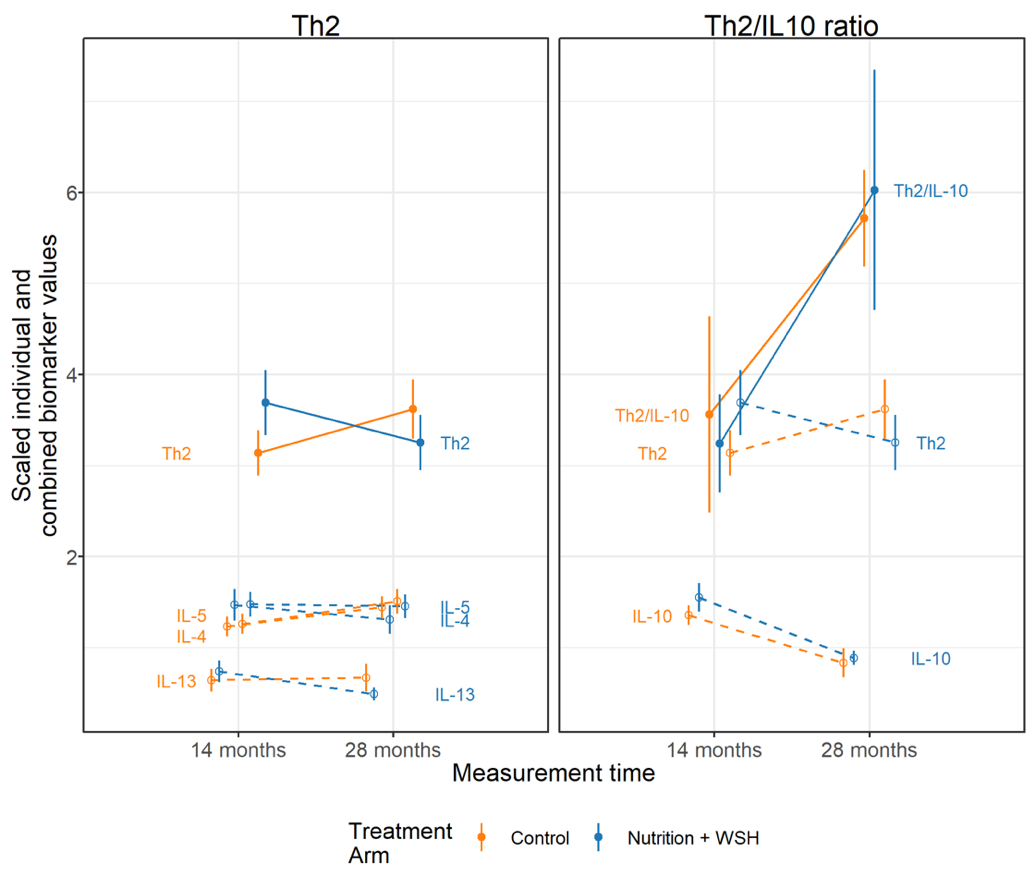

**B.**

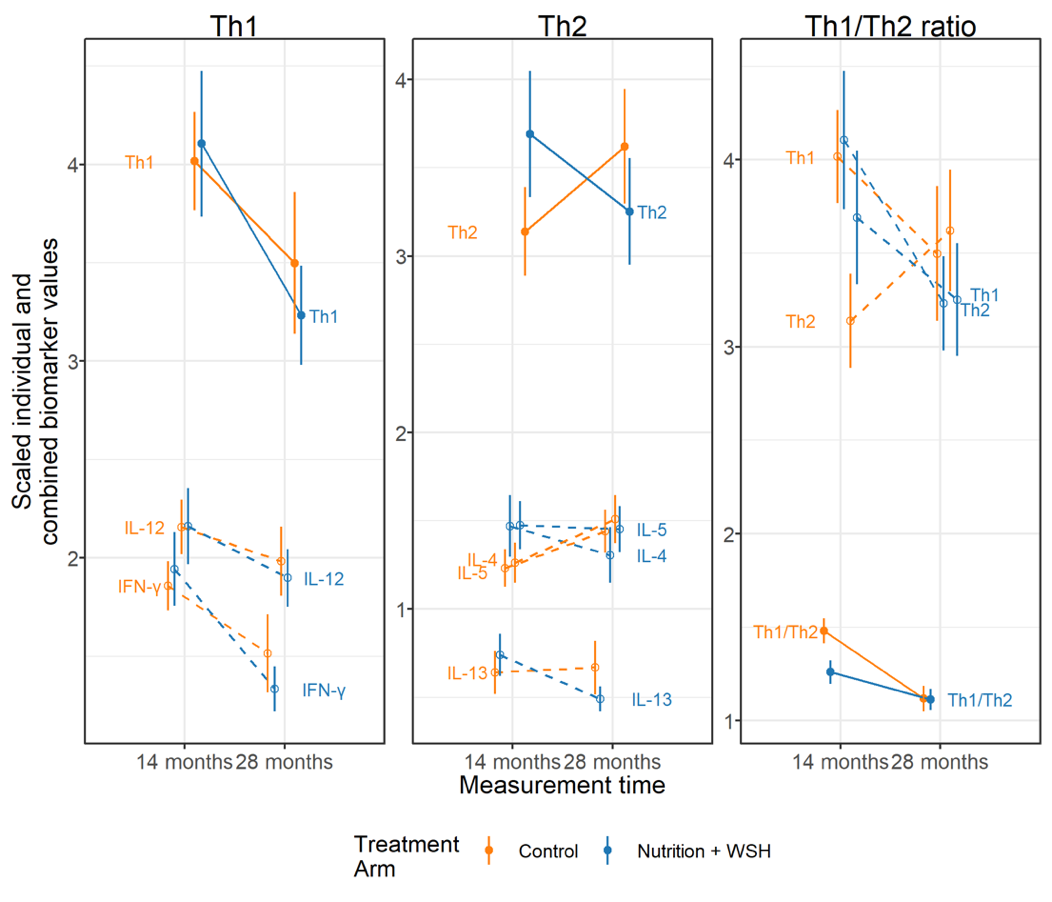

**C.**

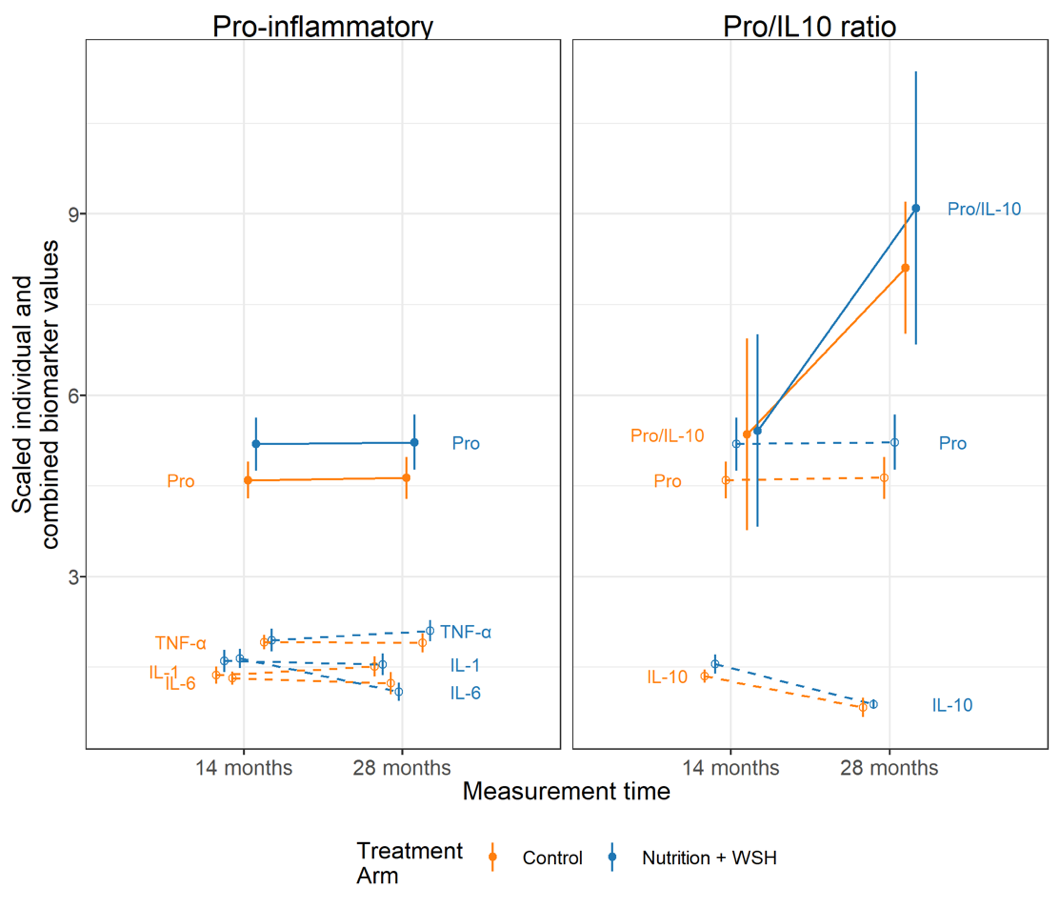
**D.**

Pro-inflammatory cytokines (IL-1β, IL-6, TNF-α); Th1 cytokines (IL-12, IFN-γ); Th2 cytokines (IL-4, IL-5, IL-13); Th17 cytokines (IL-17A, IL-21)

**Supplementary Table 1. Enrollment characteristics by intervention group within the WASH Benefits main trial study population, within the immune status study population enrolled in Year 1, and within the immune status study population lost to follow-up at Year 2**

|  | WASH Benefits Main Trial | | Immune Status Study: Had outcomes at Year 1 | | Immune Status Study: Lost to follow-up at Year 2 | |
| --- | --- | --- | --- | --- | --- | --- |
| No. of compounds: | Control (N=1382) | N + WSH Intervention (N=686) | Control (N=301) | N+WSH Intervention (N=327) | Control (N=56) | N+WSH Intervention (N=39) |
| **Maternal** |  |  |  |  |  |  |
| Age(years) | 24 (5) | 24 (6) | 23 (5) | 24 (5) | 23 (4) | 23 (5) |
| Years of education | 6 (3) | 6 (3) | 7 (3) | 6 (3) | 7 (3) | 5 (3) |
| **Paternal** |  |  |  |  |  |  |
| Years of education | 5 (4) | 5 (4) | 6 (4) | 5 (4) | 6 (4) | 5 (4) |
| Works in agriculture | 414 (30%) | 207 (30%) | 74 (25%) | 93 (29%) | 12 (21%) | 7 (18%) |
| **Household** |  |  |  |  |  |  |
| Number of people | 5 (2) | 5 (2) | 5 (3) | 5 (2) | 5 (3) | 5 (2) |
| Has electricity | 784 (57%) | 412 (60%) | 181 (60%) | 203 (62%) | 31 (55%) | 23 (59%) |
| Has a cement floor | 145 (10%) | 72 (10%) | 49 (16%) | 38 (12%) | 9 (16%) | 4 (10%) |
| Acres of agricultural land owned | 0.15 (0.21) | 0.14 (0.38) | 0.18 (0.26) | 0.13 (0.13) | 0.16 (0.21) | 0.11 (0.11) |
| **Drinking Water** |  |  |  |  |  |  |
| Shallow tubewell primary water source | 1038 (75%) | 504 (73%) | 215 (71%) | 230 (70%) | 43 (77%) | 26 (67%) |
| Stored water observed at home | 666 (48%) | 331 (48%) | 148 (49%) | 175 (54%) | 28 (50%) | 24 (62%) |
| Reported treating water yesterday | 4 (0%) | 2 (0%) | 1 (0%) | 1 (0%) | 0 (0%) | 0 (0%) |
| Distance (mins) to primary water source | 1 (3) | 1 (2) | 1 (1) | 1 (2) | 1 (1) | 1 (1) |
| **Sanitation** |  |  |  |  |  |  |
| Reported daily open defecation |  |  |  |  |  |  |
| Adult men | 97 (7%) | 50 (7%) | 12 (4%) | 31 (10%) | 2 (4%) | 3 (8%) |
| Adult women | 62 (4%) | 24 (4%) | 8 (3%) | 16 (5%) | 1 (2%) | 1 (3%) |
| Children: 8 to <15 years | 53 (10%) | 28 (10%) | 5 (4%) | 16 (11%) | 2 (9%) | 2 (17%) |
| Children: 3 to <8 years | 267 (38%) | 134 (37%) | 42 (28%) | 69 (36%) | 7 (28%) | 8 (36%) |
| Children: 0 to <3 years^a^ | 245 (82%) | 123 (88%) | 53 (77%) | 61 (87%) | 7 (88%) | 3 (75%) |
| Latrine |  |  |  |  |  |  |
| Owned^b^ | 750 (54%) | 367 (53%) | 188 (62%) | 169 (52%) | 39 (70%) | 21 (54%) |
| Concrete Slab | 1251 (95%) | 621 (94%) | 285 (97%) | 289 (93%) | 56 (100%) | 37 (97%) |
| Functional water seal | 358 (31%) | 155 (27%) | 103 (38%) | 78 (30%) | 25 (45%) | 12 (38%) |
| Visible stool on slab or floor | 625 (48%) | 298 (46%) | 136 (46%) | 157 (52%) | 21 (38%) | 18 (47%) |
| Owned a child potty | 61 (4%) | 30 (4%) | 20 (7%) | 15 (5%) | 6 (11%) | 0 (0%) |
| Human feces observed in the |  |  |  |  |  |  |
| House | 114 (8%) | 49 (8%) | 17 (6%) | 24 (7%) | 5 (9%) | 2 (5%) |
| Child's play area | 21 (2%) | 7 (1%) | 3 (1%) | 2 (1%) | 0 (0%) | 0 (0%) |
| **Handwashing location** |  |  |  |  |  |  |
| Within six steps of latrine |  |  |  |  |  |  |
| Has water | 178 (14%) | 72 (11%) | 53 (19%) | 37 (12%) | 11 (22%) | 2 (6%) |
| Has soap | 88 (7%) | 36 (6%) | 27 (10%) | 19 (6%) | 7 (14%) | 2 (6%) |
| Within six steps of kitchen |  |  |  |  |  |  |
| Has water | 118 (9%) | 60 (9%) | 31 (11%) | 29 (10%) | 5 (10%) | 2 (6%) |
| Has soap | 33 (3%) | 18 (3%) | 10 (4%) | 9 (3%) | 0 (0%) | 0 (0%) |
| **Nutrition** |  |  |  |  |  |  |
| Household is food secure^c^ | 932 (67%) | 485 (71%) | 220 (73%) | 232 (71%) | 42 (75%) | 28 (72%) |

Data are n (%) or mean (SD). Percentages were estimated from slightly smaller denominators than those shown at the top of the table for the following variables due to missing values: mother’s age, father’s education, father works in agriculture, acres of land owned, open defecation, latrine has a concrete slab, latrine has a functional water seal, visible stool on latrine slab or floor, ownership of child potty, observed feces in the house or child’s play area, handwashing variables.

^a^Open defecation does not include diaper disposal of feces.

^b^Households who do not own a latrine typically share a latrine with extended family members who live in the same compound.

^c^Assessed by the Household Food Insecurity Access Scale.

**Supplementary Table 2. Effect of intervention on individual immune status and growth factor measurements at age 14 months**

|  |  |  |  |  |  | Unadjusted difference: Intervention vs. Control | | Age- and sex- adjusted difference: Intervention vs. Control | | Fully adjusted difference: Intervention vs. Control^a^ | | IPCW adjusted difference: Intervention vs. Control^b^ | |
| --- | --- | --- | --- | --- | --- | --- | --- | --- | --- | --- | --- | --- | --- |
| Outcome, Arm | N | Absolute Mean | Absolute SD | Mean | SD | 95% CI | P-value^c^ | 95% CI | P-value^c^ | 95% CI | P-value^c^ | 95% CI | P-value^c^ |
| **Ln IL-1β (pg/ml)** |  |  |  |  |  |  |  |  |  |  |  |  |  |
| Control | 285 | 1.07 | 0.68 | -0.26 | 1.01 |  |  |  |  |  |  |  |  |
| Nutrition + WSH | 309 | 1.25 | 0.85 | -0.19 | 1.15 | 0.07 (-0.2, 0.34) | 1 | 0.07 (-0.21, 0.34) | 1 | 0.04 (-0.24, 0.32) | 1 | 0.05 (-0.23, 0.33) | 1 |
| **Ln IL-6 (pg/ml)** |  |  |  |  |  |  |  |  |  |  |  |  |  |
| Control | 285 | 2.49 | 1.51 | 0.72 | 0.67 |  |  |  |  |  |  |  |  |
| Nutrition + WSH | 309 | 3.11 | 2.15 | 0.86 | 0.82 | 0.15 (-0.01, 0.31) | 0.14 | 0.12 (-0.04, 0.28) | 0.29 | 0.12 (-0.04, 0.28) | 0.27 | 0.11 (-0.06, 0.28) | 0.39 |
| **Ln TNF-α (pg/ml)** |  |  |  |  |  |  |  |  |  |  |  |  |  |
| Control | 285 | 6.99 | 3.33 | 1.83 | 0.5 |  |  |  |  |  |  |  |  |
| Nutrition + WSH | 311 | 7.14 | 3.94 | 1.8 | 0.61 | -0.03 (-0.17, 0.11) | 1 | -0.02 (-0.16, 0.12) | 1 | 0.01 (-0.14, 0.16) | 1 | -0.07 (-0.23, 0.08) | 0.69 |
| **Ln CRP (mg/L)** |  |  |  |  |  |  |  |  |  |  |  |  |  |
| Control | 253 | 4.43 | 16.1 | -0.1 | 1.63 |  |  |  |  |  |  |  |  |
| Nutrition + WSH | 290 | 4.35 | 9.87 | 0.27 | 1.57 | 0.37 (0.07, 0.67) | 0.03 | 0.32 (0.04, 0.61) | 0.05 | 0.26 (-0.08, 0.61) | 0.27 | 0.22 (-0.05, 0.5) | 0.22 |
| **Ln IL-12 (pg/ml)** |  |  |  |  |  |  |  |  |  |  |  |  |  |
| Control | 285 | 2.95 | 1.23 | 0.98 | 0.52 |  |  |  |  |  |  |  |  |
| Nutrition + WSH | 310 | 2.95 | 1.48 | 0.88 | 0.8 | -0.09 (-0.24, 0.05) | 0.42 | -0.1 (-0.25, 0.05) | 0.41 | -0.06 (-0.23, 0.11) | 0.94 | -0.09 (-0.24, 0.05) | 0.44 |
| **Ln IFN-γ (pg/ml)** |  |  |  |  |  |  |  |  |  |  |  |  |  |
| Control | 285 | 8.53 | 3.99 | 2.05 | 0.45 |  |  |  |  |  |  |  |  |
| Nutrition + WSH | 311 | 8.91 | 5.07 | 2.03 | 0.6 | -0.02 (-0.14, 0.11) | 1 | -0.02 (-0.15, 0.11) | 1 | 0.03 (-0.12, 0.17) | 1 | 0 (-0.13, 0.14) | 1 |
| **Ln IL-4 (pg/ml)** |  |  |  |  |  |  |  |  |  |  |  |  |  |
| Control | 285 | 55.54 | 33.48 | 3.87 | 0.54 |  |  |  |  |  |  |  |  |
| Nutrition + WSH | 310 | 65.89 | 53.12 | 3.96 | 0.67 | 0.09 (-0.07, 0.25) | 0.55 | 0.09 (-0.07, 0.25) | 0.58 | 0.12 (-0.04, 0.27) | 0.29 | 0.13 (-0.03, 0.29) | 0.24 |
| **Ln IL-5 (pg/ml)** |  |  |  |  |  |  |  |  |  |  |  |  |  |
| Control | 285 | 1.96 | 1.35 | 0.49 | 0.67 |  |  |  |  |  |  |  |  |
| Nutrition + WSH | 311 | 2.29 | 1.7 | 0.66 | 0.6 | 0.17 (0.05, 0.3) | 0.02 | 0.16 (0.02, 0.29) | 0.05 | 0.21 (0.07, 0.36) | <0.01 | 0.2 (0.08, 0.32) | <0.01 |
| **Ln IL-13 (pg/ml)** |  |  |  |  |  |  |  |  |  |  |  |  |  |
| Control | 285 | 8.33 | 14.23 | 1.6 | 0.96 |  |  |  |  |  |  |  |  |
| Nutrition + WSH | 308 | 9.59 | 11.72 | 1.73 | 1.16 | 0.13 (-0.13, 0.38) | 0.65 | 0.13 (-0.13, 0.39) | 0.67 | 0.14 (-0.13, 0.41) | 0.6 | 0.16 (-0.13, 0.44) | 0.56 |
| **Ln IL-17A (pg/ml)** |  |  |  |  |  |  |  |  |  |  |  |  |  |
| Control | 285 | 5.91 | 4.43 | 1.61 | 0.56 |  |  |  |  |  |  |  |  |
| Nutrition + WSH | 311 | 5.71 | 3.59 | 1.54 | 0.73 | -0.07 (-0.24, 0.09) | 0.74 | -0.09 (-0.25, 0.08) | 0.6 | -0.07 (-0.24, 0.1) | 0.82 | -0.11 (-0.26, 0.03) | 0.26 |
| **Ln IL-21 (pg/ml)** |  |  |  |  |  |  |  |  |  |  |  |  |  |
| Control | 282 | 1.96 | 1.35 | 0.38 | 0.9 |  |  |  |  |  |  |  |  |
| Nutrition + WSH | 305 | 2.12 | 1.64 | 0.46 | 0.86 | 0.09 (-0.12, 0.3) | 0.83 | 0.09 (-0.13, 0.3) | 0.88 | 0.07 (-0.14, 0.28) | 0.99 | 0.08 (-0.14, 0.3) | 0.98 |
| **Ln IL-10 (pg/ml)** |  |  |  |  |  |  |  |  |  |  |  |  |  |
| Control | 284 | 11.07 | 6.11 | 2.23 | 0.69 |  |  |  |  |  |  |  |  |
| Nutrition + WSH | 310 | 12.69 | 9.63 | 2.33 | 0.7 | 0.1 (-0.05, 0.25) | 0.41 | 0.1 (-0.05, 0.26) | 0.39 | 0.12 (-0.03, 0.28) | 0.25 | 0.05 (-0.11, 0.22) | 1 |
| **Ln IL-2 (pg/ml)** |  |  |  |  |  |  |  |  |  |  |  |  |  |
| Control | 283 | 1.34 | 4.48 | -0.4 | 1.18 |  |  |  |  |  |  |  |  |
| Nutrition + WSH | 304 | 1.19 | 0.96 | -0.32 | 1.19 | 0.08 (-0.2, 0.36) | 1 | 0.06 (-0.22, 0.34) | 1 | -0.03 (-0.32, 0.27) | 1 | -0.08 (-0.38, 0.22) | 1 |
| **Ln GM-CSF (pg/ml)** |  |  |  |  |  |  |  |  |  |  |  |  |  |
| Control | 285 | 108.84 | 243.15 | 4.12 | 0.93 |  |  |  |  |  |  |  |  |
| Nutrition + WSH | 311 | 87.37 | 102 | 4.06 | 0.9 | -0.06 (-0.24, 0.11) | 0.96 | -0.01 (-0.23, 0.2) | 1 | 0.02 (-0.18, 0.22) | 1 | 0.02 (-0.16, 0.2) | 1 |
| **Ln AGP (g/L)** |  |  |  |  |  |  |  |  |  |  |  |  |  |
| Control | 253 | 1.09 | 0.5 | -0.02 | 0.46 |  |  |  |  |  |  |  |  |
| Nutrition + WSH | 277 | 1.13 | 0.54 | 0.02 | 0.44 | 0.04 (-0.06, 0.14) | 0.94 | 0.04 (-0.07, 0.14) | 0.98 | 0.02 (-0.07, 0.11) | 1 | 0.02 (-0.08, 0.12) | 1 |
| **Ln IGF-1 (μg/L)** |  |  |  |  |  |  |  |  |  |  |  |  |  |
| Control | 252 | 40.76 | 21.96 | 3.55 | 0.61 |  |  |  |  |  |  |  |  |
| Nutrition + WSH | 277 | 36.42 | 20.18 | 3.44 | 0.6 | -0.11 (-0.22, 0) | 0.12 | -0.1 (-0.21, 0.01) | 0.15 | -0.04 (-0.15, 0.07) | 0.89 | -0.06 (-0.16, 0.04) | 0.47 |

Confidence intervals were adjusted for clustered observations using robust standard errors.

IL = Interleukin; TNF-α = Tumor necrosis factor-α; CRP = C-reactive protein (CRP); IFN-γ = interferon-γ; GM-CSF = Granulocyte-macrophage colony-stimulating factor; AGP = Alpha-1-acid glycoprotein; IGF-1 = Insulin-like growth factor-1

^a^Adjusted for pre-specified covariates: Field staff who collected data, month of measurement, household food insecurity, child age, child sex, mother’s age, mother’s height, mother’s education level, number of children <18 years in the household, number of individuals living in the compound, distance in minutes to the primary water source, household floor materials, household wall materials, household electricity, and household assets (wardrobe, table, chair, clock, khat, chouki, radio, television, refrigerator, bicycle, motorcycle, sewing machine, mobile phone, cattle, goats, and chickens).

^b^Inverse probability of censoring weighting.

^c^P-values shown are Bonferroni corrected to control for familywise error rates

**Supplementary Table 3. Effect of intervention on cytokine ratios at age 14 months**

|  |  |  |  |  |  | Unadjusted difference: Intervention vs. Control | | Age- and sex- adjusted difference: Intervention vs. Control | | Fully adjusted difference: Intervention vs. Control^a^ | | IPCW adjusted difference: Intervention vs. Control^b^ | |
| --- | --- | --- | --- | --- | --- | --- | --- | --- | --- | --- | --- | --- | --- |
| Outcome, Arm | N | Absolute Mean | Absolute SD | Mean | SD | 95% CI | P-value^c^ | 95% CI | P-value^c^ | 95% CI | P-value^c^ | 95% CI | P-value^c^ |
| **Ln IL-1β/IL-10** |  |  |  |  |  |  |  |  |  |  |  |  |  |
| Control | 284 | 0.13 | 0.24 | -2.5 | 0.9 |  |  |  |  |  |  |  |  |
| Nutrition + WSH | 308 | 0.14 | 0.37 | -2.52 | 1.07 | -0.02 (-0.23, 0.19) | 1 | -0.03 (-0.25, 0.18) | 1 | -0.04 (-0.26, 0.18) | 1 | -0.03 (-0.25, 0.19) | 1 |
| **Ln IL-6/IL-10** |  |  |  |  |  |  |  |  |  |  |  |  |  |
| Control | 284 | 0.39 | 1.16 | -1.52 | 0.78 |  |  |  |  |  |  |  |  |
| Nutrition + WSH | 308 | 0.51 | 1.95 | -1.47 | 0.94 | 0.05 (-0.12, 0.22) | 1 | 0.04 (-0.13, 0.21) | 1 | 0.01 (-0.16, 0.18) | 1 | 0.1 (-0.08, 0.28) | 0.54 |
| **Ln TNF-α/IL-10** |  |  |  |  |  |  |  |  |  |  |  |  |  |
| Control | 284 | 1.04 | 3.02 | -0.41 | 0.69 |  |  |  |  |  |  |  |  |
| Nutrition + WSH | 310 | 0.77 | 1 | -0.53 | 0.66 | -0.12 (-0.23, -0.02) | 0.05 | -0.12 (-0.23, -0.01) | 0.06 | -0.11 (-0.22, 0) | 0.09 | -0.1 (-0.21, 0) | 0.12 |
| **Ln IL-12/IL-10** |  |  |  |  |  |  |  |  |  |  |  |  |  |
| Control | 284 | 0.38 | 0.67 | -1.24 | 0.56 |  |  |  |  |  |  |  |  |
| Nutrition + WSH | 309 | 0.3 | 0.35 | -1.44 | 0.7 | -0.19 (-0.31, -0.08) | <0.01 | -0.2 (-0.32, -0.08) | <0.01 | -0.2 (-0.31, -0.08) | <0.01 | -0.18 (-0.3, -0.05) | 0.01 |
| **Ln IFN-γ/IL-10** |  |  |  |  |  |  |  |  |  |  |  |  |  |
| Control | 284 | 1.11 | 2.08 | -0.19 | 0.57 |  |  |  |  |  |  |  |  |
| Nutrition + WSH | 310 | 0.96 | 1.76 | -0.3 | 0.6 | -0.11 (-0.22, 0) | 0.1 | -0.11 (-0.22, 0) | 0.11 | -0.08 (-0.19, 0.03) | 0.26 | -0.06 (-0.17, 0.05) | 0.65 |
| **Ln IL-4/IL-10** |  |  |  |  |  |  |  |  |  |  |  |  |  |
| Control | 284 | 8.02 | 21.37 | 1.64 | 0.69 |  |  |  |  |  |  |  |  |
| Nutrition + WSH | 309 | 7.19 | 12.75 | 1.63 | 0.69 | 0 (-0.14, 0.13) | 1 | -0.01 (-0.15, 0.13) | 1 | -0.01 (-0.14, 0.13) | 1 | 0.07 (-0.06, 0.2) | 0.61 |
| **Ln IL-5/IL-10** |  |  |  |  |  |  |  |  |  |  |  |  |  |
| Control | 284 | 0.28 | 0.72 | -1.74 | 0.75 |  |  |  |  |  |  |  |  |
| Nutrition + WSH | 310 | 0.31 | 0.77 | -1.67 | 0.77 | 0.07 (-0.05, 0.2) | 0.52 | 0.06 (-0.07, 0.19) | 0.69 | 0.06 (-0.06, 0.19) | 0.63 | 0.1 (-0.03, 0.22) | 0.26 |
| **Ln IL-13/IL-10** |  |  |  |  |  |  |  |  |  |  |  |  |  |
| Control | 284 | 1 | 2.36 | -0.63 | 0.94 |  |  |  |  |  |  |  |  |
| Nutrition + WSH | 308 | 0.83 | 0.89 | -0.61 | 1 | 0.02 (-0.17, 0.22) | 1 | 0.02 (-0.18, 0.21) | 1 | 0.04 (-0.16, 0.24) | 1 | 0.05 (-0.15, 0.26) | 1 |
| **Ln IL-17A/IL-10** |  |  |  |  |  |  |  |  |  |  |  |  |  |
| Control | 284 | 0.78 | 1.95 | -0.62 | 0.63 |  |  |  |  |  |  |  |  |
| Nutrition + WSH | 310 | 0.59 | 1.01 | -0.78 | 0.67 | -0.16 (-0.29, -0.03) | 0.04 | -0.18 (-0.31, -0.04) | 0.02 | -0.2 (-0.34, -0.06) | 0.01 | -0.17 (-0.31, -0.03) | 0.04 |
| **Ln IL-21/IL-10** |  |  |  |  |  |  |  |  |  |  |  |  |  |
| Control | 281 | 0.3 | 0.68 | -1.86 | 1.05 |  |  |  |  |  |  |  |  |
| Nutrition + WSH | 304 | 0.27 | 0.61 | -1.87 | 1.01 | -0.01 (-0.21, 0.19) | 1 | -0.02 (-0.22, 0.18) | 1 | -0.08 (-0.28, 0.13) | 0.94 | -0.02 (-0.22, 0.18) | 1 |
| **Ln IL-2/IL-10** |  |  |  |  |  |  |  |  |  |  |  |  |  |
| Control | 282 | 0.18 | 0.67 | -2.65 | 1.19 |  |  |  |  |  |  |  |  |
| Nutrition + WSH | 303 | 0.22 | 1.33 | -2.65 | 1.25 | 0 (-0.26, 0.25) | 1 | 0.01 (-0.24, 0.27) | 1 | -0.12 (-0.39, 0.15) | 0.8 | -0.1 (-0.38, 0.18) | 0.95 |
| **Ln GM-CSF/IL-10** |  |  |  |  |  |  |  |  |  |  |  |  |  |
| Control | 284 | 0.13 | 0.24 | 1.89 | 0.99 |  |  |  |  |  |  |  |  |
| Nutrition + WSH | 310 | 0.14 | 0.37 | 1.73 | 0.96 | -0.16 (-0.32, 0.01) | 0.13 | -0.1 (-0.29, 0.09) | 0.6 | -0.13 (-0.3, 0.05) | 0.3 | -0.14 (-0.34, 0.05) | 0.3 |
| **Ln IL-12/IL-4** |  |  |  |  |  |  |  |  |  |  |  |  |  |
| Control | 285 | 0.06 | 0.02 | -2.89 | 0.5 |  |  |  |  |  |  |  |  |
| Nutrition + WSH | 309 | 0.05 | 0.02 | -3.08 | 0.66 | -0.18 (-0.31, -0.06) | <0.01 | -0.18 (-0.31, -0.06) | <0.01 | -0.18 (-0.29, -0.07) | <0.01 | -0.24 (-0.36, -0.13) | <0.001 |
| **Ln IFN-γ/IL-4** |  |  |  |  |  |  |  |  |  |  |  |  |  |
| Control | 285 | 0.18 | 0.07 | -1.82 | 0.45 |  |  |  |  |  |  |  |  |
| Nutrition + WSH | 310 | 0.17 | 0.09 | -1.93 | 0.55 | -0.11 (-0.2, -0.01) | 0.06 | -0.11 (-0.2, -0.01) | 0.05 | -0.08 (-0.18, 0.02) | 0.27 | -0.13 (-0.25, -0.02) | 0.04 |
| **Ln IL-12/IL-5** |  |  |  |  |  |  |  |  |  |  |  |  |  |
| Control | 285 | 2.15 | 3.54 | 0.49 | 0.65 |  |  |  |  |  |  |  |  |
| Nutrition + WSH | 310 | 1.52 | 0.79 | 0.22 | 0.77 | -0.27 (-0.41, -0.13) | <0.001 | -0.26 (-0.41, -0.11) | <0.001 | -0.23 (-0.38, -0.09) | <0.01 | -0.23 (-0.36, -0.09) | <0.01 |
| **Ln IFN-γ/IL-5** |  |  |  |  |  |  |  |  |  |  |  |  |  |
| Control | 285 | 6.17 | 8.64 | 1.56 | 0.62 |  |  |  |  |  |  |  |  |
| Nutrition + WSH | 311 | 4.92 | 5.61 | 1.37 | 0.68 | -0.19 (-0.32, -0.06) | <0.01 | -0.17 (-0.32, -0.02) | 0.05 | -0.15 (-0.29, 0) | 0.09 | -0.15 (-0.31, 0) | 0.09 |
| **Ln IL-12/IL-13** |  |  |  |  |  |  |  |  |  |  |  |  |  |
| Control | 285 | 0.72 | 0.64 | -0.63 | 0.82 |  |  |  |  |  |  |  |  |
| Nutrition + WSH | 307 | 0.78 | 1.9 | -0.85 | 0.95 | -0.22 (-0.39, -0.05) | 0.02 | -0.22 (-0.39, -0.05) | 0.02 | -0.23 (-0.4, -0.06) | 0.01 | -0.23 (-0.38, -0.08) | <0.01 |
| **Ln IFN-γ/IL-13** |  |  |  |  |  |  |  |  |  |  |  |  |  |
| Control | 285 | 2.12 | 2.53 | 0.44 | 0.77 |  |  |  |  |  |  |  |  |
| Nutrition + WSH | 308 | 2.42 | 5.22 | 0.3 | 0.92 | -0.14 (-0.3, 0.02) | 0.15 | -0.14 (-0.3, 0.01) | 0.15 | -0.14 (-0.3, 0.02) | 0.19 | -0.16 (-0.33, 0.01) | 0.14 |
| **Ln IL-12/IL-17A** |  |  |  |  |  |  |  |  |  |  |  |  |  |
| Control | 285 | 0.57 | 0.2 | -0.64 | 0.45 |  |  |  |  |  |  |  |  |
| Nutrition + WSH | 310 | 0.61 | 0.51 | -0.66 | 0.6 | -0.02 (-0.12, 0.08) | 1 | -0.01 (-0.12, 0.09) | 1 | 0.02 (-0.09, 0.12) | 1 | 0 (-0.11, 0.11) | 1 |
| **Ln IFN-γ/IL-17A** |  |  |  |  |  |  |  |  |  |  |  |  |  |
| Control | 285 | 1.71 | 0.86 | 0.43 | 0.48 |  |  |  |  |  |  |  |  |
| Nutrition + WSH | 311 | 2.09 | 4.94 | 0.49 | 0.52 | 0.06 (-0.07, 0.18) | 0.73 | 0.06 (-0.06, 0.19) | 0.62 | 0.08 (-0.04, 0.19) | 0.39 | 0.07 (-0.09, 0.23) | 0.81 |
| **Ln IL-12/IL-21** |  |  |  |  |  |  |  |  |  |  |  |  |  |
| Control | 282 | 3.16 | 5.65 | 0.6 | 0.96 |  |  |  |  |  |  |  |  |
| Nutrition + WSH | 304 | 2.72 | 5.5 | 0.42 | 1.08 | -0.19 (-0.4, 0.03) | 0.18 | -0.19 (-0.41, 0.03) | 0.19 | -0.11 (-0.36, 0.13) | 0.74 | -0.15 (-0.36, 0.06) | 0.31 |
| **Ln IFN-γ/IL-21** |  |  |  |  |  |  |  |  |  |  |  |  |  |
| Control | 282 | 9.13 | 16.22 | 1.67 | 0.91 |  |  |  |  |  |  |  |  |
| Nutrition + WSH | 305 | 8.37 | 19.41 | 1.57 | 0.92 | -0.1 (-0.29, 0.1) | 0.66 | -0.1 (-0.3, 0.1) | 0.63 | -0.03 (-0.24, 0.18) | 1 | -0.07 (-0.28, 0.14) | 1 |
| **Ln Pro-inflammatory cytokines^d^/IL-10** |  |  |  |  |  |  |  |  |  |  |  |  |  |
| Control | 284 |  |  | 1.29 | 0.64 |  |  |  |  |  |  |  |  |
| Nutrition + WSH | 306 |  |  | 1.3 | 0.66 | 0.01 (-0.11, 0.13) | 1 | 0 (-0.12, 0.12) | 1 | -0.02 (-0.14, 0.1) | 1 | 0.03 (-0.09, 0.15) | 1 |
| **Ln Th1^e^/IL-10** |  |  |  |  |  |  |  |  |  |  |  |  |  |
| Control | 284 |  |  | 1.18 | 0.54 |  |  |  |  |  |  |  |  |
| Nutrition + WSH | 309 |  |  | 1.05 | 0.55 | -0.13 (-0.23, -0.03) | 0.02 | -0.14 (-0.24, -0.04) | 0.01 | -0.13 (-0.24, -0.02) | 0.03 | -0.11 (-0.22, -0.01) | 0.06 |
| **Ln Th2^f^/IL-10** |  |  |  |  |  |  |  |  |  |  |  |  |  |
| Control | 284 |  |  | 0.85 | 0.67 |  |  |  |  |  |  |  |  |
| Nutrition + WSH | 307 |  |  | 0.9 | 0.64 | 0.05 (-0.06, 0.15) | 0.76 | 0.04 (-0.07, 0.14) | 1 | 0.06 (-0.04, 0.16) | 0.46 | 0.1 (0.01, 0.2) | 0.06 |
| **Ln Th17^g^/IL-10** |  |  |  |  |  |  |  |  |  |  |  |  |  |
| Control | 281 |  |  | 0.76 | 0.69 |  |  |  |  |  |  |  |  |
| Nutrition + WSH | 304 |  |  | 0.67 | 0.68 | -0.09 (-0.21, 0.03) | 0.28 | -0.1 (-0.22, 0.02) | 0.18 | -0.11 (-0.22, 0) | 0.11 | -0.1 (-0.22, 0.02) | 0.21 |
| **Ln Th1^e^/Th2^f^** |  |  |  |  |  |  |  |  |  |  |  |  |  |
| Control | 285 |  |  | 0.32 | 0.4 |  |  |  |  |  |  |  |  |
| Nutrition + WSH | 306 |  |  | 0.14 | 0.47 | -0.18 (-0.25, -0.11) | <0.001 | -0.18 (-0.26, -0.1) | <0.001 | -0.17 (-0.25, -0.09) | <0.001 | -0.22 (-0.29, -0.15) | <0.001 |
| **Ln Th1^e^/Th17^g^** |  |  |  |  |  |  |  |  |  |  |  |  |  |
| Control | 282 |  |  | 0.41 | 0.47 |  |  |  |  |  |  |  |  |
| Nutrition + WSH | 304 |  |  | 0.37 | 0.51 | -0.04 (-0.15, 0.06) | 0.82 | -0.04 (-0.15, 0.07) | 0.9 | 0 (-0.13, 0.12) | 1 | 0.02 (-0.09, 0.13) | 1 |

Confidence intervals were adjusted for clustered observations using robust standard errors.

IL = Interleukin; TNF-α = Tumor necrosis factor-α; CRP = C-reactive protein (CRP); IFN-γ = interferon-γ; GM-CSF = Granulocyte-macrophage colony-stimulating factor; AGP = Alpha-1-acid glycoprotein; IGF-1 = Insulin-like growth factor-1

^a^Adjusted for pre-specified covariates: Field staff who collected data, month of measurement, household food insecurity, child age, child sex, mother’s age, mother’s height, mother’s education level, number of children <18 years in the household, number of individuals living in the compound, distance in minutes to the primary water source, household floor materials, household wall materials, household electricity, and household assets (wardrobe, table, chair, clock, khat, chouki, radio, television, refrigerator, bicycle, motorcycle, sewing machine, mobile phone, cattle, goats, and chickens).

^b^Inverse probability of censoring weighting.

^c^P-values shown are Bonferroni corrected to control for familywise error rates

^d^Pro-inflammatory cytokines: IL-1β, IL-6, TNF-α

^e^Th1 cytokines: IL-12, IFN-γ

^f^Th2 cytokines: IL-4, IL-5, IL-13

^g^Th17 cytokines: IL-17A, IL-21

**Supplementary Table 4. Effect of intervention on individual immune status and growth factor measurements at age 28 months**

|  |  |  |  |  |  | Unadjusted difference: Intervention vs. Control | | Age- and sex- adjusted difference: Intervention vs. Control | | Fully adjusted difference: Intervention vs. Control^a^ | | IPCW adjusted difference: Intervention vs. Control^b^ | |
| --- | --- | --- | --- | --- | --- | --- | --- | --- | --- | --- | --- | --- | --- |
| Outcome, Arm | N | Absolute Mean | Absolute SD | Mean | SD | 95% CI | P-value^c^ | 95% CI | P-value^c^ | 95% CI | P-value^c^ | 95% CI | P-value^c^ |
| **Ln IL-1β (pg/ml)** |  |  |  |  |  |  |  |  |  |  |  |  |  |
| Control | 313 | 1.21 | 0.74 | -0.12 | 0.98 |  |  |  |  |  |  |  |  |
| Nutrition + WSH | 365 | 1.19 | 0.81 | -0.24 | 1.14 | -0.12 (-0.39, 0.15) | 0.79 | -0.12 (-0.39, 0.15) | 0.78 | -0.14 (-0.4, 0.12) | 0.6 | -0.04 (-0.35, 0.28) | 1 |
| **Ln IL-6 (pg/ml)** |  |  |  |  |  |  |  |  |  |  |  |  |  |
| Control | 313 | 2.78 | 2.4 | 0.76 | 0.76 |  |  |  |  |  |  |  |  |
| Nutrition + WSH | 366 | 2.45 | 1.93 | 0.54 | 0.96 | -0.21 (-0.44, 0.02) | 0.13 | -0.21 (-0.44, 0.02) | 0.13 | -0.22 (-0.45, 0.01) | 0.13 | -0.19 (-0.41, 0.03) | 0.19 |
| **Ln TNF-α (pg/ml)** |  |  |  |  |  |  |  |  |  |  |  |  |  |
| Control | 313 | 5.03 | 2.55 | 1.48 | 0.57 |  |  |  |  |  |  |  |  |
| Nutrition + WSH | 367 | 5.3 | 2.59 | 1.54 | 0.53 | 0.06 (-0.08, 0.2) | 0.78 | 0.05 (-0.09, 0.19) | 0.95 | 0.04 (-0.09, 0.17) | 1 | 0.06 (-0.08, 0.21) | 0.77 |
| **Ln CRP (mg/L)** |  |  |  |  |  |  |  |  |  |  |  |  |  |
| Control | 310 | 2.04 | 3.93 | -0.16 | 1.25 |  |  |  |  |  |  |  |  |
| Nutrition + WSH | 372 | 3.39 | 10.37 | -0.11 | 1.41 | 0.05 (-0.15, 0.26) | 1 | 0.05 (-0.16, 0.25) | 1 | 0.02 (-0.17, 0.22) | 1 | -0.11 (-0.37, 0.14) | 0.77 |
| **Ln IL-12 (pg/ml)** |  |  |  |  |  |  |  |  |  |  |  |  |  |
| Control | 313 | 2.82 | 1.45 | 0.89 | 0.64 |  |  |  |  |  |  |  |  |
| Nutrition + WSH | 367 | 2.7 | 1.4 | 0.82 | 0.7 | -0.06 (-0.23, 0.11) | 0.92 | -0.07 (-0.23, 0.1) | 0.89 | -0.08 (-0.22, 0.07) | 0.57 | 0 (-0.18, 0.19) | 1 |
| **Ln IFN-γ (pg/ml)** |  |  |  |  |  |  |  |  |  |  |  |  |  |
| Control | 313 | 8.95 | 7.24 | 2.01 | 0.64 |  |  |  |  |  |  |  |  |
| Nutrition + WSH | 367 | 7.87 | 4.4 | 1.9 | 0.61 | -0.1 (-0.26, 0.06) | 0.43 | -0.1 (-0.26, 0.06) | 0.41 | -0.09 (-0.23, 0.05) | 0.41 | 0 (-0.21, 0.21) | 1 |
| **Ln IL-4 (pg/ml)** |  |  |  |  |  |  |  |  |  |  |  |  |  |
| Control | 313 | 57.6 | 35.87 | 3.89 | 0.58 |  |  |  |  |  |  |  |  |
| Nutrition + WSH | 367 | 52.25 | 43.12 | 3.73 | 0.67 | -0.16 (-0.32, 0) | 0.1 | -0.15 (-0.31, 0.01) | 0.14 | -0.15 (-0.31, 0.01) | 0.15 | -0.18 (-0.36, 0.01) | 0.13 |
| **Ln IL-5 (pg/ml)** |  |  |  |  |  |  |  |  |  |  |  |  |  |
| Control | 313 | 1.86 | 1.35 | 0.39 | 0.79 |  |  |  |  |  |  |  |  |
| Nutrition + WSH | 367 | 1.79 | 1.13 | 0.39 | 0.7 | 0.01 (-0.15, 0.16) | 1 | 0.01 (-0.15, 0.16) | 1 | -0.02 (-0.18, 0.14) | 1 | 0.04 (-0.15, 0.24) | 1 |
| **Ln IL-13 (pg/ml)** |  |  |  |  |  |  |  |  |  |  |  |  |  |
| Control | 311 | 9.41 | 18.67 | 1.58 | 1.16 |  |  |  |  |  |  |  |  |
| Nutrition + WSH | 366 | 6.92 | 8.3 | 1.43 | 1.12 | -0.15 (-0.47, 0.17) | 0.73 | -0.15 (-0.48, 0.17) | 0.71 | -0.13 (-0.4, 0.14) | 0.67 | -0.09 (-0.44, 0.26) | 1 |
| **Ln IL-17A (pg/ml)** |  |  |  |  |  |  |  |  |  |  |  |  |  |
| Control | 313 | 5.09 | 3.52 | 1.45 | 0.63 |  |  |  |  |  |  |  |  |
| Nutrition + WSH | 367 | 5.01 | 2.93 | 1.43 | 0.65 | -0.02 (-0.19, 0.16) | 1 | -0.02 (-0.19, 0.16) | 1 | -0.02 (-0.17, 0.13) | 1 | -0.05 (-0.27, 0.17) | 1 |
| **Ln IL-21 (pg/ml)** |  |  |  |  |  |  |  |  |  |  |  |  |  |
| Control | 308 | 1.9 | 1.3 | 0.37 | 0.86 |  |  |  |  |  |  |  |  |
| Nutrition + WSH | 362 | 2.07 | 5.7 | 0.24 | 0.97 | -0.13 (-0.31, 0.06) | 0.37 | -0.13 (-0.31, 0.06) | 0.35 | -0.1 (-0.28, 0.09) | 0.59 | -0.11 (-0.29, 0.06) | 0.41 |
| **Ln IL-10 (pg/ml)** |  |  |  |  |  |  |  |  |  |  |  |  |  |
| Control | 312 | 9.69 | 14.78 | 2.02 | 0.67 |  |  |  |  |  |  |  |  |
| Nutrition + WSH | 363 | 10.32 | 8.01 | 2.09 | 0.78 | 0.07 (-0.08, 0.22) | 0.78 | 0.06 (-0.09, 0.21) | 0.82 | 0.06 (-0.08, 0.2) | 0.76 | 0.11 (-0.05, 0.26) | 0.34 |
| **Ln IL-2 (pg/ml)** |  |  |  |  |  |  |  |  |  |  |  |  |  |
| Control | 310 | 1.18 | 0.85 | -0.23 | 1.08 |  |  |  |  |  |  |  |  |
| Nutrition + WSH | 361 | 1.57 | 6.67 | -0.35 | 1.23 | -0.12 (-0.37, 0.12) | 0.64 | -0.12 (-0.37, 0.12) | 0.64 | -0.11 (-0.35, 0.14) | 0.79 | -0.1 (-0.34, 0.15) | 0.86 |
| **Ln GM-CSF (pg/ml)** |  |  |  |  |  |  |  |  |  |  |  |  |  |
| Control | 313 | 150.92 | 188.02 | 4.54 | 1.01 |  |  |  |  |  |  |  |  |
| Nutrition + WSH | 367 | 176.35 | 255.09 | 4.63 | 1.07 | 0.08 (-0.14, 0.31) | 0.94 | 0.08 (-0.15, 0.31) | 0.96 | 0.06 (-0.13, 0.25) | 1 | 0.18 (-0.03, 0.39) | 0.2 |
| **Ln AGP (g/L)** |  |  |  |  |  |  |  |  |  |  |  |  |  |
| Control | 108 | 0.9 | 0.57 | -0.23 | 0.48 |  |  |  |  |  |  |  |  |
| Nutrition + WSH | 243 | 0.98 | 0.68 | -0.18 | 0.52 | 0.06 (-0.04, 0.15) | 0.46 | 0.07 (-0.03, 0.16) | 0.38 | 0.08 (-0.02, 0.17) | 0.21 | 0.2 (0.11, 0.29) | <0.001 |
| **Ln IGF-1 (μg/L)** |  |  |  |  |  |  |  |  |  |  |  |  |  |
| Control | 325 | 52.87 | 25.1 | 3.86 | 0.48 |  |  |  |  |  |  |  |  |
| Nutrition + WSH | 379 | 50.45 | 25.02 | 3.8 | 0.5 | -0.06 (-0.13, 0.02) | 0.28 | -0.06 (-0.14, 0.02) | 0.23 | -0.01 (-0.08, 0.07) | 1 | 0.01 (-0.06, 0.08) | 1 |

Confidence intervals were adjusted for clustered observations using robust standard errors.

IL = Interleukin; TNF-α = Tumor necrosis factor-α; CRP = C-reactive protein (CRP); IFN-γ = interferon-γ; GM-CSF = Granulocyte-macrophage colony-stimulating factor; AGP = Alpha-1-acid glycoprotein; IGF-1 = Insulin-like growth factor-1

^a^Adjusted for pre-specified covariates: Field staff who collected data, month of measurement, household food insecurity, child age, child sex, mother’s age, mother’s height, mother’s education level, number of children <18 years in the household, number of individuals living in the compound, distance in minutes to the primary water source, household floor materials, household wall materials, household electricity, and household assets (wardrobe, table, chair, clock, khat, chouki, radio, television, refrigerator, bicycle, motorcycle, sewing machine, mobile phone, cattle, goats, and chickens).

^b^Inverse probability of censoring weighting.

^c^P-values shown are Bonferroni corrected to control for familywise error rates

**Supplementary Table 5. Effect of intervention on cytokine ratios at age 28 months**

|  |  |  |  |  |  | Unadjusted difference: Intervention vs. Control | | Age- and sex- adjusted difference: Intervention vs. Control | | Fully adjusted difference: Intervention vs. Control^a^ | | IPCW adjusted difference: Intervention vs. Control^b^ | |
| --- | --- | --- | --- | --- | --- | --- | --- | --- | --- | --- | --- | --- | --- |
| Outcome, Arm | N | Absolute Mean | Absolute SD | Mean | SD | 95% CI | P-value^c^ | 95% CI | P-value^c^ | 95% CI | P-value^c^ | 95% CI | P-value^c^ |
| **Ln IL-1β/IL-10** |  |  |  |  |  |  |  |  |  |  |  |  |  |
| Control | 312 | 0.15 | 0.11 | -2.14 | 0.85 |  |  |  |  |  |  |  |  |
| Nutrition + WSH | 361 | 0.17 | 0.36 | -2.34 | 1.06 | -0.2 (-0.42, 0.02) | 0.14 | -0.17 (-0.39, 0.05) | 0.25 | -0.19 (-0.41, 0.03) | 0.19 | -0.18 (-0.43, 0.07) | 0.32 |
| **Ln IL-6/IL-10** |  |  |  |  |  |  |  |  |  |  |  |  |  |
| Control | 312 | 0.45 | 0.86 | -1.26 | 0.82 |  |  |  |  |  |  |  |  |
| Nutrition + WSH | 362 | 0.48 | 1.52 | -1.53 | 1.03 | -0.27 (-0.48, -0.07) | 0.02 | -0.27 (-0.48, -0.06) | 0.02 | -0.26 (-0.47, -0.05) | 0.03 | -0.14 (-0.41, 0.13) | 0.6 |
| **Ln TNF-α/IL-10** |  |  |  |  |  |  |  |  |  |  |  |  |  |
| Control | 312 | 0.75 | 0.97 | -0.54 | 0.65 |  |  |  |  |  |  |  |  |
| Nutrition + WSH | 363 | 0.95 | 2.36 | -0.54 | 0.77 | 0 (-0.11, 0.1) | 1 | 0 (-0.11, 0.11) | 1 | 0 (-0.12, 0.12) | 1 | 0 (-0.16, 0.16) | 1 |
| **Ln IL-12/IL-10** |  |  |  |  |  |  |  |  |  |  |  |  |  |
| Control | 312 | 0.38 | 0.28 | -1.13 | 0.58 |  |  |  |  |  |  |  |  |
| Nutrition + WSH | 363 | 0.37 | 0.46 | -1.25 | 0.66 | -0.12 (-0.22, -0.02) | 0.04 | -0.11 (-0.21, 0) | 0.11 | -0.09 (-0.2, 0.01) | 0.18 | -0.07 (-0.19, 0.05) | 0.5 |
| **Ln IFN-γ/IL-10** |  |  |  |  |  |  |  |  |  |  |  |  |  |
| Control | 312 | 1.15 | 0.8 | -0.01 | 0.57 |  |  |  |  |  |  |  |  |
| Nutrition + WSH | 363 | 1.08 | 1.6 | -0.18 | 0.62 | -0.16 (-0.27, -0.06) | <0.01 | -0.15 (-0.25, -0.04) | 0.02 | -0.13 (-0.23, -0.02) | 0.04 | -0.14 (-0.26, -0.02) | 0.04 |
| **Ln IL-4/IL-10** |  |  |  |  |  |  |  |  |  |  |  |  |  |
| Control | 312 | 8.06 | 6.71 | 1.88 | 0.64 |  |  |  |  |  |  |  |  |
| Nutrition + WSH | 363 | 7.48 | 13.48 | 1.65 | 0.71 | -0.23 (-0.35, -0.1) | <0.001 | -0.2 (-0.33, -0.07) | <0.01 | -0.19 (-0.32, -0.06) | 0.01 | -0.25 (-0.4, -0.1) | <0.01 |
| **Ln IL-5/IL-10** |  |  |  |  |  |  |  |  |  |  |  |  |  |
| Control | 312 | 0.26 | 0.26 | -1.63 | 0.79 |  |  |  |  |  |  |  |  |
| Nutrition + WSH | 363 | 0.33 | 0.99 | -1.68 | 0.84 | -0.05 (-0.19, 0.09) | 0.91 | -0.05 (-0.19, 0.09) | 0.94 | -0.09 (-0.23, 0.05) | 0.42 | -0.06 (-0.23, 0.1) | 0.89 |
| **Ln IL-13/IL-10** |  |  |  |  |  |  |  |  |  |  |  |  |  |
| Control | 311 | 1.13 | 1.89 | -0.44 | 1.04 |  |  |  |  |  |  |  |  |
| Nutrition + WSH | 362 | 0.87 | 1.9 | -0.65 | 0.98 | -0.21 (-0.44, 0.01) | 0.13 | -0.21 (-0.44, 0.01) | 0.13 | -0.19 (-0.4, 0.01) | 0.13 | -0.2 (-0.45, 0.05) | 0.24 |
| **Ln IL-17A/IL-10** |  |  |  |  |  |  |  |  |  |  |  |  |  |
| Control | 312 | 0.68 | 0.59 | -0.57 | 0.61 |  |  |  |  |  |  |  |  |
| Nutrition + WSH | 363 | 0.71 | 1.04 | -0.65 | 0.69 | -0.08 (-0.18, 0.03) | 0.32 | -0.05 (-0.17, 0.06) | 0.69 | -0.04 (-0.15, 0.07) | 0.93 | -0.07 (-0.18, 0.04) | 0.39 |
| **Ln IL-21/IL-10** |  |  |  |  |  |  |  |  |  |  |  |  |  |
| Control | 307 | 0.34 | 0.62 | -1.65 | 1.06 |  |  |  |  |  |  |  |  |
| Nutrition + WSH | 358 | 0.43 | 1.56 | -1.83 | 1.21 | -0.19 (-0.4, 0.03) | 0.19 | -0.19 (-0.4, 0.03) | 0.19 | -0.16 (-0.37, 0.05) | 0.27 | -0.19 (-0.49, 0.11) | 0.43 |
| **Ln IL-2/IL-10** |  |  |  |  |  |  |  |  |  |  |  |  |  |
| Control | 309 | 0.2 | 0.54 | -2.25 | 1.09 |  |  |  |  |  |  |  |  |
| Nutrition + WSH | 357 | 0.36 | 2.69 | -2.44 | 1.35 | -0.19 (-0.43, 0.06) | 0.29 | -0.16 (-0.4, 0.09) | 0.42 | -0.15 (-0.4, 0.09) | 0.45 | -0.06 (-0.36, 0.24) | 1 |
| **Ln GM-CSF/IL-10** |  |  |  |  |  |  |  |  |  |  |  |  |  |
| Control | 312 | 18.88 | 22.62 | 2.53 | 0.92 |  |  |  |  |  |  |  |  |
| Nutrition + WSH | 363 | 22.27 | 35.78 | 2.55 | 1.02 | 0.02 (-0.16, 0.2) | 1 | 0.05 (-0.14, 0.24) | 1 | 0.03 (-0.14, 0.2) | 1 | 0.09 (-0.07, 0.25) | 0.54 |
| **Ln IL-12/IL-4** |  |  |  |  |  |  |  |  |  |  |  |  |  |
| Control | 313 | 0.06 | 0.02 | -3.01 | 0.52 |  |  |  |  |  |  |  |  |
| Nutrition + WSH | 367 | 0.06 | 0.03 | -2.91 | 0.57 | 0.1 (-0.02, 0.21) | 0.18 | 0.1 (-0.02, 0.21) | 0.18 | 0.1 (-0.01, 0.21) | 0.14 | 0.2 (0.07, 0.34) | <0.01 |
| **Ln IFN-γ/IL-4** |  |  |  |  |  |  |  |  |  |  |  |  |  |
| Control | 313 | 0.18 | 0.12 | -1.89 | 0.58 |  |  |  |  |  |  |  |  |
| Nutrition + WSH | 367 | 0.18 | 0.09 | -1.83 | 0.53 | 0.06 (-0.05, 0.16) | 0.54 | 0.06 (-0.05, 0.16) | 0.54 | 0.07 (-0.04, 0.17) | 0.45 | 0.12 (-0.03, 0.27) | 0.23 |
| **Ln IL-12/IL-5** |  |  |  |  |  |  |  |  |  |  |  |  |  |
| Control | 313 | 2.53 | 4.89 | 0.5 | 0.83 |  |  |  |  |  |  |  |  |
| Nutrition + WSH | 367 | 2.14 | 3.36 | 0.43 | 0.81 | -0.07 (-0.23, 0.09) | 0.77 | -0.07 (-0.23, 0.09) | 0.77 | -0.03 (-0.18, 0.13) | 1 | -0.05 (-0.28, 0.17) | 1 |
| **Ln IFN-γ/IL-5** |  |  |  |  |  |  |  |  |  |  |  |  |  |
| Control | 313 | 8.44 | 19.1 | 1.62 | 0.86 |  |  |  |  |  |  |  |  |
| Nutrition + WSH | 367 | 6.4 | 9.42 | 1.51 | 0.78 | -0.11 (-0.26, 0.05) | 0.34 | -0.11 (-0.26, 0.05) | 0.34 | -0.06 (-0.21, 0.1) | 0.93 | -0.03 (-0.22, 0.16) | 1 |
| **Ln IL-12/IL-13** |  |  |  |  |  |  |  |  |  |  |  |  |  |
| Control | 311 | 0.83 | 1.56 | -0.69 | 0.93 |  |  |  |  |  |  |  |  |
| Nutrition + WSH | 366 | 0.93 | 1.96 | -0.61 | 0.92 | 0.08 (-0.11, 0.28) | 0.81 | 0.08 (-0.11, 0.28) | 0.8 | 0.07 (-0.1, 0.25) | 0.86 | 0.04 (-0.21, 0.28) | 1 |
| **Ln IFN-γ/IL-13** |  |  |  |  |  |  |  |  |  |  |  |  |  |
| Control | 311 | 2.76 | 7.25 | 0.43 | 0.92 |  |  |  |  |  |  |  |  |
| Nutrition + WSH | 366 | 2.77 | 5.96 | 0.47 | 0.89 | 0.04 (-0.16, 0.25) | 1 | 0.05 (-0.16, 0.25) | 1 | 0.04 (-0.14, 0.22) | 1 | -0.02 (-0.22, 0.19) | 1 |
| **Ln IL-12/IL-17A** |  |  |  |  |  |  |  |  |  |  |  |  |  |
| Control | 313 | 0.69 | 0.78 | -0.56 | 0.6 |  |  |  |  |  |  |  |  |
| Nutrition + WSH | 367 | 0.61 | 0.42 | -0.61 | 0.5 | -0.05 (-0.15, 0.06) | 0.73 | -0.05 (-0.15, 0.06) | 0.73 | -0.06 (-0.15, 0.04) | 0.49 | -0.01 (-0.13, 0.12) | 1 |
| **Ln IFN-γ/IL-17A** |  |  |  |  |  |  |  |  |  |  |  |  |  |
| Control | 313 | 2.16 | 3.34 | 0.56 | 0.58 |  |  |  |  |  |  |  |  |
| Nutrition + WSH | 367 | 1.9 | 2.1 | 0.47 | 0.54 | -0.09 (-0.19, 0.02) | 0.22 | -0.09 (-0.19, 0.02) | 0.22 | -0.07 (-0.18, 0.03) | 0.35 | -0.03 (-0.16, 0.09) | 1 |
| **Ln IL-12/IL-21** |  |  |  |  |  |  |  |  |  |  |  |  |  |
| Control | 308 | 3.06 | 6.18 | 0.52 | 1 |  |  |  |  |  |  |  |  |
| Nutrition + WSH | 362 | 3.37 | 6.05 | 0.57 | 1.14 | 0.05 (-0.18, 0.28) | 1 | 0.05 (-0.18, 0.28) | 1 | 0.03 (-0.18, 0.24) | 1 | 0.11 (-0.2, 0.43) | 0.97 |
| **Ln IFN-γ/IL-21** |  |  |  |  |  |  |  |  |  |  |  |  |  |
| Control | 308 | 10.26 | 23.31 | 1.64 | 1.05 |  |  |  |  |  |  |  |  |
| Nutrition + WSH | 362 | 9.92 | 18.41 | 1.66 | 1.1 | 0.02 (-0.22, 0.25) | 1 | 0.02 (-0.22, 0.25) | 1 | 0.02 (-0.21, 0.26) | 1 | 0.01 (-0.3, 0.32) | 1 |
| **Ln Pro-inflammatory cytokines^d^/IL-10** |  |  |  |  |  |  |  |  |  |  |  |  |  |
| Control | 312 |  |  | 1.89 | 0.57 |  |  |  |  |  |  |  |  |
| Nutrition + WSH | 360 |  |  | 1.79 | 0.7 | -0.09 (-0.21, 0.03) | 0.26 | -0.09 (-0.21, 0.03) | 0.27 | -0.09 (-0.21, 0.03) | 0.31 | -0.05 (-0.23, 0.13) | 1 |
| **Ln Th1^e^/IL-10** |  |  |  |  |  |  |  |  |  |  |  |  |  |
| Control | 312 |  |  | 1.55 | 0.49 |  |  |  |  |  |  |  |  |
| Nutrition + WSH | 363 |  |  | 1.41 | 0.6 | -0.14 (-0.23, -0.06) | <0.01 | -0.13 (-0.22, -0.04) | <0.01 | -0.12 (-0.21, -0.02) | 0.03 | -0.11 (-0.21, -0.02) | 0.05 |
| **Ln Th2^f^/IL-10** |  |  |  |  |  |  |  |  |  |  |  |  |  |
| Control | 311 |  |  | 1.55 | 0.61 |  |  |  |  |  |  |  |  |
| Nutrition + WSH | 362 |  |  | 1.4 | 0.71 | -0.15 (-0.26, -0.04) | 0.02 | -0.14 (-0.25, -0.03) | 0.03 | -0.15 (-0.25, -0.04) | 0.02 | -0.17 (-0.3, -0.04) | 0.02 |
| **Ln Th17^g^/IL-10** |  |  |  |  |  |  |  |  |  |  |  |  |  |
| Control | 307 |  |  | 1.02 | 0.58 |  |  |  |  |  |  |  |  |
| Nutrition + WSH | 358 |  |  | 0.95 | 0.7 | -0.08 (-0.18, 0.02) | 0.27 | -0.05 (-0.16, 0.05) | 0.62 | -0.04 (-0.14, 0.06) | 0.93 | -0.03 (-0.13, 0.07) | 1 |
| **Ln Th1^e^/Th2^f^** |  |  |  |  |  |  |  |  |  |  |  |  |  |
| Control | 311 |  |  | 0 | 0.51 |  |  |  |  |  |  |  |  |
| Nutrition + WSH | 366 |  |  | 0.01 | 0.5 | 0 (-0.09, 0.1) | 1 | 0 (-0.09, 0.1) | 1 | 0.01 (-0.09, 0.1) | 1 | 0.02 (-0.13, 0.16) | 1 |
| **Ln Th1^e^/Th17^g^** |  |  |  |  |  |  |  |  |  |  |  |  |  |
| Control | 308 |  |  | 0.53 | 0.45 |  |  |  |  |  |  |  |  |
| Nutrition + WSH | 362 |  |  | 0.46 | 0.49 | -0.07 (-0.16, 0.03) | 0.31 | -0.07 (-0.16, 0.03) | 0.31 | -0.04 (-0.13, 0.05) | 0.88 | -0.02 (-0.12, 0.07) | 1 |

Confidence intervals were adjusted for clustered observations using robust standard errors.

IL = Interleukin; TNF-α = Tumor necrosis factor-α; CRP = C-reactive protein (CRP); IFN-γ = interferon-γ; GM-CSF = Granulocyte-macrophage colony-stimulating factor; AGP = Alpha-1-acid glycoprotein; IGF-1 = Insulin-like growth factor-1

^a^Adjusted for pre-specified covariates: Field staff who collected data, month of measurement, household food insecurity, child age, child sex, mother’s age, mother’s height, mother’s education level, number of children <18 years in the household, number of individuals living in the compound, distance in minutes to the primary water source, household floor materials, household wall materials, household electricity, and household assets (wardrobe, table, chair, clock, khat, chouki, radio, television, refrigerator, bicycle, motorcycle, sewing machine, mobile phone, cattle, goats, and chickens).

^b^Inverse probability of censoring weighting.

^c^P-values shown are Bonferroni corrected to control for familywise error rates

^d^Pro-inflammatory cytokines: IL-1β, IL-6, TNF-α

^e^Th1 cytokines: IL-12, IFN-γ

^f^Th2 cytokines: IL-4, IL-5, IL-13

^g^Th17 cytokines: IL-17A, IL-21

**Supplementary Table 6. Effect of intervention on change in cytokine ratios between ages 14 and 28 months**

|  |  |  |  |  |  | Unadjusted difference: Intervention vs. Control | | Age- and sex- adjusted difference: Intervention vs. Control | | Fully adjusted difference: Intervention vs. Control^a^ | | IPCW adjusted difference: Intervention vs. Control^b^ | |
| --- | --- | --- | --- | --- | --- | --- | --- | --- | --- | --- | --- | --- | --- |
| Outcome, Arm | N | Absolute Mean | Absolute SD | Mean | SD | 95% CI | P-value^c^ | 95% CI | P-value^c^ | 95% CI | P-value^c^ | 95% CI | P-value^c^ |
| **Ln ΔIL-1β/IL-10** |  |  |  |  |  |  |  |  |  |  |  |  |  |
| Control | 216 | 0.02 | 0.26 | 0.35 | 1.08 |  |  |  |  |  |  |  |  |
| Nutrition + WSH | 261 | 0.05 | 0.4 | 0.21 | 1.37 | -0.14 (-0.39, 0.11) | 0.52 | -0.13 (-0.38, 0.12) | 0.65 | -0.15 (-0.41, 0.12) | 0.55 | -0.14 (-0.4, 0.12) | 0.6 |
| **Ln ΔIL-6/IL-10** |  |  |  |  |  |  |  |  |  |  |  |  |  |
| Control | 216 | 0.12 | 1.15 | 0.24 | 1.04 |  |  |  |  |  |  |  |  |
| Nutrition + WSH | 264 | 0.13 | 1.99 | -0.02 | 1.32 | -0.27 (-0.51, -0.03) | 0.05 | -0.24 (-0.48, -0.01) | 0.09 | -0.2 (-0.48, 0.07) | 0.3 | -0.2 (-0.45, 0.06) | 0.27 |
| **Ln ΔTNF-α/IL-10** |  |  |  |  |  |  |  |  |  |  |  |  |  |
| Control | 216 | -0.37 | 3.57 | -0.14 | 0.86 |  |  |  |  |  |  |  |  |
| Nutrition + WSH | 265 | 0.3 | 2.76 | 0.03 | 0.93 | 0.17 (-0.02, 0.36) | 0.14 | 0.17 (-0.02, 0.36) | 0.16 | 0.1 (-0.08, 0.28) | 0.57 | 0.17 (-0.06, 0.39) | 0.3 |
| **Ln ΔIL-12/IL-10** |  |  |  |  |  |  |  |  |  |  |  |  |  |
| Control | 216 | 0 | 0.78 | 0.12 | 0.84 |  |  |  |  |  |  |  |  |
| Nutrition + WSH | 264 | 0.1 | 0.48 | 0.22 | 0.88 | 0.11 (-0.09, 0.31) | 0.59 | 0.12 (-0.08, 0.33) | 0.47 | 0.12 (-0.07, 0.31) | 0.45 | 0.11 (-0.08, 0.3) | 0.49 |
| **Ln ΔIFN-γ/IL-10** |  |  |  |  |  |  |  |  |  |  |  |  |  |
| Control | 216 | 0.02 | 2.41 | 0.16 | 0.85 |  |  |  |  |  |  |  |  |
| Nutrition + WSH | 265 | 0.23 | 1.17 | 0.16 | 0.75 | 0 (-0.18, 0.18) | 1 | 0.01 (-0.18, 0.19) | 1 | -0.02 (-0.2, 0.17) | 1 | 0 (-0.2, 0.2) | 1 |
| **Ln ΔIL-4/IL-10** |  |  |  |  |  |  |  |  |  |  |  |  |  |
| Control | 216 | -0.27 | 23.79 | 0.23 | 0.87 |  |  |  |  |  |  |  |  |
| Nutrition + WSH | 264 | 0.69 | 18.13 | 0.05 | 0.94 | -0.18 (-0.39, 0.03) | 0.18 | -0.18 (-0.4, 0.03) | 0.18 | -0.16 (-0.38, 0.06) | 0.32 | -0.18 (-0.39, 0.02) | 0.15 |
| **Ln ΔIL-5/IL-10** |  |  |  |  |  |  |  |  |  |  |  |  |  |
| Control | 216 | -0.04 | 0.83 | 0.12 | 0.93 |  |  |  |  |  |  |  |  |
| Nutrition + WSH | 265 | 0.11 | 1.24 | 0.05 | 1.04 | -0.07 (-0.26, 0.13) | 1 | -0.09 (-0.27, 0.1) | 0.73 | -0.15 (-0.32, 0.02) | 0.18 | -0.13 (-0.34, 0.08) | 0.48 |
| **Ln ΔIL-13/IL-10** |  |  |  |  |  |  |  |  |  |  |  |  |  |
| Control | 215 | 0.26 | 2.8 | 0.21 | 1.18 |  |  |  |  |  |  |  |  |
| Nutrition + WSH | 263 | 0.03 | 2.25 | -0.04 | 1.24 | -0.25 (-0.53, 0.04) | 0.17 | -0.25 (-0.53, 0.04) | 0.19 | -0.22 (-0.5, 0.05) | 0.22 | -0.22 (-0.57, 0.13) | 0.43 |
| **Ln ΔIL-17A/IL-10** |  |  |  |  |  |  |  |  |  |  |  |  |  |
| Control | 216 | -0.15 | 2.32 | 0.05 | 0.9 |  |  |  |  |  |  |  |  |
| Nutrition + WSH | 265 | 0.23 | 1.17 | 0.2 | 0.86 | 0.16 (-0.04, 0.35) | 0.23 | 0.17 (-0.02, 0.37) | 0.17 | 0.16 (-0.05, 0.37) | 0.29 | 0.25 (0.04, 0.47) | 0.05 |
| **Ln ΔIL-21/IL-10** |  |  |  |  |  |  |  |  |  |  |  |  |  |
| Control | 208 | 0.04 | 0.88 | 0.26 | 1.41 |  |  |  |  |  |  |  |  |
| Nutrition + WSH | 257 | 0.14 | 1.31 | 0.06 | 1.49 | -0.2 (-0.5, 0.11) | 0.41 | -0.19 (-0.48, 0.1) | 0.4 | -0.08 (-0.39, 0.22) | 1 | -0.06 (-0.36, 0.23) | 1 |
| **Ln ΔIL-2/IL-10** |  |  |  |  |  |  |  |  |  |  |  |  |  |
| Control | 214 | 0.02 | 0.66 | 0.37 | 1.4 |  |  |  |  |  |  |  |  |
| Nutrition + WSH | 254 | 0.2 | 3.49 | 0.25 | 1.66 | -0.12 (-0.47, 0.23) | 1 | -0.13 (-0.48, 0.22) | 0.94 | -0.1 (-0.45, 0.25) | 1 | -0.17 (-0.56, 0.22) | 0.8 |
| **Ln ΔGM-CSF/IL-10** |  |  |  |  |  |  |  |  |  |  |  |  |  |
| Control | 216 | 5.26 | 47.93 | 0.65 | 1.08 |  |  |  |  |  |  |  |  |
| Nutrition + WSH | 265 | 15.22 | 39.79 | 0.9 | 1.37 | 0.25 (0.04, 0.46) | 0.04 | 0.22 (-0.02, 0.45) | 0.15 | 0.08 (-0.13, 0.3) | 0.88 | 0.12 (-0.08, 0.33) | 0.48 |
| **Ln ΔIL-12/IL-4** |  |  |  |  |  |  |  |  |  |  |  |  |  |
| Control | 218 | 0 | 0.03 | -0.09 | 0.71 |  |  |  |  |  |  |  |  |
| Nutrition + WSH | 266 | 0.01 | 0.03 | 0.17 | 0.82 | 0.26 (0.09, 0.44) | <0.01 | 0.27 (0.08, 0.46) | 0.01 | 0.29 (0.11, 0.47) | <0.01 | 0.25 (0.03, 0.47) | 0.05 |
| **Ln ΔIFN-γ/IL-4** |  |  |  |  |  |  |  |  |  |  |  |  |  |
| Control | 218 | 0 | 0.13 | -0.06 | 0.7 |  |  |  |  |  |  |  |  |
| Nutrition + WSH | 267 | 0.02 | 0.11 | 0.11 | 0.71 | 0.17 (0.01, 0.32) | 0.07 | 0.18 (0.02, 0.33) | 0.06 | 0.16 (0, 0.32) | 0.09 | 0.18 (-0.01, 0.37) | 0.12 |
| **Ln ΔIL-12/IL-5** |  |  |  |  |  |  |  |  |  |  |  |  |  |
| Control | 218 | 0.31 | 3.52 | 0.02 | 0.84 |  |  |  |  |  |  |  |  |
| Nutrition + WSH | 267 | 0.56 | 3.75 | 0.16 | 1.13 | 0.15 (-0.05, 0.35) | 0.29 | 0.15 (-0.05, 0.35) | 0.26 | 0.22 (0.04, 0.4) | 0.04 | 0.3 (0.07, 0.53) | 0.02 |
| **Ln ΔIFN-γ/IL-5** |  |  |  |  |  |  |  |  |  |  |  |  |  |
| Control | 218 | 1.17 | 10.28 | 0.05 | 0.8 |  |  |  |  |  |  |  |  |
| Nutrition + WSH | 268 | 1.19 | 9.23 | 0.1 | 0.95 | 0.06 (-0.11, 0.23) | 1 | 0.06 (-0.11, 0.23) | 1 | 0.1 (-0.08, 0.27) | 0.56 | 0.18 (-0.03, 0.38) | 0.19 |
| **Ln ΔIL-12/IL-13** |  |  |  |  |  |  |  |  |  |  |  |  |  |
| Control | 216 | 0.09 | 1.41 | -0.08 | 1 |  |  |  |  |  |  |  |  |
| Nutrition + WSH | 265 | 0.08 | 2.8 | 0.27 | 1.18 | 0.35 (0.14, 0.57) | <0.01 | 0.37 (0.15, 0.59) | <0.01 | 0.35 (0.12, 0.58) | <0.01 | 0.38 (0.12, 0.64) | <0.01 |
| **Ln ΔIFN-γ/IL-13** |  |  |  |  |  |  |  |  |  |  |  |  |  |
| Control | 216 | 0.46 | 6.99 | -0.05 | 0.92 |  |  |  |  |  |  |  |  |
| Nutrition + WSH | 266 | 0.42 | 7.89 | 0.19 | 1.07 | 0.24 (0.03, 0.44) | 0.05 | 0.23 (0.03, 0.43) | 0.04 | 0.19 (0, 0.38) | 0.1 | 0.17 (-0.07, 0.41) | 0.35 |
| **Ln ΔIL-12/IL-17A** |  |  |  |  |  |  |  |  |  |  |  |  |  |
| Control | 218 | 0.11 | 0.78 | 0.08 | 0.75 |  |  |  |  |  |  |  |  |
| Nutrition + WSH | 267 | -0.04 | 0.6 | 0 | 0.8 | -0.08 (-0.23, 0.08) | 0.63 | -0.08 (-0.24, 0.08) | 0.62 | -0.1 (-0.25, 0.05) | 0.42 | -0.12 (-0.28, 0.04) | 0.27 |
| **Ln ΔIFN-γ/IL-17A** |  |  |  |  |  |  |  |  |  |  |  |  |  |
| Control | 218 | 0.4 | 3.76 | 0.11 | 0.64 |  |  |  |  |  |  |  |  |
| Nutrition + WSH | 268 | -0.3 | 5.61 | -0.06 | 0.68 | -0.17 (-0.32, -0.03) | 0.04 | -0.18 (-0.33, -0.03) | 0.03 | -0.16 (-0.31, -0.01) | 0.06 | -0.19 (-0.33, -0.04) | 0.02 |
| **Ln ΔIL-12/IL-21** |  |  |  |  |  |  |  |  |  |  |  |  |  |
| Control | 210 | -0.22 | 7.35 | -0.09 | 1.37 |  |  |  |  |  |  |  |  |
| Nutrition + WSH | 259 | 0.85 | 8.36 | 0.18 | 1.48 | 0.27 (-0.06, 0.6) | 0.23 | 0.27 (-0.06, 0.61) | 0.22 | 0.12 (-0.21, 0.46) | 0.94 | 0.08 (-0.3, 0.46) | 1 |
| **Ln ΔIFN-γ/IL-21** |  |  |  |  |  |  |  |  |  |  |  |  |  |
| Control | 210 | 0.25 | 24.1 | -0.06 | 1.29 |  |  |  |  |  |  |  |  |
| Nutrition + WSH | 260 | 1.99 | 28.58 | 0.11 | 1.38 | 0.17 (-0.12, 0.46) | 0.51 | 0.17 (-0.12, 0.47) | 0.5 | 0.05 (-0.29, 0.38) | 1 | 0.02 (-0.31, 0.36) | 1 |
| **Ln ΔPro-inflammatory cytokines^d^/IL-10** |  |  |  |  |  |  |  |  |  |  |  |  |  |
| Control | 216 |  |  | 0.59 | 0.78 |  |  |  |  |  |  |  |  |
| Nutrition + WSH | 260 |  |  | 0.52 | 0.88 | -0.06 (-0.22, 0.1) | 0.88 | -0.06 (-0.22, 0.1) | 0.95 | -0.07 (-0.23, 0.1) | 0.88 | -0.08 (-0.25, 0.1) | 0.79 |
| **Ln ΔTh1^e^/IL-10** |  |  |  |  |  |  |  |  |  |  |  |  |  |
| Control | 216 |  |  | 0.38 | 0.74 |  |  |  |  |  |  |  |  |
| Nutrition + WSH | 264 |  |  | 0.41 | 0.7 | 0.03 (-0.14, 0.2) | 1 | 0.03 (-0.14, 0.19) | 1 | 0.04 (-0.13, 0.21) | 1 | 0.01 (-0.15, 0.17) | 1 |
| **Ln ΔTh2^f^/IL-10** |  |  |  |  |  |  |  |  |  |  |  |  |  |
| Control | 215 |  |  | 0.69 | 0.82 |  |  |  |  |  |  |  |  |
| Nutrition + WSH | 262 |  |  | 0.53 | 0.84 | -0.15 (-0.32, 0.02) | 0.15 | -0.15 (-0.32, 0.02) | 0.17 | -0.19 (-0.36, -0.02) | 0.06 | -0.19 (-0.38, 0) | 0.09 |
| **Ln ΔTh17^g^/IL-10** |  |  |  |  |  |  |  |  |  |  |  |  |  |
| Control | 208 |  |  | 0.28 | 0.86 |  |  |  |  |  |  |  |  |
| Nutrition + WSH | 257 |  |  | 0.34 | 0.87 | 0.06 (-0.1, 0.22) | 0.91 | 0.07 (-0.08, 0.23) | 0.71 | 0.09 (-0.08, 0.26) | 0.62 | 0.08 (-0.08, 0.25) | 0.66 |
| **Ln ΔTh1^e^/Th2^f^** |  |  |  |  |  |  |  |  |  |  |  |  |  |
| Control | 216 |  |  | -0.31 | 0.54 |  |  |  |  |  |  |  |  |
| Nutrition + WSH | 264 |  |  | -0.13 | 0.62 | 0.18 (0.06, 0.29) | <0.01 | 0.17 (0.06, 0.29) | <0.01 | 0.18 (0.06, 0.31) | <0.01 | 0.17 (0.05, 0.3) | 0.01 |
| **Ln ΔTh1^e^/Th17^g^** |  |  |  |  |  |  |  |  |  |  |  |  |  |
| Control | 210 |  |  | 0.12 | 0.57 |  |  |  |  |  |  |  |  |
| Nutrition + WSH | 259 |  |  | 0.07 | 0.68 | -0.05 (-0.18, 0.08) | 0.89 | -0.05 (-0.18, 0.08) | 0.93 | -0.07 (-0.2, 0.06) | 0.59 | -0.09 (-0.22, 0.05) | 0.39 |

Confidence intervals were adjusted for clustered observations using robust standard errors.

IL = Interleukin; TNF-α = Tumor necrosis factor-α; CRP = C-reactive protein (CRP); IFN-γ = interferon-γ; GM-CSF = Granulocyte-macrophage colony-stimulating factor; AGP = Alpha-1-acid glycoprotein; IGF-1 = Insulin-like growth factor-1

^a^Adjusted for pre-specified covariates: Field staff who collected data, month of measurement, household food insecurity, child age, child sex, mother’s age, mother’s height, mother’s education level, number of children <18 years in the household, number of individuals living in the compound, distance in minutes to the primary water source, household floor materials, household wall materials, household electricity, and household assets (wardrobe, table, chair, clock, khat, chouki, radio, television, refrigerator, bicycle, motorcycle, sewing machine, mobile phone, cattle, goats, and chickens).

^b^Inverse probability of censoring weighting.

^c^P-values shown are Bonferroni corrected to control for familywise error rates

^d^Pro-inflammatory cytokines: IL-1β, IL-6, TNF-α

^e^Th1 cytokines: IL-12, IFN-γ

^f^Th2 cytokines: IL-4, IL-5, IL-13

^g^Th17 cytokines: IL-17A, IL-21

**Supplementary Table 7. Effect of intervention on change in individual immune status and growth factor measurements between ages 14 and 28 months**

|  |  |  |  |  |  | Unadjusted difference: Intervention vs. Control | | Age- and sex- adjusted difference: Intervention vs. Control | | Fully adjusted difference: Intervention vs. Control^a^ | | IPCW adjusted difference: Intervention vs. Control^b^ | |
| --- | --- | --- | --- | --- | --- | --- | --- | --- | --- | --- | --- | --- | --- |
| Outcome, Arm | N | Absolute Mean | Absolute SD | Mean | SD | 95% CI | P-value^c^ | 95% CI | P-value^c^ | 95% CI | P-value^c^ | 95% CI | P-value^c^ |
| **Ln ΔIL-1β (pg/ml)** |  |  |  |  |  |  |  |  |  |  |  |  |  |
| Control | 218 | 0.11 | 0.82 | 0.13 | 1.22 |  |  |  |  |  |  |  |  |
| Nutrition + WSH | 264 | -0.05 | 1.03 | -0.03 | 1.41 | -0.15 (-0.46, 0.15) | 0.66 | -0.09 (-0.38, 0.2) | 1 | -0.08 (-0.38, 0.22) | 1 | -0.11 (-0.41, 0.2) | 0.99 |
| **Ln ΔIL-6 (pg/ml)** |  |  |  |  |  |  |  |  |  |  |  |  |  |
| Control | 218 | 0.28 | 2.85 | 0.02 | 0.92 |  |  |  |  |  |  |  |  |
| Nutrition + WSH | 267 | -0.58 | 2.65 | -0.29 | 1.18 | -0.31 (-0.54, -0.08) | 0.02 | -0.26 (-0.51, -0.01) | 0.08 | -0.24 (-0.48, 0) | 0.1 | -0.21 (-0.48, 0.06) | 0.27 |
| **Ln ΔTNF-α (pg/ml)** |  |  |  |  |  |  |  |  |  |  |  |  |  |
| Control | 218 | -2.11 | 3.61 | -0.36 | 0.63 |  |  |  |  |  |  |  |  |
| Nutrition + WSH | 268 | -1.77 | 4.44 | -0.24 | 0.74 | 0.13 (-0.06, 0.31) | 0.36 | 0.11 (-0.09, 0.31) | 0.56 | 0.11 (-0.08, 0.31) | 0.52 | 0.12 (-0.07, 0.31) | 0.45 |
| **Ln ΔCRP (mg/L)** |  |  |  |  |  |  |  |  |  |  |  |  |  |
| Control | 196 | -2.24 | 17.04 | -0.1 | 1.91 |  |  |  |  |  |  |  |  |
| Nutrition + WSH | 248 | -0.5 | 13.65 | -0.3 | 1.88 | -0.2 (-0.53, 0.13) | 0.46 | -0.22 (-0.55, 0.12) | 0.41 | -0.13 (-0.49, 0.23) | 0.97 | 2.34 (-1.7, 6.37) | 0.51 |
| **Ln ΔIL-12 (pg/ml)** |  |  |  |  |  |  |  |  |  |  |  |  |  |
| Control | 218 | -0.15 | 1.53 | -0.09 | 0.76 |  |  |  |  |  |  |  |  |
| Nutrition + WSH | 267 | -0.21 | 1.58 | -0.05 | 0.84 | 0.04 (-0.16, 0.24) | 1 | 0.05 (-0.15, 0.25) | 1 | 0.07 (-0.12, 0.25) | 0.95 | -0.01 (-0.25, 0.23) | 1 |
| **Ln ΔIFN-γ (pg/ml)** |  |  |  |  |  |  |  |  |  |  |  |  |  |
| Control | 218 | 0.12 | 5.17 | -0.06 | 0.66 |  |  |  |  |  |  |  |  |
| Nutrition + WSH | 268 | -1.03 | 5.52 | -0.11 | 0.7 | -0.06 (-0.23, 0.12) | 1 | -0.05 (-0.22, 0.13) | 1 | -0.02 (-0.2, 0.17) | 1 | -0.02 (-0.29, 0.26) | 1 |
| **Ln ΔIL-4 (pg/ml)** |  |  |  |  |  |  |  |  |  |  |  |  |  |
| Control | 218 | 1.74 | 45.37 | 0 | 0.7 |  |  |  |  |  |  |  |  |
| Nutrition + WSH | 267 | -12.57 | 67.79 | -0.22 | 0.82 | -0.22 (-0.42, -0.03) | 0.05 | -0.21 (-0.42, 0) | 0.09 | -0.17 (-0.37, 0.04) | 0.22 | -0.25 (-0.49, 0) | 0.1 |
| **Ln ΔIL-5 (pg/ml)** |  |  |  |  |  |  |  |  |  |  |  |  |  |
| Control | 218 | -0.15 | 1.44 | -0.11 | 0.76 |  |  |  |  |  |  |  |  |
| Nutrition + WSH | 268 | -0.42 | 1.89 | -0.22 | 0.79 | -0.11 (-0.27, 0.05) | 0.32 | -0.11 (-0.26, 0.05) | 0.38 | -0.14 (-0.3, 0.02) | 0.19 | -0.14 (-0.32, 0.04) | 0.26 |
| **Ln ΔIL-13 (pg/ml)** |  |  |  |  |  |  |  |  |  |  |  |  |  |
| Control | 216 | 2.05 | 16.95 | -0.01 | 1.2 |  |  |  |  |  |  |  |  |
| Nutrition + WSH | 266 | -2.84 | 12.24 | -0.32 | 1.28 | -0.31 (-0.64, 0.02) | 0.13 | -0.3 (-0.62, 0.03) | 0.15 | -0.25 (-0.57, 0.08) | 0.28 | -0.24 (-0.65, 0.16) | 0.47 |
| **Ln ΔIL-17A (pg/ml)** |  |  |  |  |  |  |  |  |  |  |  |  |  |
| Control | 218 | -0.95 | 5.03 | -0.17 | 0.74 |  |  |  |  |  |  |  |  |
| Nutrition + WSH | 268 | -0.45 | 4.33 | -0.05 | 0.81 | 0.12 (-0.1, 0.34) | 0.59 | 0.13 (-0.09, 0.35) | 0.49 | 0.16 (-0.06, 0.38) | 0.29 | 0.09 (-0.13, 0.31) | 0.82 |
| **Ln ΔIL-21 (pg/ml)** |  |  |  |  |  |  |  |  |  |  |  |  |  |
| Control | 210 | -0.04 | 1.72 | 0.01 | 1.16 |  |  |  |  |  |  |  |  |
| Nutrition + WSH | 260 | -0.29 | 2.16 | -0.23 | 1.24 | -0.24 (-0.5, 0.02) | 0.13 | -0.24 (-0.5, 0.02) | 0.15 | -0.11 (-0.36, 0.15) | 0.83 | -0.1 (-0.37, 0.17) | 0.92 |
| **Ln ΔIL-10 (pg/ml)** |  |  |  |  |  |  |  |  |  |  |  |  |  |
| Control | 216 | -2.16 | 7.52 | -0.22 | 0.83 |  |  |  |  |  |  |  |  |
| Nutrition + WSH | 265 | -2.81 | 11.11 | -0.27 | 0.89 | -0.04 (-0.25, 0.16) | 1 | -0.05 (-0.25, 0.15) | 1 | -0.02 (-0.21, 0.17) | 1 | 0.01 (-0.2, 0.21) | 1 |
| **Ln ΔIL-2 (pg/ml)** |  |  |  |  |  |  |  |  |  |  |  |  |  |
| Control | 216 | 0.05 | 1.41 | 0.13 | 1.39 |  |  |  |  |  |  |  |  |
| Nutrition + WSH | 257 | 0.51 | 7.94 | -0.03 | 1.53 | -0.16 (-0.52, 0.19) | 0.73 | -0.17 (-0.51, 0.17) | 0.64 | -0.11 (-0.4, 0.18) | 0.92 | -0.2 (-0.62, 0.22) | 0.69 |
| **Ln ΔGM-CSF (pg/ml)** |  |  |  |  |  |  |  |  |  |  |  |  |  |
| Control | 218 | 48.57 | 249.62 | 0.42 | 0.92 |  |  |  |  |  |  |  |  |
| Nutrition + WSH | 268 | 97.86 | 272.83 | 0.63 | 1.09 | 0.2 (-0.02, 0.43) | 0.15 | 0.19 (-0.04, 0.42) | 0.2 | 0.11 (-0.12, 0.33) | 0.7 | 0.18 (-0.11, 0.48) | 0.45 |
| **Ln ΔAGP (g/L)** |  |  |  |  |  |  |  |  |  |  |  |  |  |
| Control | 70 | -0.23 | 0.74 | -0.26 | 0.61 |  |  |  |  |  |  |  |  |
| Nutrition + WSH | 138 | -0.14 | 0.81 | -0.21 | 0.62 | 0.06 (-0.12, 0.23) | 1 | 0.08 (-0.11, 0.26) | 0.84 | 0.02 (-0.17, 0.21) | 1 | 0.12 (-0.09, 0.34) | 0.54 |
| **Ln ΔIGF-1 (μg/L)** |  |  |  |  |  |  |  |  |  |  |  |  |  |
| Control | 209 | 12.64 | 21.47 | 0.33 | 0.55 |  |  |  |  |  |  |  |  |
| Nutrition + WSH | 240 | 10.97 | 20.97 | 0.31 | 0.54 | -0.02 (-0.11, 0.08) | 1 | -0.03 (-0.12, 0.07) | 1 | -0.06 (-0.16, 0.05) | 0.64 | -0.05 (-0.14, 0.04) | 0.6 |

Confidence intervals were adjusted for clustered observations using robust standard errors.

IL = Interleukin; TNF-α = Tumor necrosis factor-α; CRP = C-reactive protein (CRP); IFN-γ = interferon-γ; GM-CSF = Granulocyte-macrophage colony-stimulating factor; AGP = Alpha-1-acid glycoprotein; IGF-1 = Insulin-like growth factor-1

^a^Adjusted for pre-specified covariates: Field staff who collected data, month of measurement, household food insecurity, child age, child sex, mother’s age, mother’s height, mother’s education level, number of children <18 years in the household, number of individuals living in the compound, distance in minutes to the primary water source, household floor materials, household wall materials, household electricity, and household assets (wardrobe, table, chair, clock, khat, chouki, radio, television, refrigerator, bicycle, motorcycle, sewing machine, mobile phone, cattle, goats, and chickens).

^b^Inverse probability of censoring weighting.

^c^P-values shown are Bonferroni corrected to control for familywise error rates

**Supplementary Table 8. Effect of intervention on sum score of inflammation at ages 14 months and 28 months.**

|  |  | |  | |  | | Unadjusted difference: Intervention vs. Control | | Age- and sex- adjusted difference: Intervention vs. Control | | Fully adjusted difference: Intervention vs. Control^a^ | | IPCW adjusted difference: Intervention vs. Control^b^ | |
| --- | --- | --- | --- | --- | --- | --- | --- | --- | --- | --- | --- | --- | --- | --- |
| Outcome, Arm | | N | | Mean | | SD | 95% CI | P-value^c^ | 95% CI | P-value^c^ | 95% CI | P-value^c^ | 95% CI | P-value^c^ |
| **Sum score at 14 months** | |  | |  | |  |  |  |  |  |  |  |  |  |
| Control | | 285 | | -0.04 | | 0.93 |  |  |  |  |  |  |  |  |
| Nutrition + WSH | | 311 | | 0.04 | | 1.06 | 0.08 (-0.2, 0.35) | 1 | 0.07 (-0.21, 0.35) | 1 | 0.1 (-0.21, 0.4) | 1 | 0.05 (-0.23, 0.34) | 1 |
| **Sum score at 28 months** | |  | |  | |  |  |  |  |  |  |  |  |  |
| Control | | 313 | | 0.06 | | 0.97 |  |  |  |  |  |  |  |  |
| Nutrition + WSH | | 367 | | -0.05 | | 1.03 | -0.12 (-0.4, 0.17) | 0.84 | -0.12 (-0.4, 0.16) | 0.83 | -0.13 (-0.39, 0.13) | 0.66 | -0.07 (-0.33, 0.2) | 1 |

Confidence intervals were adjusted for clustered observations using robust standard errors.

^a^Adjusted for pre-specified covariates: Field staff who collected data, month of measurement, household food insecurity, child age, child sex, mother’s age, mother’s height, mother’s education level, number of children <18 years in the household, number of individuals living in the compound, distance in minutes to the primary water source, household floor materials, household wall materials, household electricity, and household assets (wardrobe, table, chair, clock, khat, chouki, radio, television, refrigerator, bicycle, motorcycle, sewing machine, mobile phone, cattle, goats, and chickens).

^b^Inverse probability of censoring weighting.

^c^P-values shown are Bonferroni corrected to control for familywise error rates

**Supplementary Table 9. Effect modification with child sex on individual immune status and growth factor measurements at age 14 months.**

|  | Female | | | | | Male | | | | | P-value for interaction^a^ |
| --- | --- | --- | --- | --- | --- | --- | --- | --- | --- | --- | --- |
| Outcome, Arm | N | Mean | SD | Unadjusted difference: Intervention vs. Control (95% CI) | P-value^a^ | N | Mean | SD | Unadjusted difference: Intervention vs. Control (95% CI) | P-value^a^ |  |
| **Ln IL-1β (pg/ml)** |  |  |  |  |  |  |  |  |  |  |  |
| Control | 143 | -0.2 | 1 |  |  | 142 | -0.32 | 1.03 |  |  |  |
| Nutrition + WSH | 164 | -0.16 | 1.15 | 0.04 (-0.29, 0.37) | 1 | 145 | -0.23 | 1.15 | 0.09 (-0.19, 0.38) | 1 | 1 |
| **Ln IL-6 (pg/ml)** |  |  |  |  |  |  |  |  |  |  |  |
| Control | 143 | 0.74 | 0.68 |  |  | 142 | 0.69 | 0.65 |  |  |  |
| Nutrition + WSH | 164 | 0.91 | 0.82 | 0.17 (-0.02, 0.36) | 0.17 | 145 | 0.81 | 0.83 | 0.12 (-0.06, 0.3) | 0.38 | 1 |
| **Ln TNF-α (pg/ml)** |  |  |  |  |  |  |  |  |  |  |  |
| Control | 143 | 1.83 | 0.48 |  |  | 142 | 1.83 | 0.51 |  |  |  |
| Nutrition + WSH | 164 | 1.79 | 0.6 | -0.04 (-0.22, 0.15) | 1 | 147 | 1.82 | 0.62 | -0.02 (-0.18, 0.15) | 1 | 1 |
| **Ln CRP (mg/L)** |  |  |  |  |  |  |  |  |  |  |  |
| Control | 126 | -0.16 | 1.63 |  |  | 127 | -0.04 | 1.64 |  |  |  |
| Nutrition + WSH | 152 | 0.4 | 1.7 | 0.56 (0.17, 0.94) | <0.01 | 138 | 0.13 | 1.39 | 0.17 (-0.16, 0.5) | 0.64 | 0.12 |
| **Ln IL-12 (pg/ml)** |  |  |  |  |  |  |  |  |  |  |  |
| Control | 143 | 1.03 | 0.44 |  |  | 142 | 0.93 | 0.59 |  |  |  |
| Nutrition + WSH | 163 | 0.93 | 0.79 | -0.1 (-0.27, 0.08) | 0.56 | 147 | 0.83 | 0.82 | -0.1 (-0.28, 0.08) | 0.58 | 1 |
| **Ln IFN-γ (pg/ml)** |  |  |  |  |  |  |  |  |  |  |  |
| Control | 143 | 2.06 | 0.43 |  |  | 142 | 2.03 | 0.46 |  |  |  |
| Nutrition + WSH | 164 | 2.05 | 0.6 | -0.01 (-0.17, 0.15) | 1 | 147 | 2 | 0.6 | -0.03 (-0.16, 0.1) | 1 | 1 |
| **Ln IL-4 (pg/ml)** |  |  |  |  |  |  |  |  |  |  |  |
| Control | 143 | 3.88 | 0.56 |  |  | 142 | 3.86 | 0.51 |  |  |  |
| Nutrition + WSH | 164 | 4 | 0.66 | 0.12 (-0.07, 0.31) | 0.45 | 146 | 3.91 | 0.68 | 0.06 (-0.12, 0.24) | 1 | 1 |
| **Ln IL-5 (pg/ml)** |  |  |  |  |  |  |  |  |  |  |  |
| Control | 143 | 0.5 | 0.7 |  |  | 142 | 0.47 | 0.65 |  |  |  |
| Nutrition + WSH | 164 | 0.69 | 0.54 | 0.18 (0.01, 0.36) | 0.09 | 147 | 0.63 | 0.67 | 0.16 (0.01, 0.3) | 0.06 | 1 |
| **Ln IL-13 (pg/ml)** |  |  |  |  |  |  |  |  |  |  |  |
| Control | 143 | 1.62 | 0.92 |  |  | 142 | 1.58 | 0.99 |  |  |  |
| Nutrition + WSH | 161 | 1.78 | 1.2 | 0.16 (-0.2, 0.53) | 0.77 | 147 | 1.67 | 1.11 | 0.09 (-0.16, 0.34) | 0.94 | 1 |
| **Ln IL-17A (pg/ml)** |  |  |  |  |  |  |  |  |  |  |  |
| Control | 143 | 1.65 | 0.55 |  |  | 142 | 1.57 | 0.56 |  |  |  |
| Nutrition + WSH | 164 | 1.6 | 0.72 | -0.05 (-0.22, 0.13) | 1 | 147 | 1.47 | 0.74 | -0.11 (-0.32, 0.1) | 0.61 | 1 |
| **Ln IL-21 (pg/ml)** |  |  |  |  |  |  |  |  |  |  |  |
| Control | 141 | 0.39 | 0.9 |  |  | 141 | 0.36 | 0.91 |  |  |  |
| Nutrition + WSH | 161 | 0.54 | 0.84 | 0.16 (-0.09, 0.4) | 0.42 | 144 | 0.37 | 0.88 | 0.01 (-0.24, 0.26) | 1 | 0.52 |
| **Ln IL-10 (pg/ml)** |  |  |  |  |  |  |  |  |  |  |  |
| Control | 143 | 2.23 | 0.73 |  |  | 141 | 2.24 | 0.64 |  |  |  |
| Nutrition + WSH | 163 | 2.4 | 0.67 | 0.17 (-0.03, 0.36) | 0.19 | 147 | 2.26 | 0.73 | 0.02 (-0.17, 0.2) | 1 | 0.41 |
| **Ln IL-2 (pg/ml)** |  |  |  |  |  |  |  |  |  |  |  |
| Control | 142 | -0.42 | 1.21 |  |  | 141 | -0.37 | 1.16 |  |  |  |
| Nutrition + WSH | 161 | -0.27 | 1.18 | 0.15 (-0.18, 0.47) | 0.75 | 143 | -0.36 | 1.2 | 0.01 (-0.34, 0.37) | 1 | 0.98 |
| **Ln GM-CSF (pg/ml)** |  |  |  |  |  |  |  |  |  |  |  |
| Control | 143 | 4.15 | 0.87 |  |  | 142 | 4.09 | 1 |  |  |  |
| Nutrition + WSH | 164 | 4.15 | 0.88 | 0 (-0.21, 0.21) | 1 | 147 | 3.96 | 0.92 | -0.14 (-0.38, 0.11) | 0.54 | 0.68 |
| **Ln AGP (g/L)** |  |  |  |  |  |  |  |  |  |  |  |
| Control | 123 | -0.04 | 0.46 |  |  | 130 | 0 | 0.46 |  |  |  |
| Nutrition + WSH | 149 | 0.08 | 0.45 | 0.12 (0, 0.24) | 0.1 | 128 | -0.05 | 0.41 | -0.05 (-0.17, 0.06) | 0.72 | 0.01 |
| **Ln IGF-1 (μg/L)** |  |  |  |  |  |  |  |  |  |  |  |
| Control | 123 | 3.65 | 0.59 |  |  | 129 | 3.45 | 0.62 |  |  |  |
| Nutrition + WSH | 149 | 3.47 | 0.66 | -0.19 (-0.35, -0.03) | 0.04 | 128 | 3.41 | 0.52 | -0.04 (-0.19, 0.11) | 1 | 0.37 |

Confidence intervals were adjusted for clustered observations using robust standard errors.

IL = Interleukin; TNF-α = Tumor necrosis factor-α; CRP = C-reactive protein (CRP); IFN-γ = interferon-γ; GM-CSF = Granulocyte-macrophage colony-stimulating factor; AGP = Alpha-1-acid glycoprotein; IGF-1 = Insulin-like growth factor-1

^a^P-values shown are Bonferroni corrected to control for familywise error rates

**Supplementary Table 10. Effect modification with child sex on cytokine ratios at age 14 months.**

|  | Female | | | | | Male | | | | | P-value for interaction^a^ |
| --- | --- | --- | --- | --- | --- | --- | --- | --- | --- | --- | --- |
| Outcome | N | Mean | SD | Unadjusted difference: Intervention vs. Control (95% CI) | P-value^a^ | N | Mean | SD | Unadjusted difference: Intervention vs. Control (95% CI) | P-value^a^ |  |
| **Ln IL-1β/IL-10** |  |  |  |  |  |  |  |  |  |  |  |
| Control | 143 | -2.43 | 0.9 |  |  | 141 | -2.56 | 0.89 |  |  |  |
| Nutrition + WSH | 163 | -2.55 | 1.05 | -0.12 (-0.35, 0.12) | 0.67 | 145 | -2.48 | 1.1 | 0.08 (-0.16, 0.32) | 1 | 0.12 |
| **Ln IL-6/IL-10** |  |  |  |  |  |  |  |  |  |  |  |
| Control | 143 | -1.49 | 0.8 |  |  | 141 | -1.55 | 0.76 |  |  |  |
| Nutrition + WSH | 163 | -1.49 | 0.92 | 0 (-0.2, 0.2) | 1 | 145 | -1.44 | 0.96 | 0.11 (-0.12, 0.33) | 0.7 | 0.77 |
| **Ln TNF-α/IL-10** |  |  |  |  |  |  |  |  |  |  |  |
| Control | 143 | -0.4 | 0.72 |  |  | 141 | -0.41 | 0.66 |  |  |  |
| Nutrition + WSH | 163 | -0.61 | 0.67 | -0.2 (-0.36, -0.04) | 0.03 | 147 | -0.44 | 0.65 | -0.03 (-0.17, 0.1) | 1 | 0.22 |
| **Ln IL-12/IL-10** |  |  |  |  |  |  |  |  |  |  |  |
| Control | 143 | -1.2 | 0.58 |  |  | 141 | -1.29 | 0.54 |  |  |  |
| Nutrition + WSH | 162 | -1.45 | 0.68 | -0.25 (-0.39, -0.1) | <0.01 | 147 | -1.43 | 0.72 | -0.14 (-0.3, 0.02) | 0.16 | 0.57 |
| **Ln IFN-γ/IL-10** |  |  |  |  |  |  |  |  |  |  |  |
| Control | 143 | -0.17 | 0.6 |  |  | 141 | -0.21 | 0.53 |  |  |  |
| Nutrition + WSH | 163 | -0.33 | 0.57 | -0.17 (-0.29, -0.04) | 0.02 | 147 | -0.26 | 0.63 | -0.04 (-0.19, 0.11) | 1 | 0.31 |
| **Ln IL-4/IL-10** |  |  |  |  |  |  |  |  |  |  |  |
| Control | 143 | 1.65 | 0.72 |  |  | 141 | 1.62 | 0.67 |  |  |  |
| Nutrition + WSH | 163 | 1.62 | 0.7 | -0.04 (-0.21, 0.14) | 1 | 146 | 1.66 | 0.69 | 0.04 (-0.13, 0.21) | 1 | 0.95 |
| **Ln IL-5/IL-10** |  |  |  |  |  |  |  |  |  |  |  |
| Control | 143 | -1.73 | 0.8 |  |  | 141 | -1.76 | 0.7 |  |  |  |
| Nutrition + WSH | 163 | -1.71 | 0.71 | 0.02 (-0.16, 0.2) | 1 | 147 | -1.63 | 0.82 | 0.13 (-0.01, 0.28) | 0.14 | 0.51 |
| **Ln IL-13/IL-10** |  |  |  |  |  |  |  |  |  |  |  |
| Control | 143 | -0.61 | 0.93 |  |  | 141 | -0.66 | 0.95 |  |  |  |
| Nutrition + WSH | 161 | -0.63 | 1.05 | -0.03 (-0.3, 0.25) | 1 | 147 | -0.58 | 0.94 | 0.08 (-0.16, 0.31) | 1 | 1 |
| **Ln IL-17A/IL-10** |  |  |  |  |  |  |  |  |  |  |  |
| Control | 143 | -0.58 | 0.7 |  |  | 141 | -0.66 | 0.55 |  |  |  |
| Nutrition + WSH | 163 | -0.77 | 0.62 | -0.19 (-0.38, -0.01) | 0.07 | 147 | -0.79 | 0.73 | -0.13 (-0.27, 0.02) | 0.16 | 0.96 |
| **Ln IL-21/IL-10** |  |  |  |  |  |  |  |  |  |  |  |
| Control | 141 | -1.84 | 1.04 |  |  | 140 | -1.88 | 1.06 |  |  |  |
| Nutrition + WSH | 160 | -1.85 | 0.99 | -0.01 (-0.27, 0.25) | 1 | 144 | -1.89 | 1.04 | -0.01 (-0.26, 0.24) | 1 | 1 |
| **Ln IL-2/IL-10** |  |  |  |  |  |  |  |  |  |  |  |
| Control | 142 | -2.68 | 1.22 |  |  | 140 | -2.62 | 1.16 |  |  |  |
| Nutrition + WSH | 160 | -2.67 | 1.3 | 0.01 (-0.3, 0.31) | 1 | 143 | -2.62 | 1.2 | -0.01 (-0.33, 0.32) | 1 | 1 |
| **Ln GM-CSF/IL-10** |  |  |  |  |  |  |  |  |  |  |  |
| Control | 143 | -2.43 | 0.9 |  |  | 141 | -2.56 | 0.89 |  |  |  |
| Nutrition + WSH | 163 | -2.55 | 1.05 | -0.16 (-0.38, 0.06) | 0.3 | 145 | -2.48 | 1.1 | -0.16 (-0.36, 0.05) | 0.26 | 1 |
| **Ln IL-12/IL-4** |  |  |  |  |  |  |  |  |  |  |  |
| Control | 143 | -2.86 | 0.43 |  |  | 142 | -2.93 | 0.56 |  |  |  |
| Nutrition + WSH | 163 | -3.07 | 0.68 | -0.21 (-0.37, -0.06) | 0.01 | 146 | -3.09 | 0.65 | -0.16 (-0.3, -0.01) | 0.06 | 1 |
| **Ln IFN-γ/IL-4** |  |  |  |  |  |  |  |  |  |  |  |
| Control | 143 | -1.82 | 0.44 |  |  | 142 | -1.83 | 0.45 |  |  |  |
| Nutrition + WSH | 164 | -1.95 | 0.54 | -0.13 (-0.26, 0) | 0.12 | 146 | -1.91 | 0.56 | -0.08 (-0.19, 0.03) | 0.29 | 1 |
| **Ln IL-12/IL-5** |  |  |  |  |  |  |  |  |  |  |  |
| Control | 143 | 0.53 | 0.56 |  |  | 142 | 0.45 | 0.73 |  |  |  |
| Nutrition + WSH | 163 | 0.25 | 0.77 | -0.28 (-0.45, -0.1) | <0.01 | 147 | 0.2 | 0.77 | -0.25 (-0.43, -0.08) | <0.01 | 1 |
| **Ln IFN-γ/IL-5** |  |  |  |  |  |  |  |  |  |  |  |
| Control | 143 | 1.56 | 0.6 |  |  | 142 | 1.56 | 0.63 |  |  |  |
| Nutrition + WSH | 164 | 1.37 | 0.64 | -0.19 (-0.36, -0.03) | 0.04 | 147 | 1.37 | 0.72 | -0.18 (-0.33, -0.03) | 0.03 | 1 |
| **Ln IL-12/IL-13** |  |  |  |  |  |  |  |  |  |  |  |
| Control | 143 | -0.59 | 0.75 |  |  | 142 | -0.66 | 0.88 |  |  |  |
| Nutrition + WSH | 160 | -0.85 | 0.94 | -0.26 (-0.5, -0.01) | 0.08 | 147 | -0.85 | 0.97 | -0.19 (-0.41, 0.03) | 0.19 | 1 |
| **Ln IFN-γ/IL-13** |  |  |  |  |  |  |  |  |  |  |  |
| Control | 143 | 0.44 | 0.74 |  |  | 142 | 0.44 | 0.81 |  |  |  |
| Nutrition + WSH | 161 | 0.28 | 0.93 | -0.16 (-0.4, 0.08) | 0.36 | 147 | 0.33 | 0.91 | -0.12 (-0.31, 0.08) | 0.47 | 1 |
| **Ln IL-12/IL-17A** |  |  |  |  |  |  |  |  |  |  |  |
| Control | 143 | -0.62 | 0.42 |  |  | 142 | -0.65 | 0.49 |  |  |  |
| Nutrition + WSH | 163 | -0.67 | 0.55 | -0.05 (-0.16, 0.06) | 0.71 | 147 | -0.64 | 0.65 | 0.01 (-0.13, 0.15) | 1 | 0.82 |
| **Ln IFN-γ/IL-17A** |  |  |  |  |  |  |  |  |  |  |  |
| Control | 143 | 0.41 | 0.5 |  |  | 142 | 0.45 | 0.46 |  |  |  |
| Nutrition + WSH | 164 | 0.45 | 0.46 | 0.04 (-0.08, 0.15) | 1 | 147 | 0.53 | 0.58 | 0.08 (-0.09, 0.25) | 0.73 | 1 |
| **Ln IL-12/IL-21** |  |  |  |  |  |  |  |  |  |  |  |
| Control | 141 | 0.64 | 0.89 |  |  | 141 | 0.57 | 1.02 |  |  |  |
| Nutrition + WSH | 160 | 0.38 | 1.01 | -0.26 (-0.53, 0.02) | 0.13 | 144 | 0.46 | 1.15 | -0.11 (-0.37, 0.15) | 0.8 | 0.7 |
| **Ln IFN-γ/IL-21** |  |  |  |  |  |  |  |  |  |  |  |
| Control | 141 | 1.67 | 0.9 |  |  | 141 | 1.67 | 0.93 |  |  |  |
| Nutrition + WSH | 161 | 1.51 | 0.87 | -0.16 (-0.41, 0.09) | 0.43 | 144 | 1.64 | 0.96 | -0.03 (-0.25, 0.2) | 1 | 0.69 |
| **Ln Pro-inflammatory cytokines^b^/IL-10** |  |  |  |  |  |  |  |  |  |  |  |
| Control | 143 | 1.31 | 0.67 |  |  | 141 | 1.27 | 0.6 |  |  |  |
| Nutrition + WSH | 163 | 1.25 | 0.61 | -0.06 (-0.2, 0.08) | 0.84 | 143 | 1.35 | 0.7 | 0.08 (-0.08, 0.24) | 0.62 | 0.22 |
| **Ln Th1^c^/IL-10** |  |  |  |  |  |  |  |  |  |  |  |
| Control | 143 | 1.21 | 0.57 |  |  | 141 | 1.14 | 0.51 |  |  |  |
| Nutrition + WSH | 162 | 1.02 | 0.52 | -0.19 (-0.32, -0.07) | <0.01 | 147 | 1.08 | 0.58 | -0.06 (-0.2, 0.08) | 0.76 | 0.26 |
| **Ln Th2^d^/IL-10** |  |  |  |  |  |  |  |  |  |  |  |
| Control | 143 | 0.87 | 0.7 |  |  | 141 | 0.84 | 0.63 |  |  |  |
| Nutrition + WSH | 161 | 0.86 | 0.61 | -0.01 (-0.15, 0.14) | 1 | 146 | 0.95 | 0.67 | 0.1 (-0.04, 0.25) | 0.3 | 0.54 |
| **Ln Th17^e^/IL-10** |  |  |  |  |  |  |  |  |  |  |  |
| Control | 141 | 0.79 | 0.74 |  |  | 140 | 0.73 | 0.65 |  |  |  |
| Nutrition + WSH | 160 | 0.68 | 0.67 | -0.12 (-0.29, 0.06) | 0.39 | 144 | 0.67 | 0.69 | -0.06 (-0.2, 0.08) | 0.8 | 1 |
| **Ln Th1^c^/Th2^d^** |  |  |  |  |  |  |  |  |  |  |  |
| Control | 143 | 0.34 | 0.38 |  |  | 142 | 0.3 | 0.43 |  |  |  |
| Nutrition + WSH | 160 | 0.14 | 0.48 | -0.2 (-0.3, -0.1) | <0.001 | 146 | 0.14 | 0.47 | -0.16 (-0.27, -0.06) | <0.01 | 1 |
| **Ln Th1^c^/Th17^e^** |  |  |  |  |  |  |  |  |  |  |  |
| Control | 141 | 0.42 | 0.47 |  |  | 141 | 0.41 | 0.48 |  |  |  |
| Nutrition + WSH | 160 | 0.33 | 0.51 | -0.08 (-0.22, 0.06) | 0.48 | 144 | 0.41 | 0.52 | 0 (-0.13, 0.12) | 1 | 0.66 |

Confidence intervals were adjusted for clustered observations using robust standard errors.

IL = Interleukin; TNF-α = Tumor necrosis factor-α; CRP = C-reactive protein (CRP); IFN-γ = interferon-γ; GM-CSF = Granulocyte-macrophage colony-stimulating factor; AGP = Alpha-1-acid glycoprotein; IGF-1 = Insulin-like growth factor-1

^a^P-values shown are Bonferroni corrected to control for familywise error rates

^b^Pro-inflammatory cytokines: IL-1β, IL-6, TNF-α

^c^Th1 cytokines: IL-12, IFN-γ

^d^Th2 cytokines: IL-4, IL-5, IL-13

^e^Th17 cytokines: IL-17A, IL-21

**Supplementary Table 11. Effect modification with child sex on individual immune status and growth factor measurements at age 28 months.**

|  | Female | | | | | Male | | | | | P-value for interaction^a^ |
| --- | --- | --- | --- | --- | --- | --- | --- | --- | --- | --- | --- |
| Outcome, Arm | N | Mean | SD | Unadjusted difference: Intervention vs. Control (95% CI) | P-value^a^ | N | Mean | SD | Unadjusted difference: Intervention vs. Control (95% CI) | P-value^a^ |  |
| **Ln IL-1β (pg/ml)** |  |  |  |  |  |  |  |  |  |  |  |
| Control | 159 | -0.04 | 0.88 |  |  | 154 | -0.2 | 1.08 |  |  |  |
| Nutrition + WSH | 189 | -0.16 | 1.14 | -0.12 (-0.43, 0.18) | 0.86 | 176 | -0.32 | 1.15 | -0.11 (-0.42, 0.2) | 0.96 | 1 |
| **Ln IL-6 (pg/ml)** |  |  |  |  |  |  |  |  |  |  |  |
| Control | 159 | 0.74 | 0.75 |  |  | 154 | 0.78 | 0.78 |  |  |  |
| Nutrition + WSH | 190 | 0.56 | 0.93 | -0.17 (-0.41, 0.07) | 0.32 | 176 | 0.52 | 1 | -0.26 (-0.52, 0.01) | 0.12 | 0.91 |
| **Ln TNF-α (pg/ml)** |  |  |  |  |  |  |  |  |  |  |  |
| Control | 159 | 1.51 | 0.49 |  |  | 154 | 1.46 | 0.64 |  |  |  |
| Nutrition + WSH | 190 | 1.56 | 0.49 | 0.05 (-0.11, 0.22) | 1 | 177 | 1.52 | 0.57 | 0.07 (-0.09, 0.23) | 0.83 | 1 |
| **Ln CRP (mg/L)** |  |  |  |  |  |  |  |  |  |  |  |
| Control | 155 | -0.14 | 1.26 |  |  | 155 | -0.18 | 1.25 |  |  |  |
| Nutrition + WSH | 197 | 0 | 1.32 | 0.15 (-0.12, 0.42) | 0.57 | 175 | -0.23 | 1.5 | -0.06 (-0.38, 0.27) | 1 | 0.71 |
| **Ln IL-12 (pg/ml)** |  |  |  |  |  |  |  |  |  |  |  |
| Control | 159 | 0.98 | 0.58 |  |  | 154 | 0.8 | 0.69 |  |  |  |
| Nutrition + WSH | 190 | 0.87 | 0.69 | -0.1 (-0.28, 0.08) | 0.54 | 177 | 0.77 | 0.7 | -0.03 (-0.23, 0.17) | 1 | 0.81 |
| **Ln IFN-γ (pg/ml)** |  |  |  |  |  |  |  |  |  |  |  |
| Control | 159 | 2.07 | 0.59 |  |  | 154 | 1.94 | 0.69 |  |  |  |
| Nutrition + WSH | 190 | 1.96 | 0.64 | -0.11 (-0.28, 0.07) | 0.47 | 177 | 1.84 | 0.58 | -0.1 (-0.28, 0.08) | 0.55 | 1 |
| **Ln IL-4 (pg/ml)** |  |  |  |  |  |  |  |  |  |  |  |
| Control | 159 | 3.92 | 0.54 |  |  | 154 | 3.87 | 0.62 |  |  |  |
| Nutrition + WSH | 190 | 3.81 | 0.65 | -0.11 (-0.26, 0.05) | 0.34 | 177 | 3.65 | 0.68 | -0.22 (-0.41, -0.03) | 0.05 | 0.25 |
| **Ln IL-5 (pg/ml)** |  |  |  |  |  |  |  |  |  |  |  |
| Control | 159 | 0.37 | 0.69 |  |  | 154 | 0.4 | 0.89 |  |  |  |
| Nutrition + WSH | 190 | 0.49 | 0.57 | 0.12 (-0.03, 0.26) | 0.23 | 177 | 0.29 | 0.8 | -0.11 (-0.34, 0.11) | 0.65 | 0.06 |
| **Ln IL-13 (pg/ml)** |  |  |  |  |  |  |  |  |  |  |  |
| Control | 158 | 1.67 | 1.02 |  |  | 153 | 1.49 | 1.29 |  |  |  |
| Nutrition + WSH | 189 | 1.58 | 1.01 | -0.1 (-0.39, 0.2) | 1 | 177 | 1.28 | 1.22 | -0.21 (-0.62, 0.19) | 0.61 | 0.86 |
| **Ln IL-17A (pg/ml)** |  |  |  |  |  |  |  |  |  |  |  |
| Control | 159 | 1.47 | 0.62 |  |  | 154 | 1.43 | 0.64 |  |  |  |
| Nutrition + WSH | 190 | 1.53 | 0.65 | 0.07 (-0.11, 0.24) | 0.88 | 177 | 1.33 | 0.64 | -0.11 (-0.31, 0.1) | 0.63 | 0.07 |
| **Ln IL-21 (pg/ml)** |  |  |  |  |  |  |  |  |  |  |  |
| Control | 156 | 0.39 | 0.88 |  |  | 152 | 0.34 | 0.84 |  |  |  |
| Nutrition + WSH | 186 | 0.32 | 0.93 | -0.08 (-0.31, 0.16) | 1 | 176 | 0.17 | 1 | -0.18 (-0.4, 0.05) | 0.24 | 0.96 |
| **Ln IL-10 (pg/ml)** |  |  |  |  |  |  |  |  |  |  |  |
| Control | 158 | 2.1 | 0.66 |  |  | 154 | 1.94 | 0.68 |  |  |  |
| Nutrition + WSH | 190 | 2.2 | 0.69 | 0.11 (-0.07, 0.28) | 0.49 | 173 | 1.96 | 0.85 | 0.02 (-0.16, 0.2) | 1 | 0.79 |
| **Ln IL-2 (pg/ml)** |  |  |  |  |  |  |  |  |  |  |  |
| Control | 157 | -0.23 | 1.09 |  |  | 153 | -0.23 | 1.08 |  |  |  |
| Nutrition + WSH | 188 | -0.29 | 1.2 | -0.06 (-0.35, 0.22) | 1 | 173 | -0.42 | 1.27 | -0.19 (-0.54, 0.16) | 0.59 | 1 |
| **Ln GM-CSF (pg/ml)** |  |  |  |  |  |  |  |  |  |  |  |
| Control | 159 | 4.58 | 0.9 |  |  | 154 | 4.5 | 1.1 |  |  |  |
| Nutrition + WSH | 190 | 4.73 | 1.08 | 0.15 (-0.11, 0.41) | 0.49 | 177 | 4.51 | 1.05 | 0.01 (-0.26, 0.27) | 1 | 0.57 |
| **Ln AGP (g/L)** |  |  |  |  |  |  |  |  |  |  |  |
| Control | 60 | -0.2 | 0.5 |  |  | 48 | -0.28 | 0.46 |  |  |  |
| Nutrition + WSH | 129 | -0.15 | 0.5 | 0.05 (-0.1, 0.19) | 1 | 114 | -0.21 | 0.53 | 0.07 (-0.04, 0.19) | 0.43 | 1 |
| **Ln IGF-1 (μg/L)** |  |  |  |  |  |  |  |  |  |  |  |
| Control | 165 | 3.97 | 0.46 |  |  | 160 | 3.74 | 0.47 |  |  |  |
| Nutrition + WSH | 200 | 3.88 | 0.52 | -0.09 (-0.19, 0.02) | 0.2 | 179 | 3.71 | 0.45 | -0.03 (-0.14, 0.08) | 1 | 0.91 |

Confidence intervals were adjusted for clustered observations using robust standard errors.

IL = Interleukin; TNF-α = Tumor necrosis factor-α; CRP = C-reactive protein (CRP); IFN-γ = interferon-γ; GM-CSF = Granulocyte-macrophage colony-stimulating factor; AGP = Alpha-1-acid glycoprotein; IGF-1 = Insulin-like growth factor-1

^a^P-values shown are Bonferroni corrected to control for familywise error rates

**Supplementary Table 12. Effect modification with child sex on cytokine ratios at age 28 months.**

|  | Female | | | | | Male | | | | | P-value for interaction^a^ |
| --- | --- | --- | --- | --- | --- | --- | --- | --- | --- | --- | --- |
| Outcome | N | Mean | SD | Unadjusted difference: Intervention vs. Control (95% CI) | P-value^a^ | N | Mean | SD | Unadjusted difference: Intervention vs. Control (95% CI) | P-value^a^ |  |
| **Ln IL-1β/IL-10** |  |  |  |  |  |  |  |  |  |  |  |
| Control | 158 | -2.13 | 0.82 |  |  | 154 | -2.15 | 0.89 |  |  |  |
| Nutrition + WSH | 189 | -2.36 | 1.04 | -0.23 (-0.5, 0.04) | 0.18 | 172 | -2.32 | 1.08 | -0.17 (-0.43, 0.09) | 0.42 | 1 |
| **Ln IL-6/IL-10** |  |  |  |  |  |  |  |  |  |  |  |
| Control | 158 | -1.35 | 0.82 |  |  | 154 | -1.17 | 0.81 |  |  |  |
| Nutrition + WSH | 190 | -1.64 | 0.98 | -0.29 (-0.53, -0.05) | 0.04 | 172 | -1.42 | 1.07 | -0.25 (-0.5, 0) | 0.1 | 1 |
| **Ln TNF-α/IL-10** |  |  |  |  |  |  |  |  |  |  |  |
| Control | 158 | -0.58 | 0.58 |  |  | 154 | -0.49 | 0.72 |  |  |  |
| Nutrition + WSH | 190 | -0.64 | 0.69 | -0.05 (-0.21, 0.1) | 0.96 | 173 | -0.43 | 0.84 | 0.05 (-0.13, 0.24) | 1 | 0.82 |
| **Ln IL-12/IL-10** |  |  |  |  |  |  |  |  |  |  |  |
| Control | 158 | -1.12 | 0.51 |  |  | 154 | -1.15 | 0.65 |  |  |  |
| Nutrition + WSH | 190 | -1.33 | 0.59 | -0.21 (-0.34, -0.09) | <0.01 | 173 | -1.17 | 0.71 | -0.02 (-0.16, 0.12) | 1 | 0.07 |
| **Ln IFN-γ/IL-10** |  |  |  |  |  |  |  |  |  |  |  |
| Control | 158 | -0.02 | 0.5 |  |  | 154 | 0 | 0.63 |  |  |  |
| Nutrition + WSH | 190 | -0.24 | 0.57 | -0.22 (-0.34, -0.1) | <0.001 | 173 | -0.1 | 0.66 | -0.1 (-0.25, 0.05) | 0.37 | 0.43 |
| **Ln IL-4/IL-10** |  |  |  |  |  |  |  |  |  |  |  |
| Control | 158 | 1.83 | 0.65 |  |  | 154 | 1.93 | 0.63 |  |  |  |
| Nutrition + WSH | 190 | 1.61 | 0.69 | -0.22 (-0.37, -0.06) | 0.01 | 173 | 1.7 | 0.74 | -0.23 (-0.41, -0.05) | 0.03 | 1 |
| **Ln IL-5/IL-10** |  |  |  |  |  |  |  |  |  |  |  |
| Control | 158 | -1.72 | 0.69 |  |  | 154 | -1.54 | 0.87 |  |  |  |
| Nutrition + WSH | 190 | -1.71 | 0.77 | 0.01 (-0.16, 0.17) | 1 | 173 | -1.66 | 0.91 | -0.11 (-0.32, 0.1) | 0.59 | 0.72 |
| **Ln IL-13/IL-10** |  |  |  |  |  |  |  |  |  |  |  |
| Control | 158 | -0.42 | 0.97 |  |  | 153 | -0.46 | 1.11 |  |  |  |
| Nutrition + WSH | 189 | -0.62 | 0.9 | -0.2 (-0.44, 0.04) | 0.2 | 173 | -0.69 | 1.05 | -0.23 (-0.56, 0.1) | 0.33 | 1 |
| **Ln IL-17A/IL-10** |  |  |  |  |  |  |  |  |  |  |  |
| Control | 158 | -0.63 | 0.6 |  |  | 154 | -0.51 | 0.61 |  |  |  |
| Nutrition + WSH | 190 | -0.67 | 0.69 | -0.04 (-0.18, 0.1) | 1 | 173 | -0.62 | 0.69 | -0.11 (-0.27, 0.04) | 0.3 | 0.92 |
| **Ln IL-21/IL-10** |  |  |  |  |  |  |  |  |  |  |  |
| Control | 155 | -1.7 | 1.04 |  |  | 152 | -1.6 | 1.09 |  |  |  |
| Nutrition + WSH | 186 | -1.87 | 1.11 | -0.18 (-0.43, 0.08) | 0.35 | 172 | -1.79 | 1.32 | -0.19 (-0.49, 0.11) | 0.42 | 1 |
| **Ln IL-2/IL-10** |  |  |  |  |  |  |  |  |  |  |  |
| Control | 156 | -2.33 | 1.02 |  |  | 153 | -2.18 | 1.16 |  |  |  |
| Nutrition + WSH | 188 | -2.49 | 1.28 | -0.16 (-0.47, 0.14) | 0.6 | 169 | -2.38 | 1.43 | -0.2 (-0.56, 0.15) | 0.53 | 1 |
| **Ln GM-CSF/IL-10** |  |  |  |  |  |  |  |  |  |  |  |
| Control | 158 | 2.5 | 0.92 |  |  | 154 | 2.56 | 0.91 |  |  |  |
| Nutrition + WSH | 190 | 2.53 | 0.97 | 0.03 (-0.19, 0.25) | 1 | 173 | 2.57 | 1.08 | 0 (-0.22, 0.23) | 1 | 1 |
| **Ln IL-12/IL-4** |  |  |  |  |  |  |  |  |  |  |  |
| Control | 159 | -2.94 | 0.5 |  |  | 154 | -3.07 | 0.54 |  |  |  |
| Nutrition + WSH | 190 | -2.94 | 0.6 | 0.01 (-0.12, 0.14) | 1 | 177 | -2.88 | 0.53 | 0.19 (0.04, 0.34) | 0.02 | 0.06 |
| **Ln IFN-γ/IL-4** |  |  |  |  |  |  |  |  |  |  |  |
| Control | 159 | -1.85 | 0.53 |  |  | 154 | -1.93 | 0.63 |  |  |  |
| Nutrition + WSH | 190 | -1.85 | 0.61 | 0 (-0.13, 0.13) | 1 | 177 | -1.81 | 0.44 | 0.12 (-0.01, 0.25) | 0.16 | 0.3 |
| **Ln IL-12/IL-5** |  |  |  |  |  |  |  |  |  |  |  |
| Control | 159 | 0.6 | 0.65 |  |  | 154 | 0.4 | 0.97 |  |  |  |
| Nutrition + WSH | 190 | 0.38 | 0.76 | -0.22 (-0.4, -0.04) | 0.03 | 177 | 0.48 | 0.86 | 0.08 (-0.12, 0.29) | 0.85 | 0.01 |
| **Ln IFN-γ/IL-5** |  |  |  |  |  |  |  |  |  |  |  |
| Control | 159 | 1.69 | 0.66 |  |  | 154 | 1.54 | 1.02 |  |  |  |
| Nutrition + WSH | 190 | 1.47 | 0.74 | -0.22 (-0.39, -0.06) | 0.02 | 177 | 1.56 | 0.82 | 0.01 (-0.2, 0.23) | 1 | 0.09 |
| **Ln IL-12/IL-13** |  |  |  |  |  |  |  |  |  |  |  |
| Control | 158 | -0.69 | 0.83 |  |  | 153 | -0.7 | 1.03 |  |  |  |
| Nutrition + WSH | 189 | -0.7 | 0.79 | -0.01 (-0.2, 0.18) | 1 | 177 | -0.51 | 1.03 | 0.18 (-0.09, 0.46) | 0.38 | 0.29 |
| **Ln IFN-γ/IL-13** |  |  |  |  |  |  |  |  |  |  |  |
| Control | 158 | 0.4 | 0.82 |  |  | 153 | 0.45 | 1.01 |  |  |  |
| Nutrition + WSH | 189 | 0.38 | 0.83 | -0.02 (-0.22, 0.18) | 1 | 177 | 0.56 | 0.95 | 0.11 (-0.17, 0.39) | 0.89 | 0.71 |
| **Ln IL-12/IL-17A** |  |  |  |  |  |  |  |  |  |  |  |
| Control | 159 | -0.49 | 0.55 |  |  | 154 | -0.64 | 0.65 |  |  |  |
| Nutrition + WSH | 190 | -0.66 | 0.54 | -0.17 (-0.3, -0.04) | 0.02 | 177 | -0.56 | 0.46 | 0.08 (-0.05, 0.2) | 0.44 | <0.01 |
| **Ln IFN-γ/IL-17A** |  |  |  |  |  |  |  |  |  |  |  |
| Control | 159 | 0.6 | 0.47 |  |  | 154 | 0.51 | 0.67 |  |  |  |
| Nutrition + WSH | 190 | 0.42 | 0.62 | -0.18 (-0.31, -0.04) | 0.02 | 177 | 0.52 | 0.43 | 0.01 (-0.12, 0.13) | 1 | 0.05 |
| **Ln IL-12/IL-21** |  |  |  |  |  |  |  |  |  |  |  |
| Control | 156 | 0.59 | 0.97 |  |  | 152 | 0.45 | 1.03 |  |  |  |
| Nutrition + WSH | 186 | 0.55 | 1.13 | -0.04 (-0.31, 0.22) | 1 | 176 | 0.6 | 1.15 | 0.15 (-0.15, 0.44) | 0.67 | 0.47 |
| **Ln IFN-γ/IL-21** |  |  |  |  |  |  |  |  |  |  |  |
| Control | 156 | 1.68 | 1.02 |  |  | 152 | 1.59 | 1.08 |  |  |  |
| Nutrition + WSH | 186 | 1.64 | 1.09 | -0.04 (-0.31, 0.23) | 1 | 176 | 1.68 | 1.1 | 0.08 (-0.22, 0.38) | 1 | 0.94 |
| **Ln Pro-inflammatory cytokines^b^/IL-10** |  |  |  |  |  |  |  |  |  |  |  |
| Control | 158 | 1.83 | 0.59 |  |  | 154 | 1.95 | 0.55 |  |  |  |
| Nutrition + WSH | 189 | 1.71 | 0.65 | -0.12 (-0.28, 0.05) | 0.32 | 171 | 1.89 | 0.74 | -0.06 (-0.2, 0.08) | 0.8 | 1 |
| **Ln Th1^c^/IL-10** |  |  |  |  |  |  |  |  |  |  |  |
| Control | 158 | 1.55 | 0.44 |  |  | 154 | 1.56 | 0.54 |  |  |  |
| Nutrition + WSH | 190 | 1.34 | 0.54 | -0.21 (-0.32, -0.1) | <0.001 | 173 | 1.49 | 0.65 | -0.07 (-0.19, 0.06) | 0.59 | 0.17 |
| **Ln Th2^d^/IL-10** |  |  |  |  |  |  |  |  |  |  |  |
| Control | 158 | 1.47 | 0.6 |  |  | 153 | 1.63 | 0.6 |  |  |  |
| Nutrition + WSH | 189 | 1.35 | 0.67 | -0.11 (-0.26, 0.03) | 0.27 | 173 | 1.44 | 0.75 | -0.18 (-0.34, -0.02) | 0.05 | 1 |
| **Ln Th17^e^/IL-10** |  |  |  |  |  |  |  |  |  |  |  |
| Control | 155 | 0.97 | 0.55 |  |  | 152 | 1.08 | 0.6 |  |  |  |
| Nutrition + WSH | 186 | 0.92 | 0.63 | -0.05 (-0.18, 0.08) | 0.9 | 172 | 0.97 | 0.76 | -0.1 (-0.25, 0.05) | 0.35 | 1 |
| **Ln Th1^c^/Th2^d^** |  |  |  |  |  |  |  |  |  |  |  |
| Control | 158 | 0.08 | 0.42 |  |  | 153 | -0.08 | 0.58 |  |  |  |
| Nutrition + WSH | 189 | -0.02 | 0.54 | -0.1 (-0.21, 0.02) | 0.22 | 177 | 0.03 | 0.47 | 0.11 (-0.03, 0.25) | 0.23 | 0.03 |
| **Ln Th1^c^/Th17^e^** |  |  |  |  |  |  |  |  |  |  |  |
| Control | 156 | 0.58 | 0.41 |  |  | 152 | 0.48 | 0.49 |  |  |  |
| Nutrition + WSH | 186 | 0.42 | 0.51 | -0.16 (-0.28, -0.04) | 0.02 | 176 | 0.51 | 0.46 | 0.03 (-0.09, 0.14) | 1 | 0.02 |

Confidence intervals were adjusted for clustered observations using robust standard errors.

IL = Interleukin; TNF-α = Tumor necrosis factor-α; CRP = C-reactive protein (CRP); IFN-γ = interferon-γ; GM-CSF = Granulocyte-macrophage colony-stimulating factor; AGP = Alpha-1-acid glycoprotein; IGF-1 = Insulin-like growth factor-1

^a^P-values shown are Bonferroni corrected to control for familywise error rates

^b^Pro-inflammatory cytokines: IL-1β, IL-6, TNF-α

^c^Th1 cytokines: IL-12, IFN-γ

^d^Th2 cytokines: IL-4, IL-5, IL-13

^e^Th17 cytokines: IL-17A, IL-21
